# Cerebral perivascular spaces as predictors of dementia risk and accelerated brain atrophy

**DOI:** 10.1101/2024.04.25.24306324

**Authors:** Giuseppe Barisano, Michael Iv, the Alzheimer’s Disease Neuroimaging Initiative, Jeiran Choupan, Melanie Hayden-Gephart

**Affiliations:** Department of Neurosurgery, Stanford University, Stanford, CA, USA; Department of Radiology, Stanford University, Stanford, CA, USA; Laboratory of Neuro Imaging, University of Southern California, Los Angeles, CA, USA

## Abstract

Cerebral small vessel disease, an important risk factor for dementia, lacks robust, *in vivo* measurement methods. Perivascular spaces (PVS) on brain MRI are surrogates for small parenchymal blood vessels and their perivascular compartment, and may relate to brain health. We developed a novel, robust algorithm to automatically assess PVS count and size on MRI, and investigated their relationship with dementia risk and brain atrophy. We analyzed 46,478 clinical measurements of cognitive functioning and 20,845 brain MRI scans from 10,004 participants (71.1±9.7 years-old, 56.6% women). Fewer PVS and larger PVS diameter at baseline were associated with higher dementia risk and accelerated brain atrophy. Longitudinal trajectories of PVS markers were significantly different in non-demented individuals who converted to dementia compared with non-converters. In simulated placebo-controlled trials for treatments targeting cognitive decline, screening out participants less likely to develop dementia based on our PVS markers enhanced the power of the trial. These novel radiographic cerebrovascular markers may improve risk-stratification of individuals, potentially reducing cost and increasing throughput of clinical trials to combat dementia.

## Introduction

Cerebral small vessel disease, the most common vascular disease affecting the blood vessels within the brain parenchyma, is considered a significant and potentially reversible contributor to cognitive decline and dementia.^1,2^ *Ex vivo* studies have shown an independent association of cerebral small vessel neuropathology with cognitive function and many subtypes of dementia.^3–7^ Given the known association with dementia risk, it is important to identify robust radiographic markers of the cerebral small vessels, to properly risk-stratify patients, monitor disease, and assess the impact of treatment.

*In vivo* evaluation of cerebral small vessel disease relies on magnetic resonance imaging (MRI) and includes different signs of brain parenchymal damage (e.g., white matter lesions, lacunes, subcortical infarcts, cerebral microbleeds, and enlargement of perivascular spaces).^8^ Yet these MRI findings may not be detectable in the healthy population, lack quantitative granularity, require specific MRI sequences, or are labor intensive to process.^1,2^ Using the perivascular space as detected on MRI to non-invasively evaluate cerebral small vessel health requires only an unenhanced 3-dimensional T1-weighted images,^9^ a nearly universal brain MRI sequence. Current techniques of perivascular space estimation^10–15^ though suffer from user-dependency, and weak or unknown robustness and inter-scanner reproducibility^1,9,16–18^. These limitations prevent generalized, widespread use in the hospital, clinical trials, and multi-center longitudinal studies. New, robust tools, therefore, are needed to investigate the temporality and nonspurious dose-response relationship between perivascular spaces (as a surrogate of cerebral small vessels) and dementia.

To meet this critical need, we developed and validated a novel, fully automated approach to robustly assess the perivascular spaces in the white matter (WM-PVS) and basal ganglia (BG-PVS) requiring as input only unenhanced 3-D T1-weighted images. Despite some possible limitations in PVS visual detection compared with T2-weighted images,^8^ 3-D T1-weighted images are more commonly included in brain MRI protocols.^19^ We employed our novel method to investigate the association of baseline PVS with the risk of developing dementia and the brain atrophy trajectory. We focused on two properties: the number of perivascular spaces on T1-weighted image (hereinafter referred to as PVS count), and PVS mean diameter. We also included in our analyses the (log-transformed) volume of white matter lesions in the periventricular area (P-WML) and deep white matter (D-WML), since they can similarly be measured on T1-weighted images and are considered closely related to PVS.^8^

Given that the inclusion of individuals with low risk of cognitive decline reduces the power of clinical trials evaluating a treatment effect on cognitive impairment,^20^ we evaluated the potential benefit of our novel PVS markers as a screening tool to enrich for individuals likely to develop dementia and compared their performance with that of standard WML and atrophy markers assessed on T1-weighted images.

## Results

### Technical validation of PVS and WML markers

The spatial overlap between the PVS masks obtained with our novel fully automated approach and those obtained with a previously validated semi-automated technique^12,13,17^ was very high (average Dice similarity coefficient: 0.95 ± 0.0001) and the numbers of PVS voxels identified with the two methods were strongly correlated (Spearman’s ρ=0.94, P<0.001, Fig. S3A). Consistently, PVS measured with our method also showed a strong positive association with age, male sex, and body mass index in the Human Connectome Project dataset (Fig. S3B-D), replicating results previously published with the validated semi-automated techniques.^21–23^ These data support the reliability and accuracy of our PVS masks. While previous methods are user-dependent (i.e., the user needs to identify a threshold for each type of T1-weighted image) and lack inter-scanner reproducibility of PVS markers^1,9,16–18^ (Fig. S1A-B), our technique is able to provide measurements of PVS count and diameter in a fully automated and robust way. Indeed, both metrics showed excellent intraclass correlation coefficients (≥0.9 for WM-PVS and ≥0.8 for BG-PVS) for inter-scanner reproducibility, inter-field-strength reproducibility, and test-retest repeatability (Figure S4). We also observed a strong correlation between the numbers of PVS voxels independently identified by our algorithm on the T1- and T2-weighted images (Spearman’s ρ=0.98, P<0.001, Fig. S5A), with an average of 87.3±5.7% of PVS voxels detected on T1-weighted images that overlapped with PVS voxels on T2-weighted images. Overall, these data show that our method can accurately segment PVS on T1-weighted images and that T1-weighted images are suitable to reliably assess PVS morphological metrics and inter-subject differences. In contrast with previous methods,^10–15^ our new approach for PVS segmentation is fully automated, requires only T1-weighted images, and provides robust metrics across different scanners and protocols (Figure S4).

Concerning the segmentation of WML, which we performed with a previously validated technique,^24^ we confirmed that T1-weighted images, despite considered less sensitive than FLAIR images for the detection of WML,^8^ allow to obtain valid estimates of WML volume, as the number of WML voxels detected on T1-weighted images were strongly correlated with those detected on FLAIR (Spearman’s ρ=0.82, P<0.001, Fig. S5B), and 83.0±0.4% of WML voxels detected on T1-weighted images spatially overlapped with WML voxels detected on FLAIR. The derived measurements of WML volume also showed excellent intraclass correlation coefficients (≥ 0.95 for P-WML, ≥ 0.83 for D-WML) for inter-scanner reproducibility, inter-field-strength reproducibility, and test-retest repeatability (Figure S6).

### Study population

The baseline characteristics of the 10,004 participants stratified by study cohort are reported in Table 1. There were 8867 non-demented subjects and 1137 patients with dementia, including 996 clinically diagnosed with probable or possible Alzheimer’s dementia, 51 Lewy body dementia, 38 frontotemporal dementia, 1 vascular dementia, and 51 other or uncertain type of dementia. Accuracy of the PVS masks were visually verified for all the cases in a blinded fashion by an expert physician-scientist according to established criteria.^8^ We also developed an interactive website that allows the readers to visualize our PVS segmentations and independently verify their accuracy: https://gbarisano.shinyapps.io/pvs-dementia.

**Table 1.**
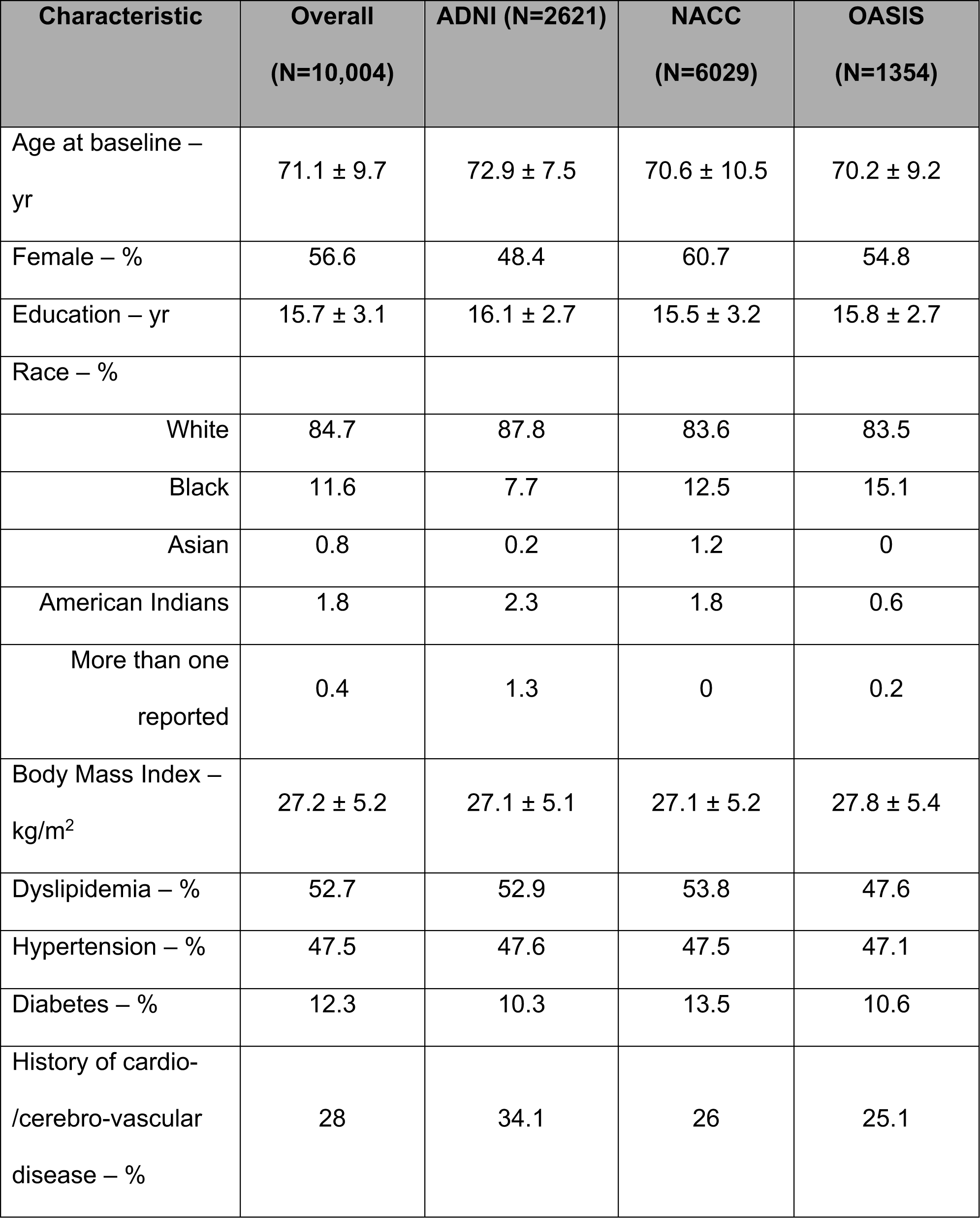
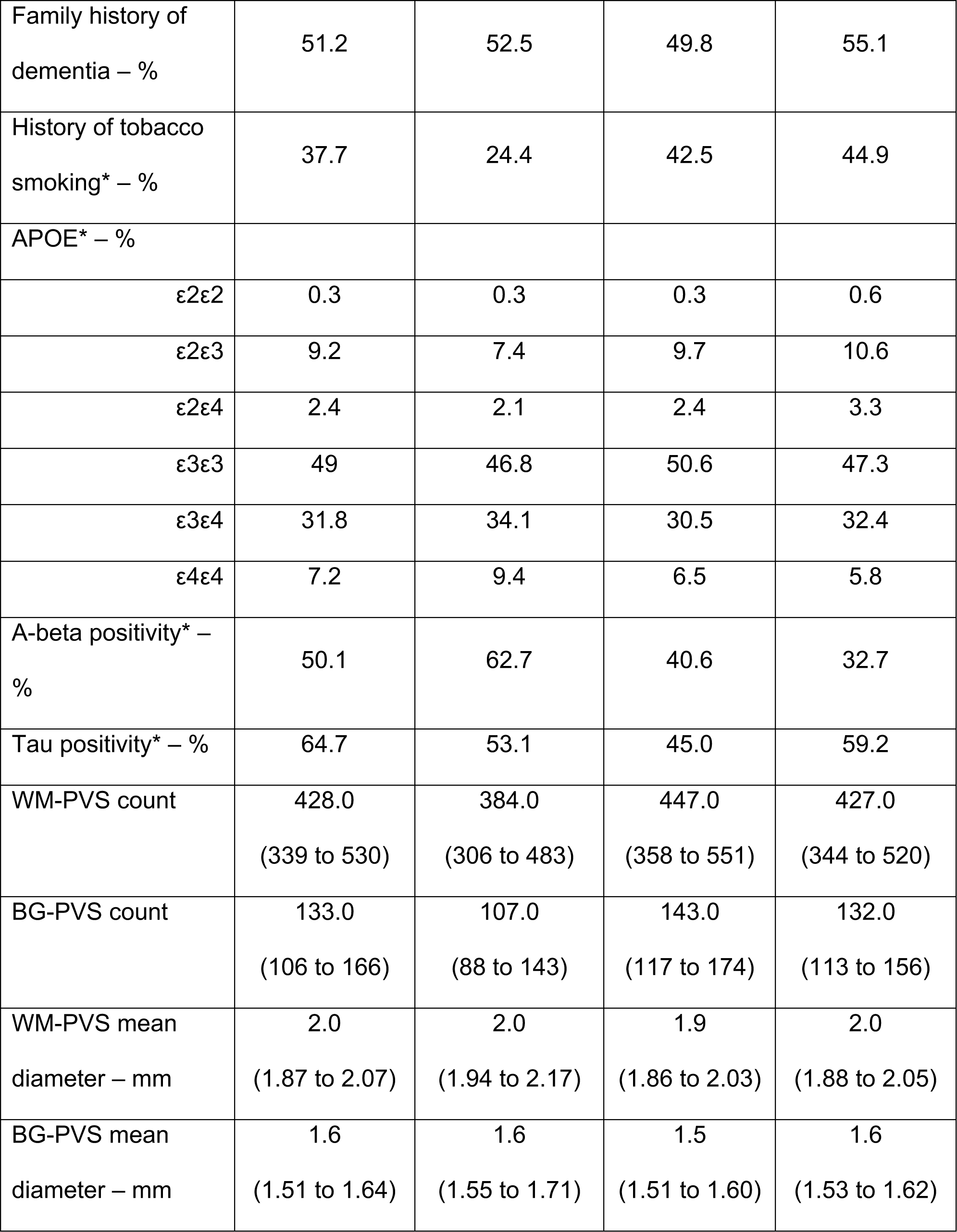
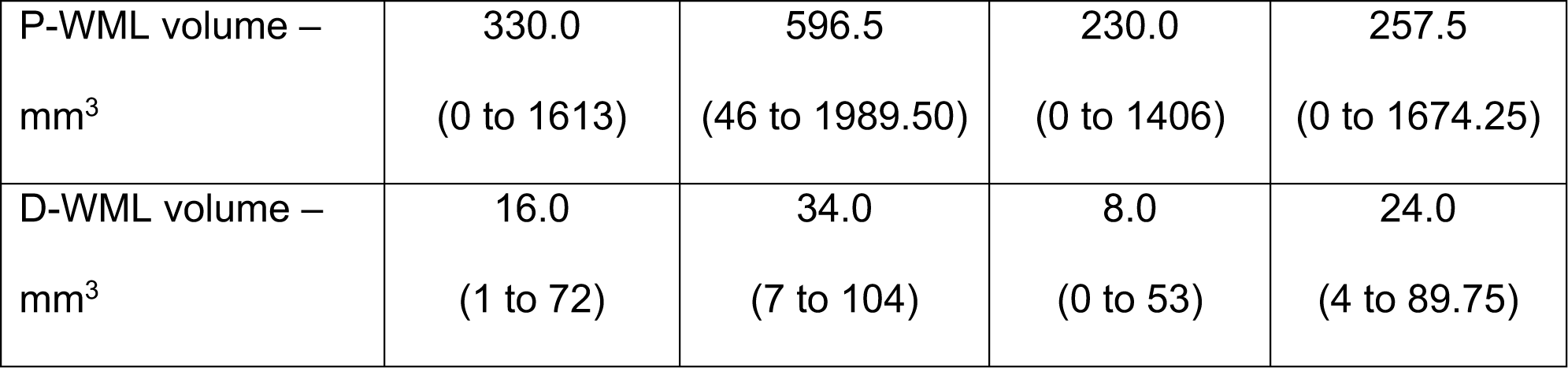
Baseline characteristics of the study population. Plus–minus values are means ± standard deviations, entries with parentheses are medians (interquartile range). Data on education were missing for 0.15% of the participants, on race for 0.8%, on body mass index for 2.6%, on dyslipidemia for 1.2%, on hypertension and diabetes for 0.7%, on history of cardio-/cerebro-vascular disease on 0.5%, on family history for dementia for 2.7%. Data from three studies — the Alzheimer’s Disease Neuroimaging Initiative (ADNI), the National Alzheimer’s Coordinating Center (NACC), and the Open Access Series of Imaging Studies (OASIS) — are shown. Race was reported by the participant. *Total number of subjects with available history of tobacco smoking, Apolipoprotein E (APOE) genotype, amyloid-beta status and tau status were 9267, 8599, 3906, and 2466, respectively.

In multivariate analyses, all PVS and WML markers were significantly associated with age, sex, body mass index, cognitive scores, and history of hypertension (Table S2). History of diabetes was associated with all PVS markers except BG-PVS diameter; history of cardiovascular disease was associated with BG-PVS diameter. In the subgroup analysis (Table S3), WM-PVS count was not significantly associated with APOE genotype, amyloid or tau status, and BG-PVS count was negatively associated with amyloid status and APOE-ε4ε4-carrier status. WM-PVS diameter, P-WML and D-WML volume were associated with positive amyloid and APOE-ε4ε4-carrier status. BG-PVS diameter was associated with positive tau and APOE-ε4ε4-carrier status.

After controlling for study cohort, demographic, and clinical variables, demented patients showed significantly lower WM- and BG-PVS count, higher WM- and BG-PVS diameter, and higher P-WML volume compared with non-demented subjects (estimated marginal means in Table S4). Consistent results for WM-PVS count, BG-PVS count, and P-WML were obtained in sensitivity analyses (Table S5), indicating that their differences are independent of APOE, amyloid, or tau status.

### PVS markers and risk of dementia

Among 7518 non-demented participants with at least 1 follow-up visit (Table S6), 1493 individuals developed dementia during a median follow-up of 4.1 years (interquartile range, 2.3-7.1), including 1306 clinically diagnosed with probable or possible Alzheimer’s dementia, 43 Lewy body dementia, 29 frontotemporal dementia, 16 vascular dementia, and 99 other or uncertain type of dementia. In the fully adjusted models of the two-stage pooled analysis, risk of dementia was significantly associated with baseline WM-PVS and BG-PVS count and diameter. Each additional 100 WM-PVS and 10 BG-PVS were significantly associated with 20% and 9% decreased dementia risk, respectively, and each additional 0.1-mm increase in WM-PVS and BG-PVS mean diameter were significantly associated with 9% and 20% increased dementia risk, respectively (Fig. 1A-D). P-WML, but not D-WML, was also significantly associated with dementia risk (Fig. 1E-F). The spline analysis with pooled data supported a linear association over the range of the PVS and P-WML markers measured in this population (Fig. 1G-K). Sensitivity analyses showed substantially unchanged results, suggesting that these effects are independent of other potential confounding factors, including the positivity status for amyloid or tau biomarkers (Table S7 and Fig. S7). The association of WM-PVS diameter with dementia risk became not statistically significant after adding amyloid status in the model.

**Figure 1.**
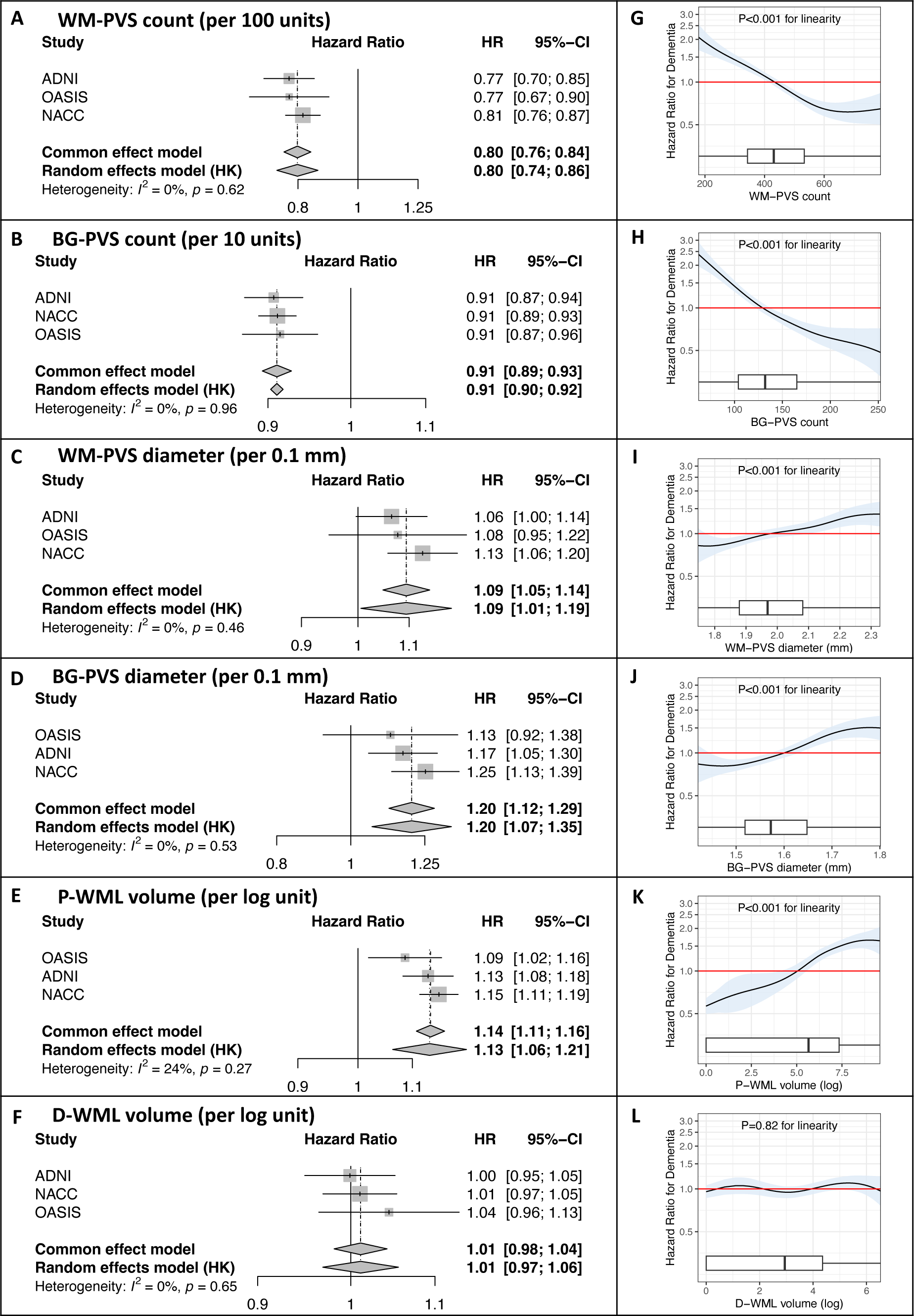
Forest plots and spline plots for the Associations of PVS and WML markers with Dementia Risk. In two-stage pooled analyses that combined individual-participant data from three studies (Panels A-F), each additional 100 WM-PVS (Panel A) and 10 BG-PVS (Panel B) were associated with 20% and 9% decrease in dementia risk, each additional 0.1 mm increase in mean WM-PVS ( Panel C) and BG-PVS (Panel D) diameter were associated with 9% and 20% increase in dementia risk, and each unit increase of the log-transformed P-WML volume (Panel E) was associated with 13% increase in dementia risk. Log-transformed D-WML volume (Panel F) was not associated with dementia risk. In each graph, the size of the squares indicates the weight given to the study, and the width of the diamond indicates the 95% confidence interval for the overall association estimate. Between-study heterogeneity was statistically assessed with the use of I^2^. The spline analysis of pooled data (Panels G-L) supported a linear association over the range of WM-PVS count (Panel G; 2.5^th^-97.5^th^ percentile, 206 to 762), BG-PVS count (Panel H; 2.5^th^-97.5^th^ percentile, 71 to 244), WM-PVS diameter (Panel I; 2.5^th^-97.5^th^ percentile, 1.77 to 2.30 mm), BG-PVS diameter (Panel J; 2.5^th^-97.5^th^ percentile, 1.44 to 1.79) and P-WMH volume (Panel K; 2.5^th^-97.5^th^ percentile, 0 to 9.1) within the overall population. Shaded areas indicate 95% confidence intervals, and the red line at 1.0 indicates the reference. Box plots at the bottom of the graphs show the distributions of the marker. The vertical bar indicates the median, and the ends of the box the interquartile range; the whiskers extend to values no farther than 1.5 times the interquartile range (which may be past the graphed area). Data from three studies — the Alzheimer’s Disease Neuroimaging Initiative (ADNI), the Open Access Series of Imaging Studies (OASIS), and the National Alzheimer’s Coordinating Center (NACC) — are shown. Hazard ratios were estimated from Cox models stratified according to study cohort with adjustment for age, sex, race, educational level, body mass index, CDR global score at the baseline, history of diabetes, cardio-/cerebro-vascular disease, hypertension, dyslipidemia, family history of dementia, intracranial volume and the time interval between the MRI scan and the clinical visit of the cognitive assessment at the baseline.

### PVS markers and brain atrophy

Among 3389 non-demented participants with at least 2 MRI scans (Table S8), we investigated the association of PVS and WML markers measured at the baseline MRI with the trajectory of brain atrophy estimated over a total of 14,229 MRI scans (average of 4 scans available per subject) during a median follow-up time of 3.1 years (interquartile range 2.0 to 5.6 years). In the mixed-effect models fully adjusted for demographic, clinical, and MRI-related covariates, the association of PVS and WML markers with the longitudinal trajectory of brain atrophy were statistically significant (Fig. 2A, Table S9). Each additional 100 WM-PVS and 10 BG-PVS identified at the baseline were associated with preservation of additional 709±100 mm^3^ and 187±33 mm^3^ of total grey matter volume every year, respectively. Each 0.1 mm increase in WM-PVS and BG-PVS diameter was associated with loss of additional 382±84 mm^3^ and 576±137 mm^3^ of total grey matter volume every year, respectively. Each logarithmic unit increase in P-WML and D-WML volume was associated with loss of additional 288±46 mm^3^ and 189±62 mm^3^ of total grey matter volume every year, respectively. Similar results were obtained for cortical thickness (Fig. 2B, Tables S9). Most of the effects for the grey matter volume and cortical thickness were observed in the temporal lobes, bilaterally, for all markers (Fig. 2A-B bottom rows, respectively; Tables S10 and S11, respectively). For white matter volume, only WM-PVS count, BG-PVS count and BG-PVS diameter were significantly associated with accelerated atrophy (Fig. 2C, Tables S9). These effects and their spatial patterns remained significant for most markers in all sensitivity analyses, indicating that the link between these markers and accelerated atrophy is independent of other potential confounding factors, including the positivity status for amyloid or tau biomarkers (Figures S8-S10, Table S13). Exceptions included WM-PVS and BG-PVS diameter, which became not significant after adding tau status in the model for grey matter volume (Fig. S8, Table S13), but remained significant for cortical thickness (Fig. S9, Table S13); BG-PVS diameter became not significant after adding amyloid status in the model for white matter volume (Fig. S10, Table S13). Overall, these data showed that fewer WM-PVS and BG-PVS counts were associated with accelerated brain atrophy. This effect was stronger on the grey matter volume and cortical thickness of the temporal lobes, irrespective of amyloid and tau positivity status. Similar associations were found for WM-PVS and BG-PVS diameter, although they were generally less robust when considering amyloid and/or tau status in the model.

**Figure 2.**
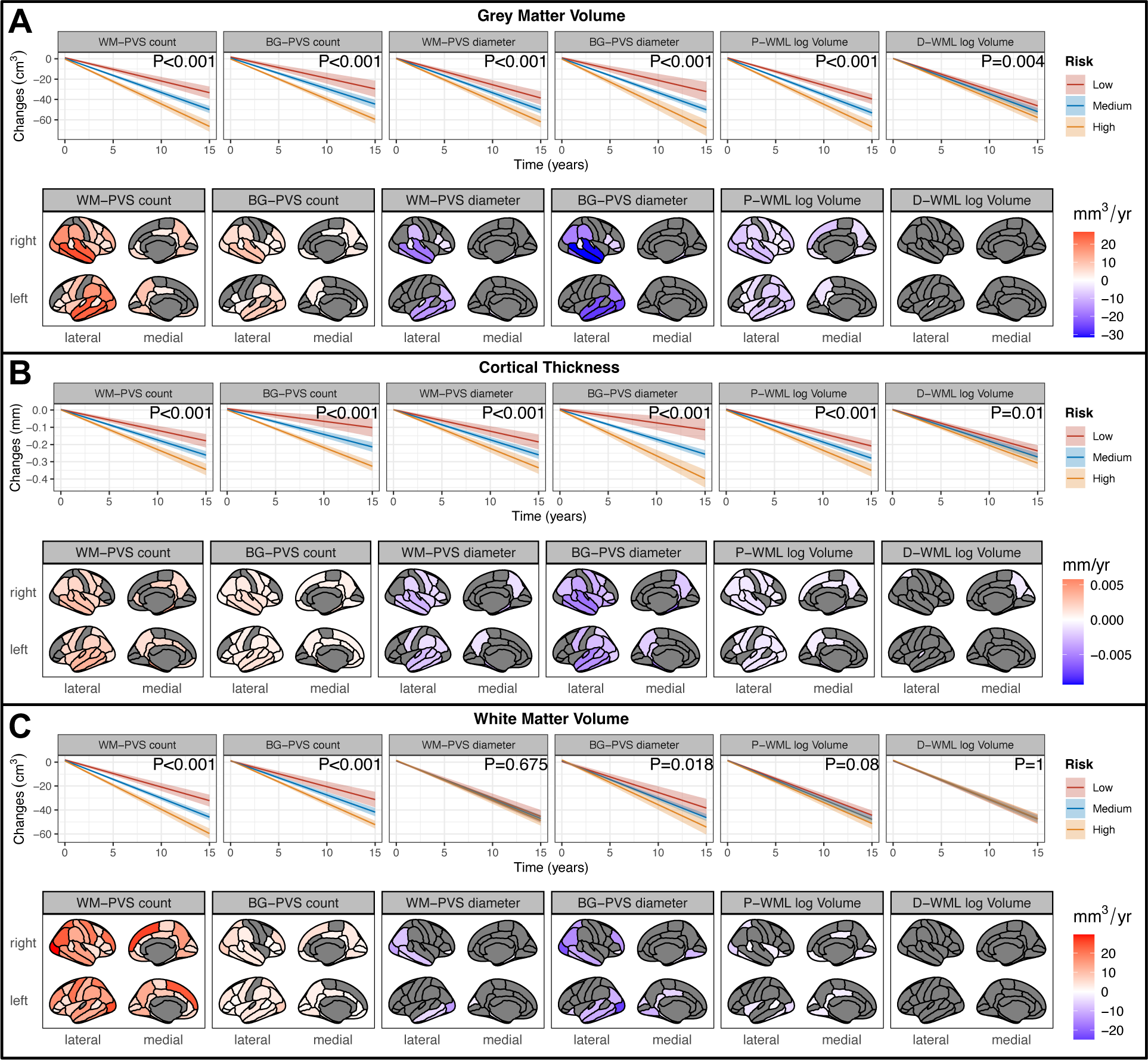
Plots for the estimated trajectories of brain atrophy in relation to PVS and WML markers. Top rows in each panel depict the effect of baseline PVS and WML markers on the trajectories of grey matter volume, cortical thickness, and white matter volume (Panels A-C, respectively). For each marker, equally spaced values from the low-risk (red), medium-risk (blue), and high-risk (yellow) tertile are shown (tertile limits in Table S16). Shaded areas indicate 95% confidence intervals. The regional analysis (bottom rows in each panel) across cortical parcellations according to the Desikan-Killiany atlas reports the estimated volume or thickness preserved (positive values in red) or lost (negative values in blue) per year for each additional unit increase in the vascular marker. Only estimated values for regions that remained statistically significant after correction for multiple comparisons (68 comparisons) are shown; non-significant regions are greyed out. See also Tables S11-13 for grey matter volume, cortical thickness, and white matter volume, respectively. Estimates and corrected significance (P values) obtained from fully adjusted linear mixed-effects models with random intercepts and slopes for each individual participant (N=3389 and 14,229 timepoints MRI scans). All models were adjusted for age, sex, race, educational level, body mass index, CDR global score at the baseline, history of diabetes, cardio-/cerebro-vascular disease, hypertension, dyslipidemia, family history of dementia, intracranial volume, the baseline value of the dependent variable (grey matter volume, cortical thickness, or white matter volume), field strength, manufacturer, and intra-individual consistency of the protocol used for the longitudinal MRI acquisitions (consistent versus non-consistent protocol).

We further assessed the relationship between the WM-PVS marker measured in a specific lobe and the corresponding atrophy in that lobe. The spatial patterns for regional WM-PVS count (Fig. S11, left column) were consistent with those observed in the main model with global WM-PVS count (Fig. 2). Less consistency was observed for regional WM-PVS diameter (Fig. S11, right column). This suggests a spatial relationship between the atrophy trajectory of a specific brain region and the corresponding WM-PVS count in that region at baseline.

Based on previous results from preclinical studies^25,26^, we hypothesized that one potential mechanism linking PVS with accelerated brain atrophy could be alterations in cerebral blood flow. Indeed, in most of the brain regions we observed a significant correlation of the regional PVS count and diameter with the corresponding regional cerebral blood flow as assessed *in vivo* with arterial spin labeling MRI (Fig. S12). This suggested that the observed effects of PVS on accelerated brain atrophy might have been related to decreased blood flow. However, the relatively low sample size of subjects with cerebral blood flow data available at the baseline (N=224) prevented us from building reliable linear mixed-effects models evaluating the relative contribution of cerebral blood flow and PVS to the brain atrophy trajectory while accounting for the relevant clinical and demographic covariates included in all our models. No significant correlations with cerebral blood flow were found for P-WML and D-WML (Fig. S12).

Finally, we estimated the longitudinal trajectory of the PVS and WML markers in non-demented individuals who converted to dementia and compared it with those who did not convert. The WM and BG regions where PVS were measured were spatially-registered and kept consistent across the intra-individual timepoints. This means that, for each subject, PVS were analyzed in the exact same voxels across timepoints. The longitudinal trajectories of PVS count and mean diameter significantly differed between converters and non-converters, both in the white matter and in the basal ganglia: PVS count in converters, who already had lower PVS count compared with non-converters at the baseline MRI (Table S14), further decreased with time. PVS count remained stable in non-converters (Figure 3A and C). On the other hand, WM-PVS and BG-PVS mean diameter (Figure 3B and D, respectively) were stable in converters, who already had significantly higher baseline PVS diameter compared with non-converters (Table S14). WM-PVS and BG-PVS mean diameter decreased in non-converters. Sensitivity analyses showed consistent results (Figure S13), indicating that the different trajectories for these markers in converters versus non-converters were independent of other potential confounding factors such as amyloid and tau status. In WM-PVS markers, the differences between converters and non-converters involved mostly the left hemisphere, especially the frontal and parietal lobes (Figure 3A and B). Significant increases over time in P-WML volume were also found, which were more prominent in converters compared with non-converters (Figure 3E). Overall, these data indicate that the differences in PVS count and diameter measured at the baseline MRI scans of dementia converters versus non-converters followed different longitudinal trajectories: in converters, larger baseline PVS remain enlarged, and lower baseline PVS count continue decreasing.

**Figure 3.**
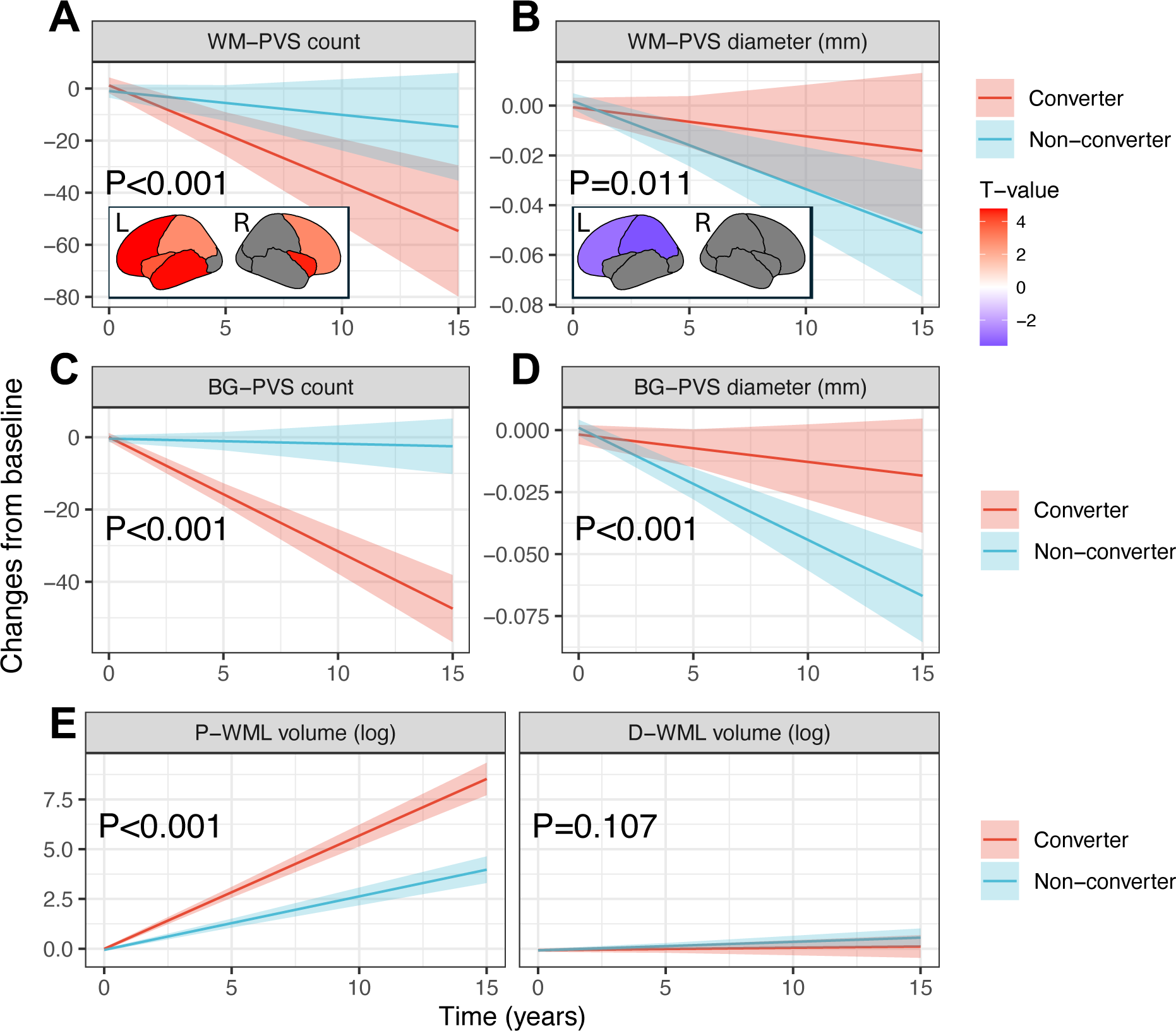
Plots for the estimated longitudinal trajectories of PVS or WML markers according to conversion to dementia status. Estimated longitudinal trajectories of WM-PVS (A-B), BG-PVS (C-D), and WML (E) markers for non-demented individuals who converted to dementia (converters, red) and those that did not convert to dementia (non-converters, cyan). Trajectories are estimated from fully adjusted linear mixed-effect models; the adjusted p-values (P) indicate whether the trajectories are significantly different between the groups after correction for multiple comparisons. Shaded areas indicate 95% confidence intervals. For WM-PVS markers (panels A-B), we estimated in each lobe of the left (L) and right (R) hemispheres the group-effect (expressed as T-value) for the longitudinal trajectories of the corresponding marker: positive values in red indicate significantly higher (i.e., less negative) slopes for non-converters versus converters, whereas negative values in blue indicate significantly lower (i.e., more negative) slopes for non-converters versus converters. Lobes where the longitudinal trajectories were not significantly different between converters and non-converters after correction for multiple comparisons are greyed-out. Estimates and corrected significance obtained from fully adjusted linear mixed-effects models with random intercepts and slopes for each individual participant (N=3389 and 14,229 timepoints MRI scans).

### PVS markers in simulated clinical trials

In 48-month placebo-controlled trials simulated with 40,703 cognitive assessments in 7518 non-demented participants, the sample size required to detect a 30% slowing in cognitive decline (assessed with the CDR) with 80% power was lower when selectively screening out participants based on the PVS or the WML markers’ tertiles (Table S16): sample size reductions were 13-37% when enrolling participants in the medium- and high-risk tertiles (Figure 4, red bars), and 28-63% when enrolling participants in the high-risk tertile only (Figure 4, blue bars). The performance was comparable to that observed for the atrophy markers cortical thickness and grey matter volume (respectively 53% and 37% reductions when including individuals in the high-risk tertile). Similar results were obtained with simulations where the cognitive decline was assessed with the MMSE (Fig. S14).

**Figure 4.**
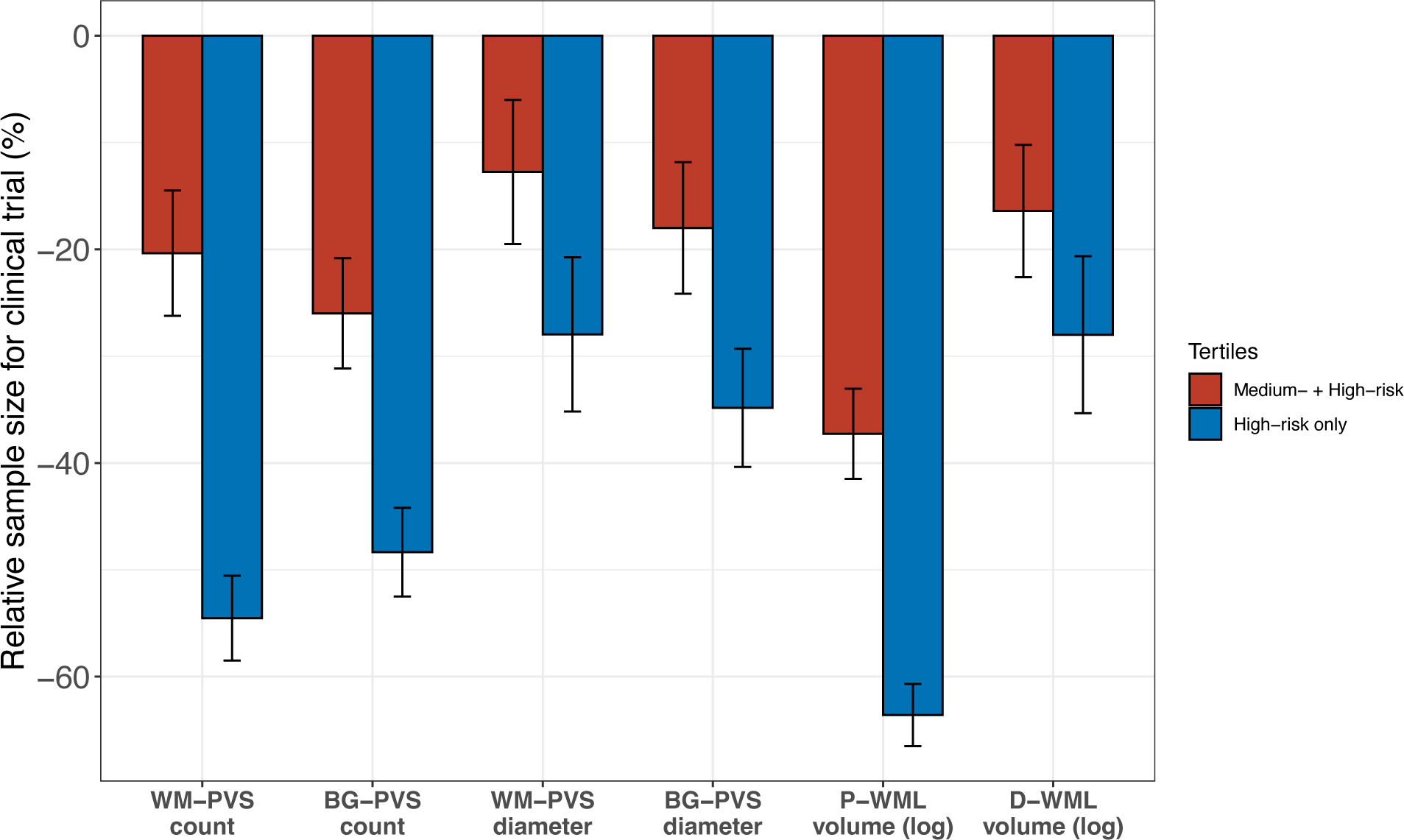
Barplots for the relative sample size in simulated clinical trials enriched using PVS or WML markers. Relative sample sizes for simulated clinical trials in non-demented individuals pooled from three studies (the Alzheimer’s Disease Neuroimaging Initiative, the Open Access Series of Imaging Studies, and the National Alzheimer’s Coordinating Center). The simulations had statistical power of 80% at α = .05 and assumed a 30% treatment effect for slopes in cognitive decline, 1:1 allocation of treatment, total trial length of 48 months, and outcome measures every 12 months. All available longitudinal cognitive data (40,307 cognitive assessments from 7518 non-demented subjects) were used in these fully-adjusted models. All estimates and standard errors are across 500 bootstrap iterations. The reference model (without enrichment and with 100% inclusion) included all the tertiles. In the two enrichment models for each marker (“medium-risk+high-risk tertiles”, red bars; “high-risk only tertile”, blue bars), only participants in the indicated tertiles were included. Tertile limits for each marker are reported in Table S16.

After controlling for demographic covariates, medium- and high-risk tertiles for PVS markers were not significantly associated with amyloid or tau positive status (Table S15), suggesting that screening non-demented participants according to our PVS markers is not linked to Alzheimer disease biomarkers. On the other hand, medium- and high-risk tertiles for P-WML volume, cortical thickness, and grey matter volume were associated with amyloid positive status (Table S15).

## Discussion

We developed a novel, fully automated, and robust algorithm to obtain unbiased, quantitative metrics of PVS from clinical brain MRI T1-weighted images. We demonstrated that our method provides accurate segmentations with high inter-scanner and inter-field-strength reproducibility. These characteristics allowed us to apply this algorithm to the brain MRI scans of 10,004 subjects whose data were pooled from three publicly available studies performed in the United States and Canada. We found that after controlling for demographic and clinical covariates, lower PVS count and higher mean PVS diameter were significantly associated with a dose-response higher risk of developing dementia, and with accelerated brain atrophy. The robustness and reliability of these results is related to the large sample size, the use of individual-level data and consistent approaches for covariate adjustments and modeling, and the consistency across multiple sensitivity analyses. These data show that PVS may represent a predictor of dementia.

Perivascular spaces represent a critical component of the glymphatic system, a system thought to be responsible for brain clearance of toxic and waste metabolites.^27^ A larger perivascular diameter might indicate alterations in the glymphatic flow and impairment of the clearance process, with subsequent accumulation of neurotoxic protein aggregates in the brain.^25,28,29^ In agreement with preclinical studies showing a relationship between glymphatic system and blood flow^25,26^, we found that PVS diameter was inversely correlated with cerebral blood flow. On the other hand, PVS count was positively correlated with brain perfusion, with a higher PVS count also associated with lower risk of dementia and slower brain atrophy. Since the PVS count obtained with our method included any MRI-visible PVS, regardless of whether they could be considered enlarged or not, PVS count may be linked to brain perfusion rather than to glymphatic dysfunction. A low PVS count might indicate cerebral hypoperfusion, a factor found associated with cognitive decline and brain atrophy.^30–33^

Studies prior to ours performed manual visual assessment of perivascular spaces, a significant impediment for large studies and clinical trials, with serious concerns regarding inter- and intra-rater variability. This has led to mixed conclusions on the relationship between PVS and dementia. For example, previous meta-analyses investigated the relationship between WM-PVS and risk of dementia, but the results were conflicting, with significant heterogeneity between studies, variability in methods, and inconsistent adjustment for confounding factors.^1,34,35^ Other studies in smaller cohorts also reported diverging results: some showed an association between perivascular space enlargement and higher risk of dementia,^36,37^ while others found no increased risk^38^ or increased risk of vascular dementia only.^39^ Importantly, in all these studies the manual visual assessment prevented a quantitative analysis of the blood vessels with small/non-enlarged perivascular spaces, a limitation eliminated by our novel algorithm. Moreover, visual readings typically do not consider in their assessment the total intracranial volume, a factor strongly associated with PVS^9^ and that we controlled for in all our models.

Our novel MRI PVS markers required only a commonly acquired volumetric T1-weighted sequence, were computed in a fully-automated fashion, and showed excellent inter-scanner and test-retest reproducibility. These features may allow for our PVS markers to be readily implemented in clinical practice, as well as retrospective analyses of currently available brain MRI data. As we showed in our clinical trial simulations, their use may reduce the cost and duration of clinical trials for dementia prevention and treatment by facilitating the identification and enrollment of individuals with increased risk of cognitive decline. Indeed, we found that after screening out participants based on our PVS markers, there was a substantial reduction in the minimal number of individuals required for detecting the intervention effect, suggesting an increase in the power of the trial. Importantly, medium and high-risk tertiles of our PVS markers were not linked to positive amyloid and tau status, in contrast with WML and atrophy markers, indicating that the selection of individuals based on our PVS markers is independent of Alzheimer disease biomarker status. In the case of trials for treatments specifically targeting Alzheimer disease hallmark pathology such as amyloid-β and tau (i.e., enrolling individuals positive for amyloid-β and/or tau), these PVS markers would allow to identify and exclude individuals at high risk of vascular/perivascular co-pathology, enhancing the trial for a more homogenous cohort of preclinical Alzheimer disease. Early detection of an increased risk of dementia could also motivate individuals to adopt healthy lifestyle modifications, encouraging healthcare professionals to implement preventive measures and initiate timely treatments and support, thereby improving patient and family member quality of life. Finally, these novel MRI markers open new opportunities to robustly investigate perivascular spaces *in vivo* in a variety of other neurological conditions and treatment paradigms, allowing to gain new insights on the human brain vasculature and glymphatics.

We acknowledge the following limitations in our study. First, our analytic approach allowed us to reduce the degree of heterogeneity across studies, but some heterogeneity remained, intrinsic to the original study design, such as the health profile of participants and availability and type of information on covariates. Second, we could not assess the potential role of residual confounding factors, such as diet and socioeconomic status, which were not available in the analyzed databases. Third, as we were restricted to populations recruited by the original studies, our cohort comprised mainly white participants, and increased representation of other racial and ethnic groups would be critical to generalization. Fourth, we cannot exclude that small lesions with vascular shape may be included in our PVS segmentation masks. Nevertheless, the influence of these potential lesions to our statistical analyses may be considered negligible, since even a few lesions in a single subject would represent a very small proportion of the total PVS count estimated in that subject (interquartile range of WM-PVS and BG-PVS per subject in our study were 339-530 and 106-166, respectively). Finally, we could not discriminate whether our PVS metrics refer to periarteriolar or perivenous compartment. However, previous studies have shown that the majority of the MRI-visible PVS overlap with arterioles,^40–42^ therefore it is reasonable to assume that the PVS metrics in our analysis refer mostly to arterioles and their periarteriolar compartment.

In conclusion, using our novel, fully-automated, robust algorithm for assessing perivascular spaces in the cerebral white matter and basal ganglia, we found a significant linear association of low PVS count and high PVS diameter with increased risk of dementia and accelerated brain atrophy. These results support a link between PVS and cognitive impairment, opening new opportunities to risk-stratify individuals for clinical trial enrollment, early healthcare interventions to combat dementia, and accelerated research in human brain glymphatics.

## Online Methods

### Study Population

We combined data from three observational community-based studies: the Alzheimer’s Disease Neuroimaging Initiative (ADNI)^43^ (MRI data downloaded on March 2023), the National Alzheimer’s Coordinating Center (NACC) database^44^ (December 2022 data freeze), and the Open Access Series of Imaging Studies (OASIS-3)^45^ (Data release 2.0, July 2022). Enrolled subjects include both cognitively unimpaired individuals and patients with cognitive impairment and dementia. All participants undergo standardized clinical and neuropsychological examination. All the studies obtained institutional review board approval and written informed consent from all participants.

We included in our analysis all participants who underwent 1) at least one clinical visit with cognitive assessment data available and 2) a brain MRI scan including an unenhanced 3-dimensional T1-weighted sequence within 12 months from the baseline clinical visit. In the analysis investigating the risk of dementia, and in the simulated clinical trials, only baseline non-demented subjects with at least 1 follow-up clinical visit were included (definition of dementia is described in the following section). In the analysis of PVS and brain atrophy trajectories assessed with MRI, only baseline non-demented subjects with at least 1 follow-up MRI scan were included.

Specific details on the individual study designs and enrollment criteria are provided below.

#### ADNI

ADNI is a longitudinal multicenter study conducted in the United States and Canada to develop clinical, imaging, genetic, and biochemical biomarkers for the early detection and tracking of Alzheimer’s disease (AD). ADNI began in 2004 and enroll cognitively unimpaired or impaired subjects between 55 and 90 years of age (inclusive) with a study partner to provide an independent evaluation of functioning.^46^ All subjects could not have any medical contraindications to Magnetic Resonance Imaging (MRI), could not be enrolled in other trials or studies concurrently, and could not take any medication that could affect cognitive function.^46^ All subjects had to have Hachinski Ischemic Score of less than or equal to 4;^47^ permitted medications stable for 4 weeks prior to screening; a Geriatric Depression Scale score of less than 6;^48^ a study partner with 10 or more hours per week of contact either in person or on the telephone and who could accompany the participant to the clinical visits; visual and auditory acuity adequate for neuropsychological testing; good general health with no diseases precluding enrollment; 6 grades of education or work history equivalent; and ability to speak English or Spanish fluently. Women had to be sterile or 2 years past childbearing potential.

At the screening visit, all subjects were required to provide informed consent as compatible with the local sites (Institutional Review Board regulations). In addition, all subjects provided demographics, family history, and medical history. All subjects were given a physical examination and a neurologic examination, and vital signs were recorded. The haplotype of apolipoprotein E (APOE) gene was assessed on blood samples. Cerebrospinal fluid samples were collected in a subsample of the participants: Amyloid-β_1-42_, total Tau, and phosphorylated Tau_181_ measurements were completed using the Roche Elecsys Cobas E601 fully automated immunoassay platform at the ADNI biomarker core (University of Pennsylvania) and were available for 61.3%, 61.2%, and 61.2% of the participants included in our analysis.

Data on amyloid tracers uptake on Positron Emission Tomography (PET), including florbetapir (AV-45), florbetaben (FBB), and Pittsburgh compound B (PiB), were available in 66.7% of participants. Quantitative measurements of standardized uptake value ratio (SUVR) were provided by the ADNI PET core (University of California, Berkeley) following protocols described in the ADNI website (https://adni.loni.usc.edu/methods/pet-analysis-method/).

Participants were classified as amyloid-positive based on abnormal amyloid level detected on cerebrospinal fluid (<1098 pg/ml)^49^ or PET (thresholds recommended in the ADNI PET core documentation: AV-45>1.11,^50^ FBB>1.08,^51^ PiB>1.5^52^), and as tau-positive based on abnormal level of total tau (>242 pg/ml) or phosphorylated tau (>19.2) on cerebrospinal fluid.^49^

Quantitative measurements of cerebral blood flow in units of ml/100mg/min from Arterial Spin Labeling MRI were available in a subsample of 1154 scans acquired on 450 subjects, and were performed by the University of California, San Francisco, following protocols described in the ADNI website. The Arterial Spin Labeling techniques included 223 3D pseudo-continuous arterial spin labeling (pCASL) and 931 pulsed arterial spin labeling (PASL, 918 2D and 13 3D). We used these measurements in an exploratory analysis to assess the correlation between cerebral blood flow and the corresponding WM-PVS count estimated with our novel algorithm, while controlling for age, sex, race, educational level, body mass index, history of diabetes, cardio-/cerebro-vascular disease, hypertension, dyslipidemia, family history of dementia, the employed arterial spin labeling technique, the total intracranial volume and the volume of the region where cerebral blood flow was assessed.

#### NACC

The NACC database comprises data collected from the Alzheimer’s Disease Centers in the United States funded by the National Institute on Aging. From 2005 to the present, these centers have been contributing data to the Uniform Data Set using a prospective, standardized, and longitudinal clinical evaluation of the enrolled subjects, including both cognitively unimpaired individuals and patients with cognitive impairment and dementia. Participants are enrolled on a referral or volunteer basis and undergo a complete examination yielding demographic data, neuropsychological testing scores, and clinical diagnosis. The haplotype of APOE is run independently by each Alzheimer’s Disease Center and reported in the NACC database. Cerebrospinal fluid samples were collected in a subsample of the participants: Amyloid-β_1-42_, total Tau, and phosphorylated Tau_181_ measurements were available in 363 (including 206 obtained through enzyme-linked immunosorbent assay, ELISA, and 157 through Luminex), 355 (including 193 ELISA and 162 Luminex), and 349 (including 192 ELISA and 157 Luminex) participants. A clinical report of abnormal level of amyloid and tau in cerebrospinal fluid was also available in 96 and 91 participants, respectively. Amyloid PET scans were available in 206 participants (111 AV-45 and 95 FBB) and were visually assessed for amyloid positivity by an experienced physician-scientist. A clinical report of abnormal uptake of amyloid and tau tracers in PET were also available in 432 and 26 participants, respectively. Participants were classified as amyloid-positive based on abnormal amyloid level detected on cerebrospinal fluid (<570 pg/ml for ELISA^53^ or <192 pg/ml for Luminex^54^) or on PET (visually) or as indicated in the clinical report, and as tau-positive based on abnormal level of total tau (>412 pg/ml for ELISA^53^ and >93 pg/ml for Luminex^54^) or phosphorylated tau (>78 pg/ml for ELISA^53^ and >23 pg/ml for Luminex^54^) on cerebrospinal fluid or as indicated in the clinical report.

#### OASIS

OASIS-3 is a retrospective compilation of data collected over the course of 15 years as part of research studies at Washington University in St. Louis.^45^ Participants were recruited from the community via flyers, word of mouth, and community engagements.^45^ Enrolled participants included individuals considered generally healthy or without medical conditions that precluded longitudinal participation or contraindications to study procedures, such as MRI and lumbar puncture.^45^ Participants underwent clinical assessments which comprised collection of medical and family history, physical examination, and neuropsychological evaluation.^45^ No cerebrospinal fluid data were available for the participants included in our analysis. Data on amyloid and tau tracers uptake on PET were available in 73.3% and 31.2% of participants, respectively. Amyloid tracers included AV-45 in 336 cases and PiB in 656 cases, whereas the employed tau tracer was Flortaucipir (18F-AV-1451). Quantitative measurements of the tracer uptake were provided in OASIS-3. The acquisition and processing protocols are fully described in the OASIS-3 documentation (https://www.oasis-brains.org/files/OASIS-3_Imaging_Data_Dictionary_v2.3.pdf). We used the cutoff values recommended in the OASIS-3 documentation to classify participants as amyloid-positive (Centiloid AV-45>20.6 or Centiloid PiB>16.4) and as tau-positive (AV-1451 Tauopathy>1.22).

### Assessment of cognitive status and dementia

In all three cohorts, the Clinical Dementia Rating (CDR) assessment^55^ was performed through standardized interview with the participant and a knowledgeable informant. Six categories of cognitive functioning (memory, orientation, judgment and problem solving, community affairs, home and hobbies, and personal care) were assessed. We used the standard CDR global score cutoff value of 1 to classify participants as demented (1 and above) or non-demented. The Mini-Mental Status Examination^56^ (MMSE) scores were available in a subgroup of 7046 participants and were used only in sensitivity analyses. MMSE evaluates orientation, memory, attention, concentration, naming, repetition, comprehension, and ability to create a sentence and to copy 2 overlapping pentagons.^56^ We did not categorize participants based on the clinical diagnosis when available due to the lack of scientific consensus and the variability in the diagnostic criteria adopted within and across the studies.

### MRI data processing

MRI data were acquired with a variety of 1.5- and 3-Tesla MRI scanners and sequences (Table S1). All T1-weighted images were processed using the *recon-all* module of the freely available FreeSurfer software package (v7.4),^57^ which resampled all the images to 1 mm isotropic resolution and performed an atlas-based brain parcellation. The longitudinal processing scheme was used for estimating brain atrophy (grey and white matter volumes, and cortical thickness) longitudinally.^58^ White matter lesions (WML) were segmented with a previously validated approach on T1-weighted images.^24^ We classified as periventricular WML the clusters of WML adjacent to the lateral ventricles; the remaining clusters of WML were classified as deep WML.

### Robust PVS segmentation method development and validation

The filter developed by Frangi et al.^16^ enhances tubular, vessel-like structures on a gray-scale image and assigns a “vesselness” value to each voxel *ν*(*s*) from eigenvectors *λ* of the Hessian matrix ℋ of the image as:

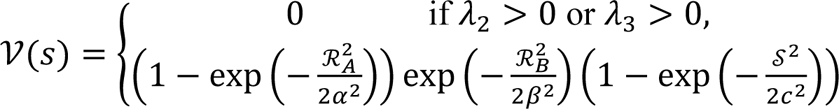

Where, 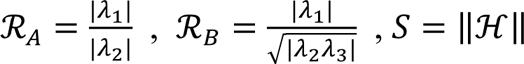.

We and others previously implemented and validated this filter for the segmentation of MRI-visible vascular and perivascular spaces on T1-weighted images using the default, recommended parameters of α=0.5, β=0.5, and c set to half the value of the maximum Hessian norm.^12,13,59^ This approach requires the user to identify a threshold on the vessel map generated by the filter to define the vessel-like structures: values above that threshold (i.e., with high “vesselness” values) are considered vascular and perivascular spaces, and values below are excluded. However, since the scale of the “vesselness” values generated by the filter differs from image to image (Figure S1 A) depending on the signal intensity values of the input image,^16^ and since the signal intensity values on T1-weighted images are represented in arbitrary units which may vary depending on the MRI machine and its calibration, this approach lacks inter-scanner reproducibility^1,9,16–18^ (Fig. S1 B) and is potentially biased even in longitudinal studies. To overcome this issue, we developed and validated a novel approach for the segmentation of MRI-visible vessel-like structures applicable to virtually any type of T1-weighted image.

The MRI data used for the method development and validation were acquired as part of the Biomarkers Consortium for Vascular Contributions to Cognitive Impairment and Dementia (MarkVCID),^60^ the Alzheimer Disease Neuroimaging Initiative (ADNI),^43^ and the Human Connectome Project (HCP development,^61^ young adults,^62^ and aging^63^), and included:

a. the inter-scanner reproducibility dataset: 19 participants who underwent four brain MRI scans within an interval of 3 to 90 days on four different 3-Tesla MRI scanners (General Electric system 750W, Philips Achieva dStream, Siemens Prisma, and Siemens TIM Trio) from MarkVCID;
b. the inter-field-strength reproducibility dataset: 299 MRI sessions from 115 ADNI participants who underwent in each session two brain MRI scans on the same day with a 1.5- and a 3-Tesla scanner;
c. the test-retest dataset: 39 participants from MarkVCID who underwent two brain MRI scans on the same MRI scanner using the same protocol within an interval of 1 to 14 days;
d. the biological validation dataset: 2163 healthy individuals from the HCP whose MRI-visible vascular and perivascular spaces have been studied extensively^21–23^, providing a reference for verifying established associations.

Tables S17 and S18 include the details for the baseline characteristics of the analyzed individuals and the MRI protocols employed in each of the above datasets, respectively. Our novel approach is based on the observation that the total number of voxels with non-zero vesselness value obtained from the Frangi filter applied on T1-weighted images is:

1. consistent across brain images of the same participant acquired with different MRI scanners (inter-scanner and inter-field-strength reproducibility, Figure S2 A and B);
2. consistent across brain images of the same participant acquired on two different MRI sessions with the same MRI scanner and protocol (test-retest repeatability, Figure S2 C);
3. Significantly associated with age, sex, and body mass index (Figure S2 D-F) as previously described for PVS.^21–23^

Therefore, we hypothesized that the value of the voxel corresponding to a single, specific percentile of the total number of voxels with non-zero “vesselness” values could be used as a threshold for consistently and robustly segmenting MRI-visible vessel-like structures across different types of T1-weighted images (Figure S1 C-D). We identified this percentile to be 85%, based on the average ratio between the number of voxels that we previously segmented as vascular and perivascular spaces in the HCP dataset^21,22^ and the corresponding total number of voxels with non-zero vesselness value.

In each individual vesselness map generated by the Frangi filter, the vesselness value corresponding to the 85^th^ percentile of the total number of non-zero voxels was automatically computed: the voxels with vesselness value above this threshold were retained and binarized to make the PVS mask, whereas those below or equal to the threshold were excluded. To improve the specificity of the PVS segmentation,^17,59^ the Frangi filter was applied on FreeSurfer’s white matter mask with the following modifications: the voxels labeled as corpus callosum by FreeSurfer were excluded; periventricular areas were excluded by subtracting FreeSurfer’s lateral ventricle binary mask enlarged by 3 units from the white matter binary mask; white matter lesions (WML) segmented with SAMSEG^24^ were also excluded. Finally, we used MATLAB’s *regionprops3* function with the default 26-connected neighborhood definition to compute PVS count and mean diameter across all PVS clusters with in-plane size of at least 2 voxels detected in the modified white matter mask of each subject.

Accuracy of the PVS segmentation was assessed via visual inspection and quantified with the Dice similarity coefficient using as a reference the PVS masks obtained with an established and previously validated technique applied on the HCP dataset (Fig. S3A).^12,13,17,23^ The Dice similarity coefficient ranges from 0, indicating no spatial overlap between two sets of binary segmentation masks, to 1, indicating complete overlap.

The robustness of the PVS metrics assessed in our study (i.e., PVS count and PVS mean diameter) across different MRI scanners (inter-scanner and inter-field-strength reproducibility) and sessions (test-retest repeatability) was assessed with the intraclass correlation coefficients, ranging from 0 to 1, where a higher value indicates higher agreement between the compared modalities (Fig. S4). Similar evaluations were also performed for WML metrics (Fig. S6)

Since typically FLAIR and T2-weighted images are considered more sensitive to WML and PVS, respectively,^8,9^ due to a higher contrast between the cerebral parenchyma and the WML/PVS, we also evaluated the suitability of assessing WML and PVS with only the T1-weighted images in two ways: 1) we measured the correlation between the number of WML/PVS voxels measured on T1-weighted versus FLAIR/T2-weighted images, to determine the strength and direction of their relationship (Fig. S5); 2) to confirm the spatial agreement of the WML/PVS voxels across the two modalities, we measured the overlap of the WML/PVS voxels identified on T1-weighted images with the WML/PVS voxels (and the adjacent 3 voxels to account for any residual misalignment between modalities) identified from FLAIR/T2-weighted images rigidly registered to the corresponding T1-weighted images.

### Statistical analysis

All the models and simulations described below were adjusted for intracranial volume and the following baseline factors (as reported on the documented clinical assessment and subject health history): age, sex, educational level, race, body mass index, CDR global score, family history of dementia (positive if any of the participants’ parents were reported to have dementia), and history of hypertension, dyslipidemia, diabetes, and cardio-/cerebro-vascular disease (i.e., any of the following: heart failure, angina, cardiac arrest, stent placement, coronary artery bypass, pacemaker, defibrillator, heart valve replacement or repair, stroke, transient ischemic attack). These factors were all available for more than 97% participants. We used missing indicators for handling missing values in the covariates.

The associations of our PVS markers with cognitive status (non-dementia versus dementia) and the other covariates at the baseline visit were assessed with general linear models stratified according to study. The association of our PVS markers at the baseline with subsequent risk of developing dementia was assessed in each study independently with Cox proportional-hazards models and the results were integrated using a random-effects meta-analysis to account for potential differences among the studies (two-stage pooled analysis). Inter-study heterogeneity was assessed statistically with the I^2^. Person-time was calculated from the baseline clinical visit until the visit where the dementia was documented or the last clinical visit, whichever occurred first. The proportional-hazard assumptions were verified by assessing the relationship between Schoenfeld residuals and time.^64^ Additionally, we analyzed these associations by combining in the same model all the individual-level data (simple pooled analysis), stratified according to study and adjusted for the same covariates, and used penalized splines to assess their deviation from linearity.^65^ In addition to the covariates used in all the other models, the linear models and Cox-regression models above were also controlled for the time interval between the MRI scan and the clinical visit of the cognitive assessment at the baseline.

Linear mixed-effects models with random intercepts and slopes for each participant were used to estimate the longitudinal trajectories of grey matter volume, cortical thickness, and white matter volume according to baseline PVS count or mean diameter. In addition to the covariates used in all the other models, the linear mixed-effects models were also adjusted for the value of the dependent variable at the baseline, and the following characteristics of the MRI scanner that may influence the longitudinal estimation of brain atrophy: field strength, manufacturer, and intra-individual consistency of the protocol used for the longitudinal MRI acquisitions (consistent versus non-consistent protocol). Interaction terms between time and all predictors were included as well to estimate the marginal per-year effects of each predictor. Linear mixed-effects models with random intercepts and slopes for each participant were also used to estimate and compare the longitudinal trajectories of PVS and WML markers in non-demented individuals who converted to dementia versus non-converters.

Sensitivity analyses for all the models above included: 1) the individual assessment of other potential confounding factors available only in a subsample of the participants (92.6% history of tobacco smoking, 90.0% Apolipoprotein E alleles, 39.0% amyloid-β and 24.7% tau status as assessed on cerebrospinal fluid and/or Positron-Emission Tomography); 2) the use of MMSE rather than CDR for the cognitive evaluation (available in 70.3% of the participants); 3) the assessment of MRI acquisition artifacts or factors that could have influenced the estimation of the vascular markers. In sensitivity analyses, hazard ratios were estimated from Cox models with stratification according to study cohort (simple pooled analysis) owing to smaller sample sizes in the individual studies.

To evaluate the potential utility of PVS markers as a screening tool in clinical trials, we computed the sample size needed to detect improvements in cognitive decline trajectories when restricting the sample based on the baseline values of the marker, and compared it to the sample size requested without any restriction. The clinical trials were simulated with 1:1 allocation of active treatment and placebo, assuming a 30% treatment effect on cognition over time, a trial duration of 48 months, and cognitive testing every 12 months. 500 simulations were generated with bootstrap iteration.

All the analyses were based on *a priori* hypotheses, but to account for two variables of interest (PVS count and mean diameter), we present P values that were corrected for multiple comparisons with the use of the Holm–Bonferroni procedure.^66^ In brain regional analyses, the correction for multiple comparisons was performed across all the analyzed regions. All statistical analyses were performed in R v4.3.3. The following R packages were used for the statistical analyses and generation of the plots: *survival* was used for fitting the Cox proportional-hazards models;^67^ *meta* was used for the meta-analysis;^68^ *lmer4* was used for fitting the non-linear mixed models;^69^ *ggeffects* was used for estimating the marginal effects in the non-linear mixed models;^70^ *longpower* was used for the simulation of clinical trials;^71^ *ggseg* was used for generating the plots with the brain statistics;^72^ *ggplot2* was used for generating all the other plots.^73^

## Data Availability

All data produced in the present study are available upon reasonable request to the authors

## Acknowledgements

The authors thank all the participants and personnel involved in the data collection.

The authors G.B. and M.H. are supported by a grant (U54CA261717 to Dr. Hayden-Gephart) from the National Institutes of Health (NIH), J.C. is supported by grants RF1MH123223, R01AG070825, and R01NS128486 from the NIH.

The authors thank Dr. Michael Greicius, M.D., from the Department of Neurology at Stanford University for valuable discussion of the manuscript.

The image computing resources provided by the Laboratory of Neuro Imaging Resource (LONIR) at the University of Southern California are supported in part by NIH National Institute of Biomedical Imaging and Bioengineering (NIBIB) grant P41EB015922.

Data used in preparation of this article include data obtained from the MarkVCID consortium. A complete listing of MarkVCID investigators can be found at: www.markvcid.org.

## ADNI

Data collection and sharing for this project was funded by the Alzheimer’s Disease Neuroimaging Initiative (ADNI) (National Institutes of Health Grant U01 AG024904) and DOD ADNI (Department of Defense award number W81XWH-12-2-0012). ADNI is funded by the National Institute on Aging, the National Institute of Biomedical Imaging and Bioengineering, and through generous contributions from the following: AbbVie, Alzheimer’s Association; Alzheimer’s Drug Discovery Foundation; Araclon Biotech; BioClinica, Inc.; Biogen; Bristol-Myers Squibb Company; CereSpir, Inc.; Cogstate; Eisai Inc.; Elan Pharmaceuticals, Inc.; Eli Lilly and Company; EuroImmun; F. Hoffmann-La Roche Ltd and its affiliated company Genentech, Inc.; Fujirebio; GE Healthcare; IXICO Ltd.; Janssen Alzheimer Immunotherapy Research & Development, LLC.; Johnson & Johnson Pharmaceutical Research & Development LLC.; Lumosity; Lundbeck; Merck & Co., Inc.; Meso Scale Diagnostics, LLC.; NeuroRx Research; Neurotrack Technologies; Novartis Pharmaceuticals Corporation; Pfizer Inc.; Piramal Imaging; Servier; Takeda Pharmaceutical Company; and Transition Therapeutics. The Canadian Institutes of Health Research is providing funds to support ADNI clinical sites in Canada. Private sector contributions are facilitated by the Foundation for the National Institutes of Health (www.fnih.org). The grantee organization is the Northern California Institute for Research and Education, and the study is coordinated by the Alzheimer’s Therapeutic Research Institute at the University of Southern California. ADNI data are disseminated by the Laboratory for Neuro Imaging at the University of Southern California.

## NACC

The National Alzheimer’s Coordinating Center (NACC) database is funded by NIA/NIH Grant U24 AG072122. NACC data are contributed by the NIA-funded ADRCs: P30 AG062429 (PI James Brewer, MD, PhD), P30 AG066468 (PI Oscar Lopez, MD), P30 AG062421 (PI Bradley Hyman, MD, PhD), P30 AG066509 (PI Thomas Grabowski, MD), P30 AG066514 (PI Mary Sano, PhD), P30 AG066530 (PI Helena Chui, MD), P30 AG066507 (PI Marilyn Albert, PhD), P30 AG066444 (PI John Morris, MD), P30 AG066518 (PI Jeffrey Kaye, MD), P30 AG066512 (PI Thomas Wisniewski, MD), P30 AG066462 (PI Scott Small, MD), P30 AG072979 (PI David Wolk, MD), P30 AG072972 (PI Charles DeCarli, MD), P30 AG072976 (PI Andrew Saykin, PsyD), P30 AG072975 (PI David Bennett, MD), P30 AG072978 (PI Neil Kowall, MD), P30 AG072977 (PI Robert Vassar, PhD), P30 AG066519 (PI Frank LaFerla, PhD), P30 AG062677 (PI Ronald Petersen, MD, PhD), P30 AG079280 (PI Eric Reiman, MD), P30 AG062422 (PI Gil Rabinovici, MD), P30 AG066511 (PI Allan Levey, MD, PhD), P30 AG072946 (PI Linda Van Eldik, PhD), P30 AG062715 (PI Sanjay Asthana, MD, FRCP), P30 AG072973 (PI Russell Swerdlow, MD), P30 AG066506 (PI Todd Golde, MD, PhD), P30 AG066508 (PI Stephen Strittmatter, MD, PhD), P30 AG066515 (PI Victor Henderson, MD, MS), P30 AG072947 (PI Suzanne Craft, PhD), P30 AG072931 (PI Henry Paulson, MD, PhD), P30 AG066546 (PI Sudha Seshadri, MD), P20 AG068024 (PI Erik Roberson, MD, PhD), P20 AG068053 (PI Justin Miller, PhD), P20 AG068077 (PI Gary Rosenberg, MD), P20 AG068082 (PI Angela Jefferson, PhD), P30 AG072958 (PI Heather Whitson, MD), P30 AG072959 (PI James Leverenz, MD).

## OASIS

Data were provided in part by the Open Access Series of Imaging Studies (OASIS) OASIS-3: Longitudinal Multimodal Neuroimaging: Principal Investigators: T. Benzinger, D. Marcus, J. Morris; NIH P30 AG066444, P50 AG00561, P30 NS09857781, P01 AG026276, P01 AG003991, R01 AG043434, UL1 TR000448, R01 EB009352, NIH P30 AG066444, AW00006993.

AV-45 doses were provided by Avid Radiopharmaceuticals, a wholly owned subsidiary of Eli Lilly.

AV-1451 doses were provided by Avid Radiopharmaceuticals, a wholly owned subsidiary of Eli Lilly.

## MarkVCID

MarkVCID was launched in 2016 by the NIH’s National Institute of Neurological Disorders and Stroke (NINDS) and National Institute on Aging (NIA), and consists of research groups across the United States. The primary goal of MarkVCID is to generate a suite of validated biomarkers ready for application to clinical trials aimed at identifying disease-modifying therapies for VCID. For up-to-date information, see www.markvcid.org. Data collection and sharing was funded by NINDS/NIA as part of the Biomarkers Consortium for Vascular Contributions to Cognitive Impairment and Dementia (MarkVCID): U24NS100591, UH2NS100599, UH2NS100605, UH2NS100588, UH2NS100608, UH2NS100606, UH2NS100598, UH2NS100614.

## Human Connectome Project

Data were provided in part by the Human Connectome Project (HCP), WU-Minn Consortium (1U54MH091657) funded by the 16 NIH Institutes and Centers that support the NIH Blueprint for Neuroscience Research; and by the McDonnell Center for Systems Neuroscience at Washington University. Data from HCP-Aging cohort was supported by the National Institute on Aging of the National Institutes of Health under Award Number U01AG052564. Data from HCP-Development cohort was supported by the National Institute of Mental Health of the National Institutes of Health under Award Number U01MH109589. The content is solely the responsibility of the authors and does not necessarily represent the official views of the National Institutes of Health. MRI and clinical data can be accessed from https://www.humanconnectome.org.

## Author contributions

Giuseppe Barisano, Jeiran Choupan and Melanie Hayden-Gephart designed the study. Giuseppe Barisano developed the method for robust perivascular space segmentation. Giuseppe Barisano processed the MRI data, gathered the demographic and clinical data, and performed the statistical analysis. Giuseppe Barisano, Jeiran Choupan and Melanie Hayden-Gephart vouch for the data and the analysis. Giuseppe Barisano developed the interactive website. Giuseppe Barisano wrote the first draft of the manuscript. All the authors contributed to result interpretation and manuscript editing. All the authors agreed to publish the paper.

Data used in preparation of this article were obtained from the Alzheimer’s Disease Neuroimaging Initiative (ADNI) database (adni.loni.usc.edu). As such, the investigators within the ADNI contributed to the design and implementation of ADNI and/or provided data but did not participate in analysis or writing of this report.

