## Supplementary Figures for "Cerebral perivascular spaces as predictors of dementia risk and accelerated brain atrophy"

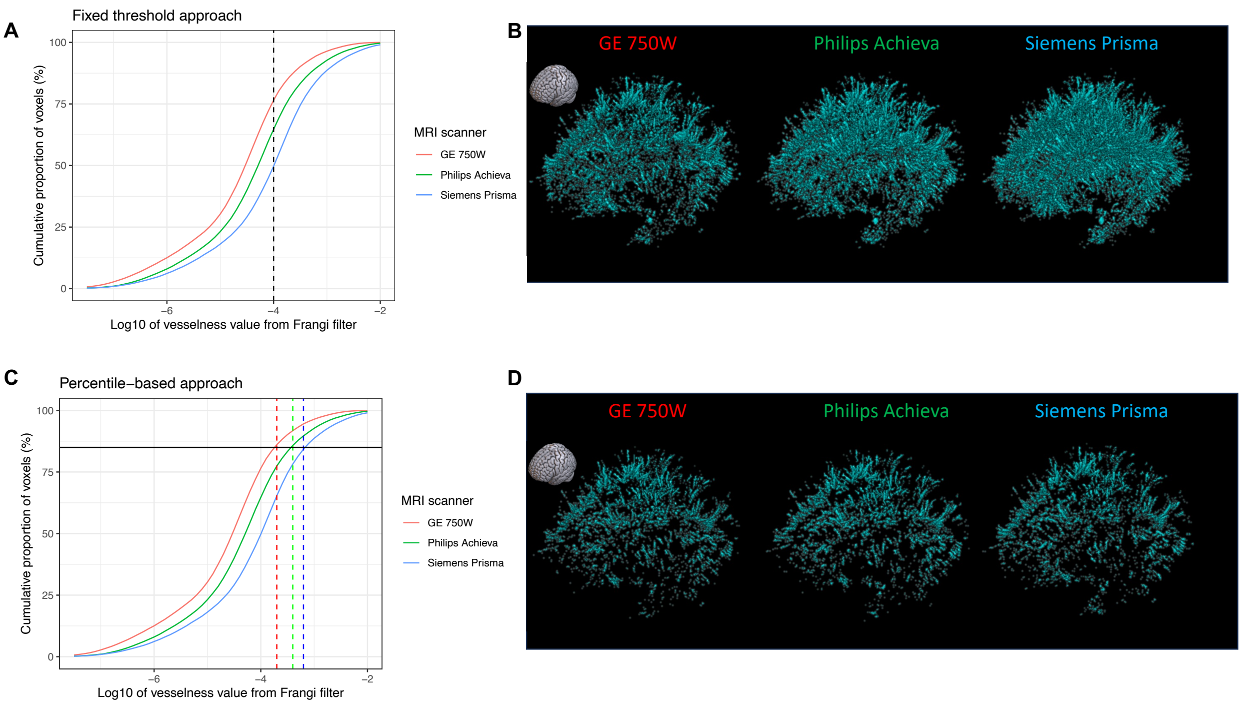


**Figure S1. Cumulative distributions of the voxel values from three vesselness maps generated with the Frangi filter applied on three different T1-weighted brain images acquired on the same participant with three different MRI scanners.** The original approach requires to set a single threshold (black vertical dashed line in panel A, in this case equal to $10^{-4}$), and to use this threshold to segment vessel-like structures (Panel A). However, this approach lacks inter-scanner reproducibility, because the scale of the vesselness maps depends on the signal intensity of the input image, which may differ among MRI scanners and protocols. In fact, in this example the threshold would lead to very different number of segmented voxels: vessel-like masks from Siemens scanner will have approximately 12.5 and 25% more voxels than those from Philips and GE scanners, respectively (Panel B). In our novel approach, we set specific thresholds to the individual images (vertical dashed lines, red for the GE-derived image, green for the Philips-derived image, and blue for the Siemens-derived image) based on the value of the voxel corresponding to the $85^{th}$ percentile (black horizontal solid line) of the total number of non-zero voxels of the vesselness map (Panel C). This approach leads to consistent PVS masks (Panel D) and robust metrics derived from the PVS masks (Figures S2 A-C and S4), while preserving inter-individual differences and accuracy (Figure S2 D-F and S3).
Panels B and D report the 3D representations of the PVS segmentation masks of the same participant obtained with 3 different MRI scanners with fixed threshold approach (panel B) and our novel percentile-based approach (panel D). The small brain icon on the top left of each panel indicates the orientation of the PVS masks.

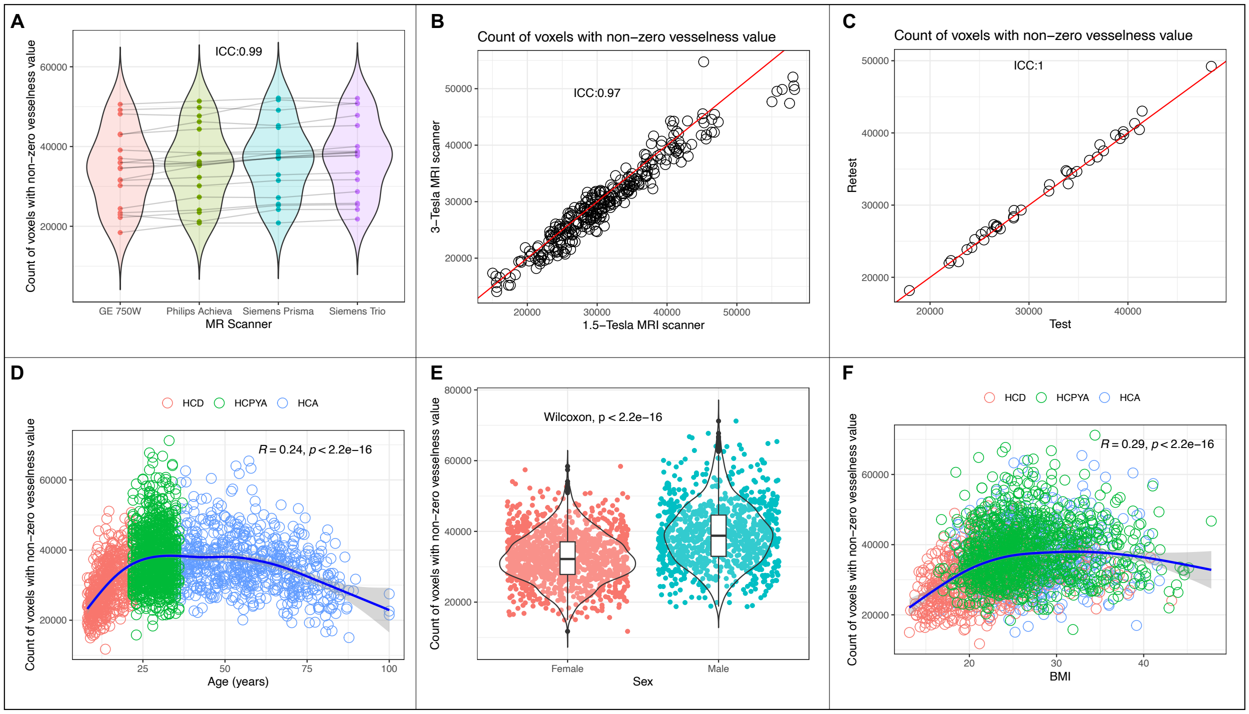


**Figure S2. Reproducibility and associations of the total number of non-zero voxels in the vesselness maps generated with the Frangi filter.**

The total number of non-zero voxels in the vesselness maps generated with the Frangi filter shows excellent inter-scanner (panel A) and inter-field-strength (panel B) reproducibility and test-retest repeatability (panel C). The total number of non-zero voxels in the vesselness maps generated with the Frangi filter is significantly associated with age (panel D), sex (panel E), and body mass index (BMI, panel F), and these associations are consistent with what previously published on MRI-visible vascular and perivascular spaces.[21–23] Therefore, it is possible to use a percentile-based approach for segmenting MRI-visible vessel-like structures (Figure S1 B), as the inter-individual differences are preserved. In panel A, data are from 19 subjects who underwent four brain MRI scans within an interval of 3 to 90 days on four different 3-Tesla MRI scanners (General Electric system 750W, Philips Achieva dStream, Siemens Prisma, and Siemens TIM Trio) from MarkVCID dataset. In panel B, data are from 299 MRI sessions from 115 ADNI participants who underwent in each session two brain MRI scans on the same day with a 1.5- and a 3-Tesla scanner. In panel C, data are from 39 participants (MarkVCID) who underwent two brain MRI scans on the same MRI scanner using the same protocol within an interval of 1 to 14 days. In panels D-F, data are from 2163 subjects from the Human Connectome Project Aging (HCA), Development (HCD) and Young Adults (HCPYA) datasets. See also Table S17 and S18 for their demographic and radiographic characteristics, respectively. ICC: Intraclass correlation coefficient. R in panels B and F is the Spearman's rho. The red lines in panels B and C are the identity lines. The blue lines in panels D and F have been generated with locally estimated smoothing.

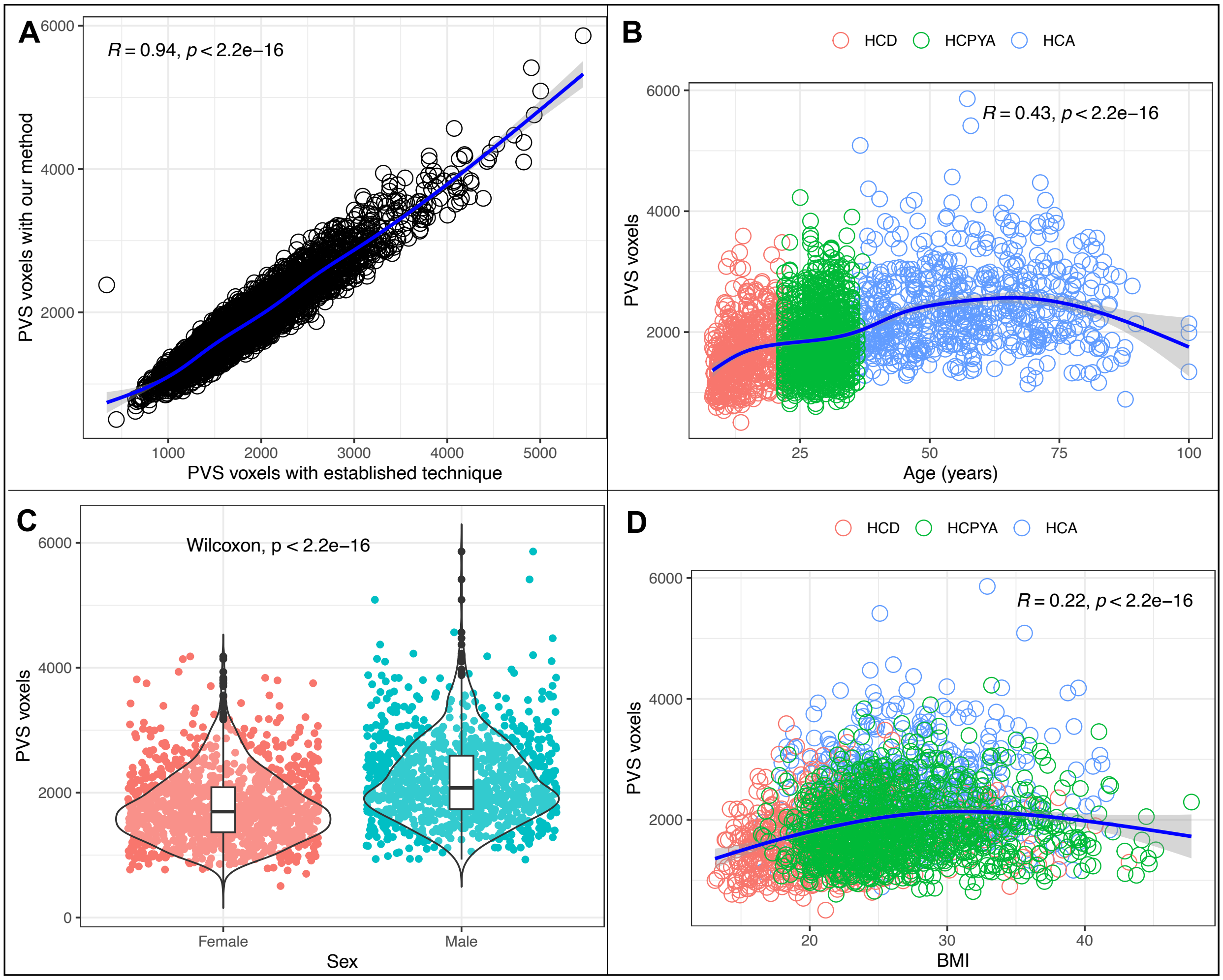


**Figure S3. Reliability of PVS segmentation.**

There was a strong correlation (panel A) between the numbers of PVS voxels identified with our fully automated technique (y-axis) and those obtained with the established and previously validated method[12,21–23] (x-axis). The average Dice similarity coefficient was 0.95. Consistently, PVS measured with our method also showed a significant positive association with age (panel B), male sex (panel C), and body mass index (panel D), as previously published with the validated semi-automated techniques in the Human Connectome Project datasets[21–23]. R in panels B and D is the Spearman's rho. Data are from 2163 subjects from the Human Connectome Project Aging (HCA), Development (HCD) and Young Adults (HCPYA) datasets. The blue lines in panels B and D have been generated with locally estimated smoothing.

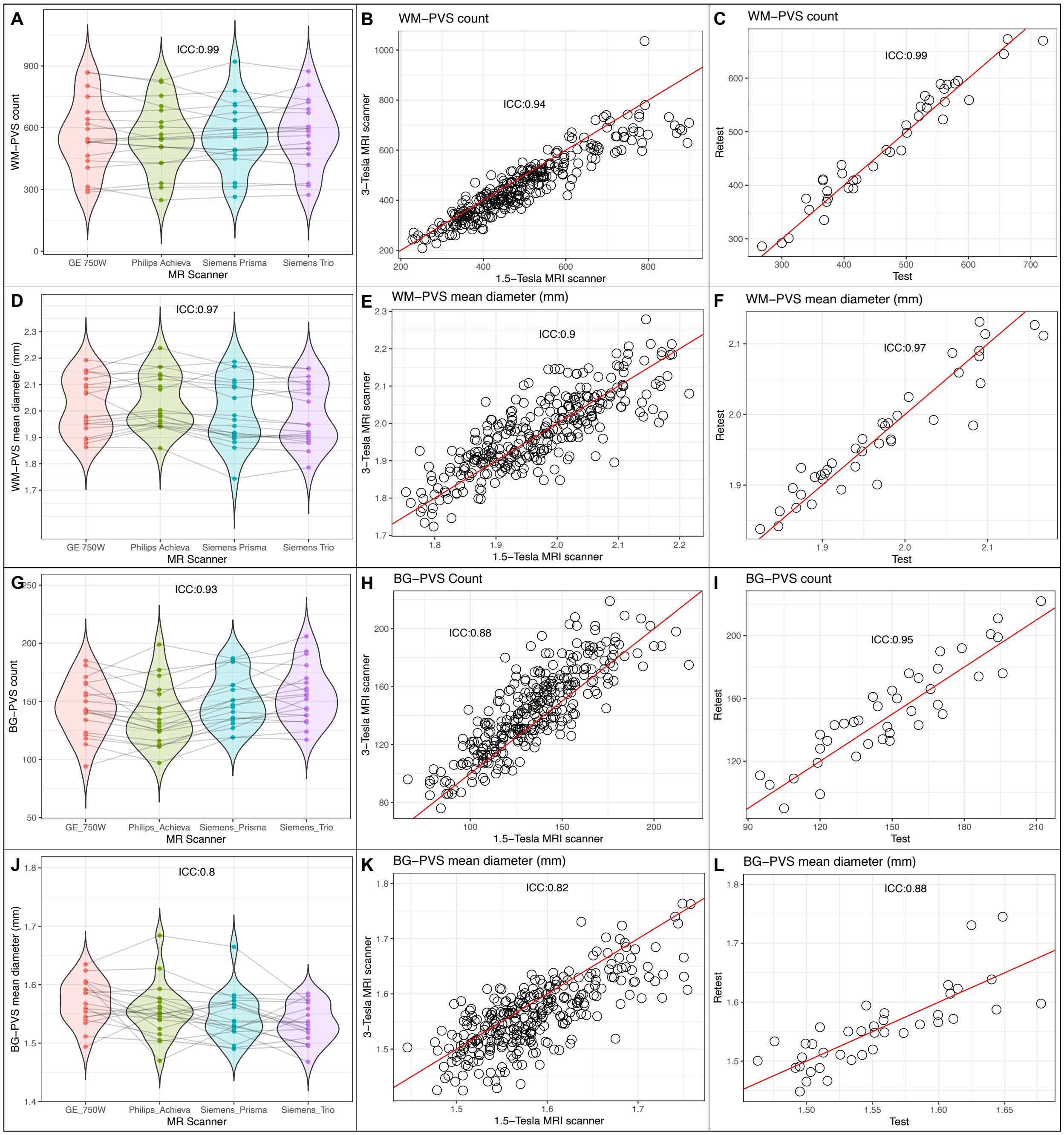


**Figure S4. Reproducibility of PVS count and diameter computed on the binary MRI-visible vascular and perivascular spaces masks obtained with our novel approach.**

The PVS count (panels A-C for WM-PVS and G-I for BG-PVS) and PVS mean diameter (panels D-F for WM-PVS and J-L for BG-PVS) computed on the binary PVS masks obtained with our novel approach show excellent inter-scanner (panels A and D for WM-PVS and panels G and J for BG-PVS, respectively) and inter-field-strength (panels B and E for WM-PVS and panels H and K for BG-PVS, respectively) reproducibility and test-retest repeatability (panels C and F for WM-PVS and panels I and L for BG-PVS, respectively). In panels A, D, G and J data are from 19 subjects who underwent four brain MRI scans within an interval of 3 to 90 days on four different 3-Tesla MRI scanners (General Electric system 750W, Philips Achieva dStream, Siemens Prisma, and Siemens TIM Trio) from MarkVCID. In panels B, E, H and K data are from 299 MRI sessions from 115 ADNI participants who underwent in each session two brain MRI scans on the same day with a 1.5- and a 3-Tesla scanner. In panels C, F, I and L data are from 39 participants (MarkVCID) who underwent two brain MRI scans on the same MRI scanner using the same protocol within an interval of 1 to 14 days. See also Table S17 and S18 for their demographic and radiographic characteristics, respectively. The red lines in panels B, C, E, F, H, I, K and L are the identity lines. ICC: Intraclass correlation coefficient.

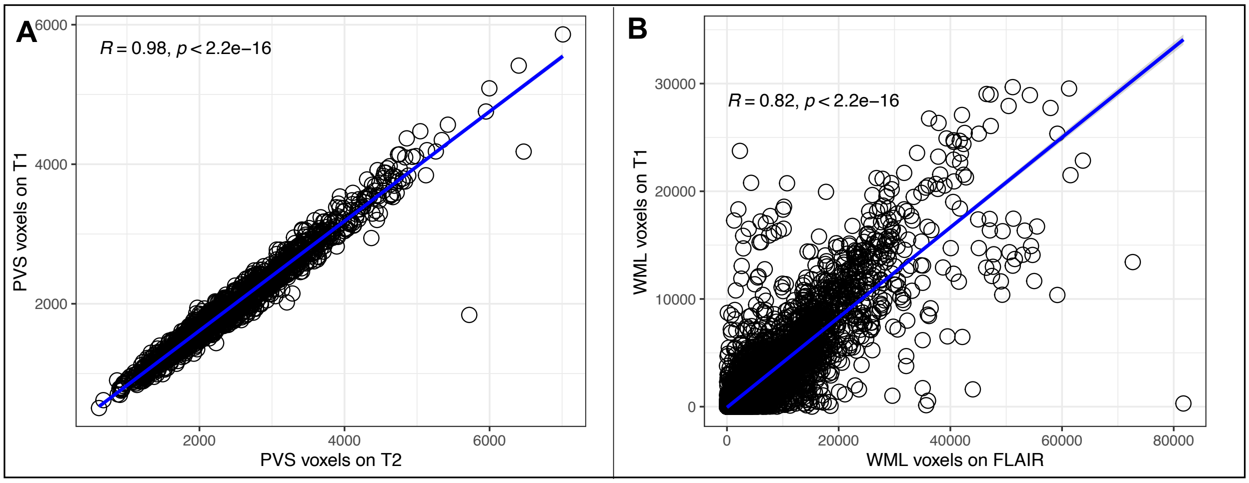


**Figure S5. Suitability of T1-weighted images for the assessment of PVS and WML.** The number of PVS voxels identified by our novel algorithm (panel A) and of WML voxels identified with an established technique[24] (panel B), both applied on T1-weighted images (y-axis), are strongly correlated with those identified by the same algorithms applied on sequences considered more sensitive for PVS (T2-weighted images) and WML (FLAIR). R in panels A and B is the Spearman's rho. Data are from 1341 subjects in the Human Connectome Project dataset (panel A) and 6707 MR sessions of 1889 subjects in ADNI (panel B).

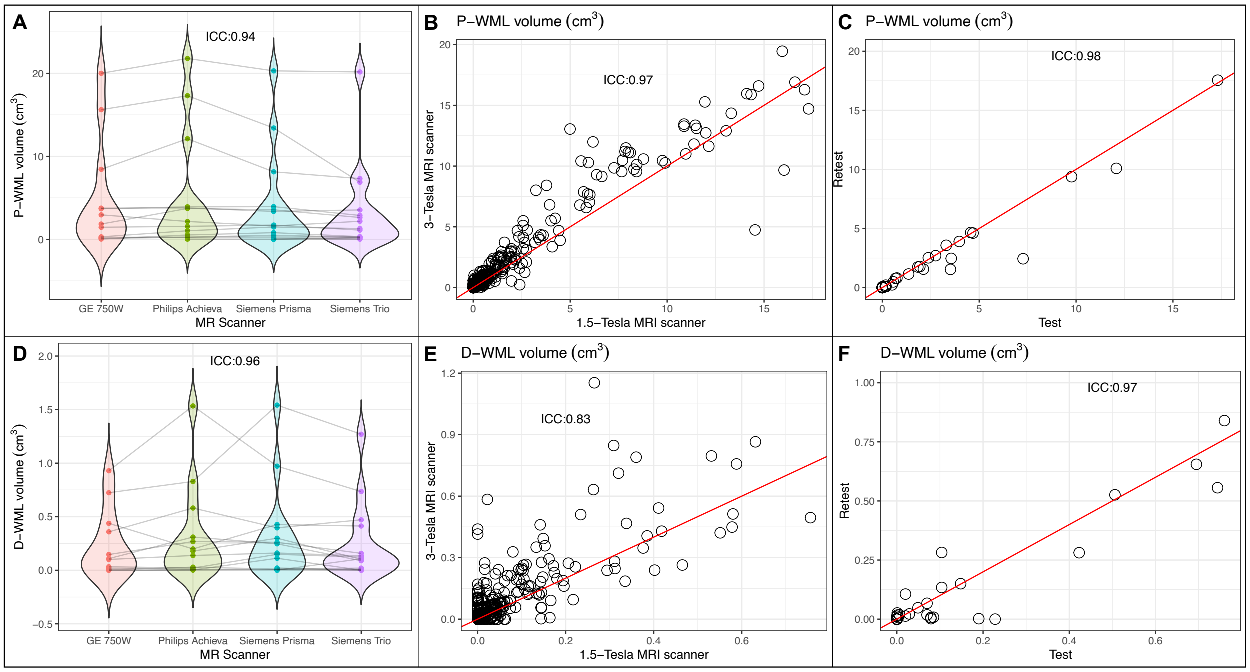


**Figure S6. Reproducibility of WML volume computed with a validated technique applied on T1-weighted images.**

The WML volumes (panels A-C for periventricular WML, P-WML, and D-F for deep WML, D-WML) computed with a validated approach[24] show excellent inter-scanner (panels A and D for P-WML and D-WML, respectively) and inter-field-strength (panels B and E for P-WML and D-WML, respectively) reproducibility and test-retest repeatability (panels C and F for P-WML and D-WML, respectively). In panels A and D data are from 19 subjects who underwent four brain MRI scans within an interval of 3 to 90 days on four different 3-Tesla MRI scanners (General Electric system 750W, Philips Achieva dStream, Siemens Prisma, and Siemens TIM Trio) from MarkVCID dataset. In panels B and E data are from 308 MRI sessions from 116 ADNI participants who underwent in each session two brain MRI scans on the same day with a 1.5- and a 3-Tesla scanner. In panels C, F, I and L data are from 39 participants (MarkVCID) who underwent two brain MRI scans on the same MRI scanner using the same protocol within an interval of 1 to 14 days. See also Table S17 and S18 for their demographic and radiographic characteristics, respectively. The red lines in panels B, C, E and F are the identity lines. ICC: Intraclass correlation coefficient.

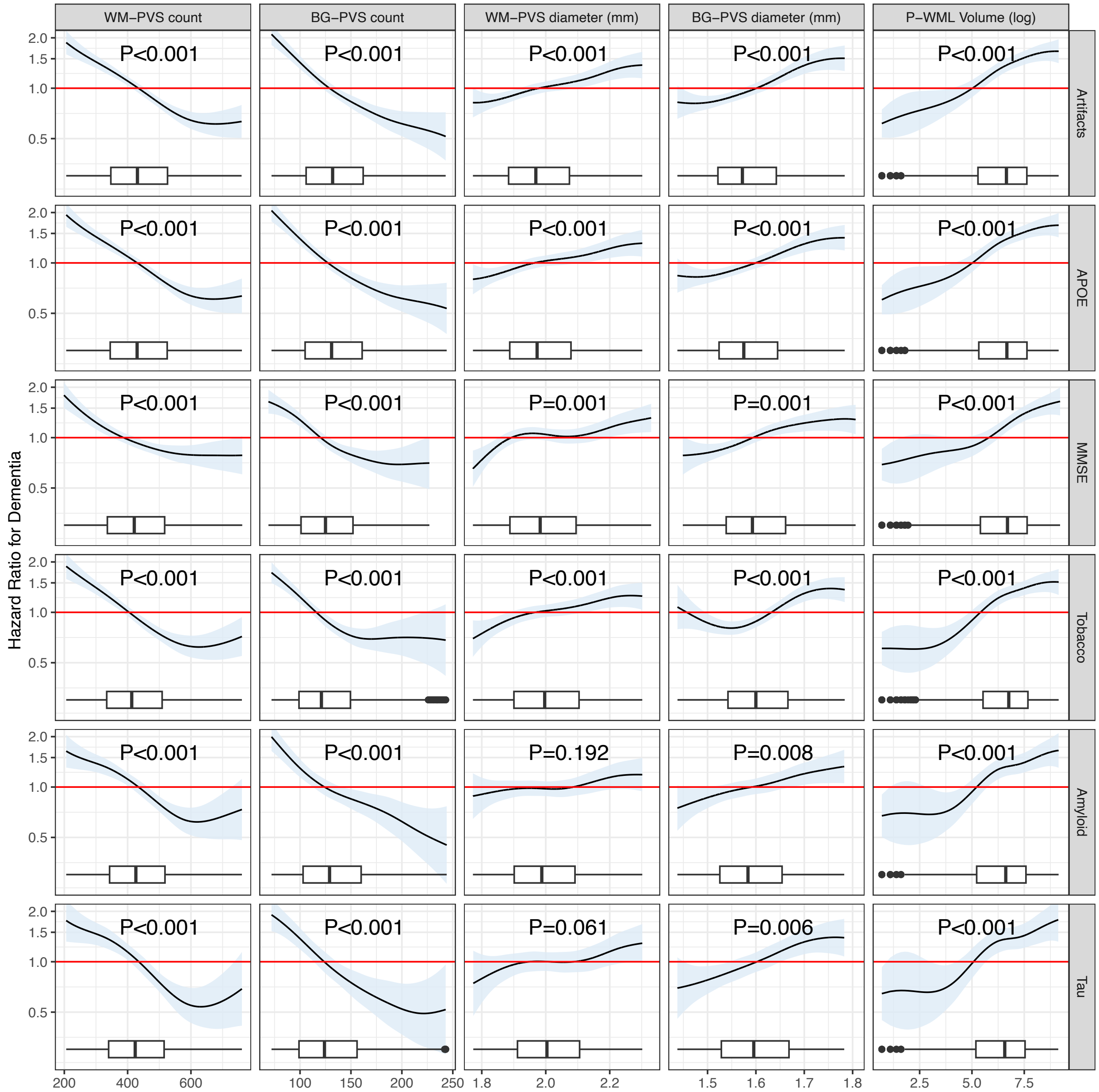


**Figure S7. Spline plots for the association of vascular markers with dementia.** Hazard ratios were estimated from fully adjusted models with the same covariates as in the primary analysis (Fig. 1). For “Artifacts”, MRI data with artifacts that could have affected the estimate of the vascular marker or acquired after the injection of a contrast agent were excluded from the model. For “MMSE” plots, the definition of dementia was based on MMSE score (dementia if MMSE equal to 24 or below) and the covariate “CDR global score at the baseline” has been replaced with “MMSE score at the baseline”. In all the other plots, the indicated variable was included as additional covariate in the model. “APOE” denotes the covariate for Apolipoprotein-E allele genotype, “MMSE” denotes the score from the Mini-Mental State Examination, “Tobacco” denotes history of tobacco smoking (positive or negative), “Amyloid” indicates the status for amyloid-beta pathology as assessed on cerebrospinal fluid and/or positron-emission tomography (positive or negative), and “Tau” indicates the status for tau pathology as assessed on cerebrospinal fluid and/or positron-emission tomography (positive or negative). Shaded areas indicate 95% confidence intervals, and the red line at 1.0 indicates the reference. The boxplots depict the distributions of the marker, with lower and upper hinges corresponding to the first and third quartiles. The whiskers extend from the hinge to values no further than 1.5 times the interquartile range from the hinge. Dots indicate values that are farther than 1.5 times the interquartile range. Limits of the plots are from 2.5th to 97.5th percentile of the distribution. Estimated hazard ratios for these models are included in Table S7.

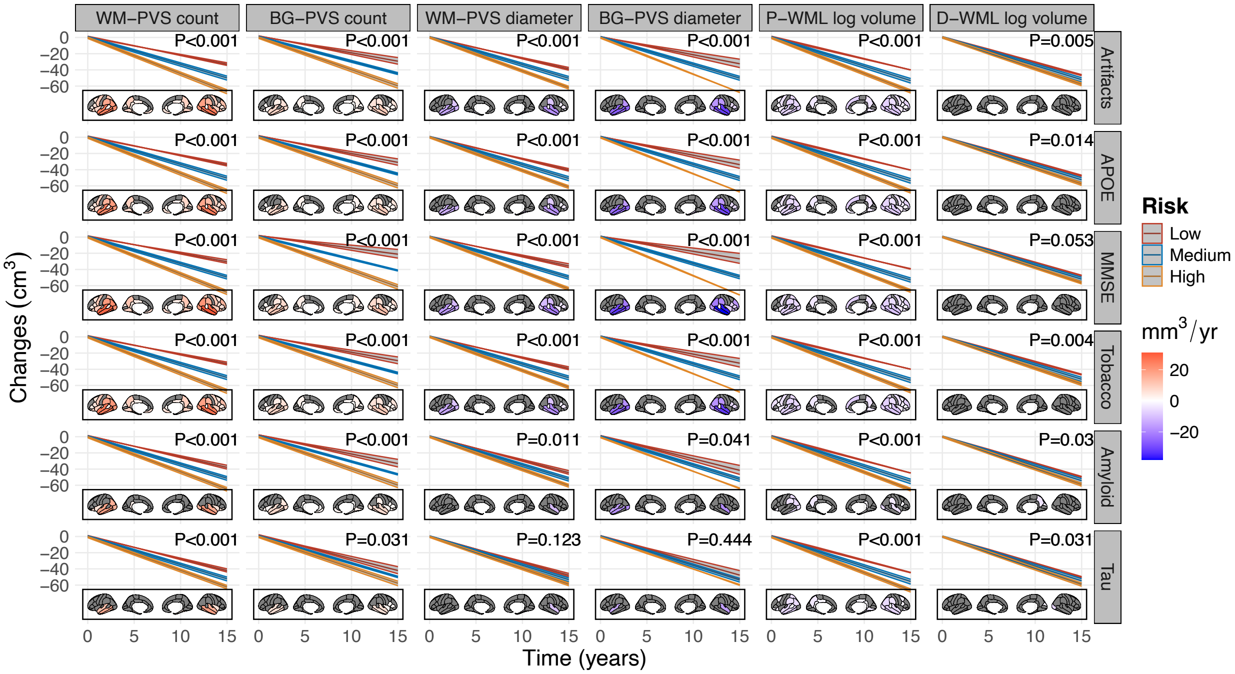


**Figure S8. Sensitivity analyses for the estimated trajectories of grey matter atrophy in relation to PVS and WML markers.**

The effects of PVS and WML markers on the trajectories of total grey matter volume were estimated from fully adjusted linear mixed-effects models with random intercepts and slopes for each individual participant and controlling for the same covariates as in the primary analysis (Fig. 2). For "Artifacts", cases where the estimate of the vascular marker may have been affected by MRI artifacts were excluded from the model. For "MMSE" plots, the covariate "CDR global score at the baseline" has been replaced with "MMSE score at the baseline". In all the other plots, the indicated variable was included as additional covariate in the model. "APOE" denotes the covariate for Apolipoprotein-E allele genotype, "MMSE" denotes the score from the Mini-Mental State Examination, "Tobacco" denotes history of tobacco smoking (positive or negative), "Amyloid" indicates the status for amyloid-beta pathology as assessed on cerebrospinal fluid and/or positron-emission tomography (positive or negative), and "Tau" indicates the status for tau pathology as assessed on cerebrospinal fluid and/or positron-emission tomography (positive or negative). Shaded areas indicate 95% confidence intervals.

Three equally spaced values from the low-risk (red), medium-risk (blue), and high-risk (yellow) tertile (Table S16) of each vascular marker are shown. See also Table S13 for coefficients of sensitivity analysis.

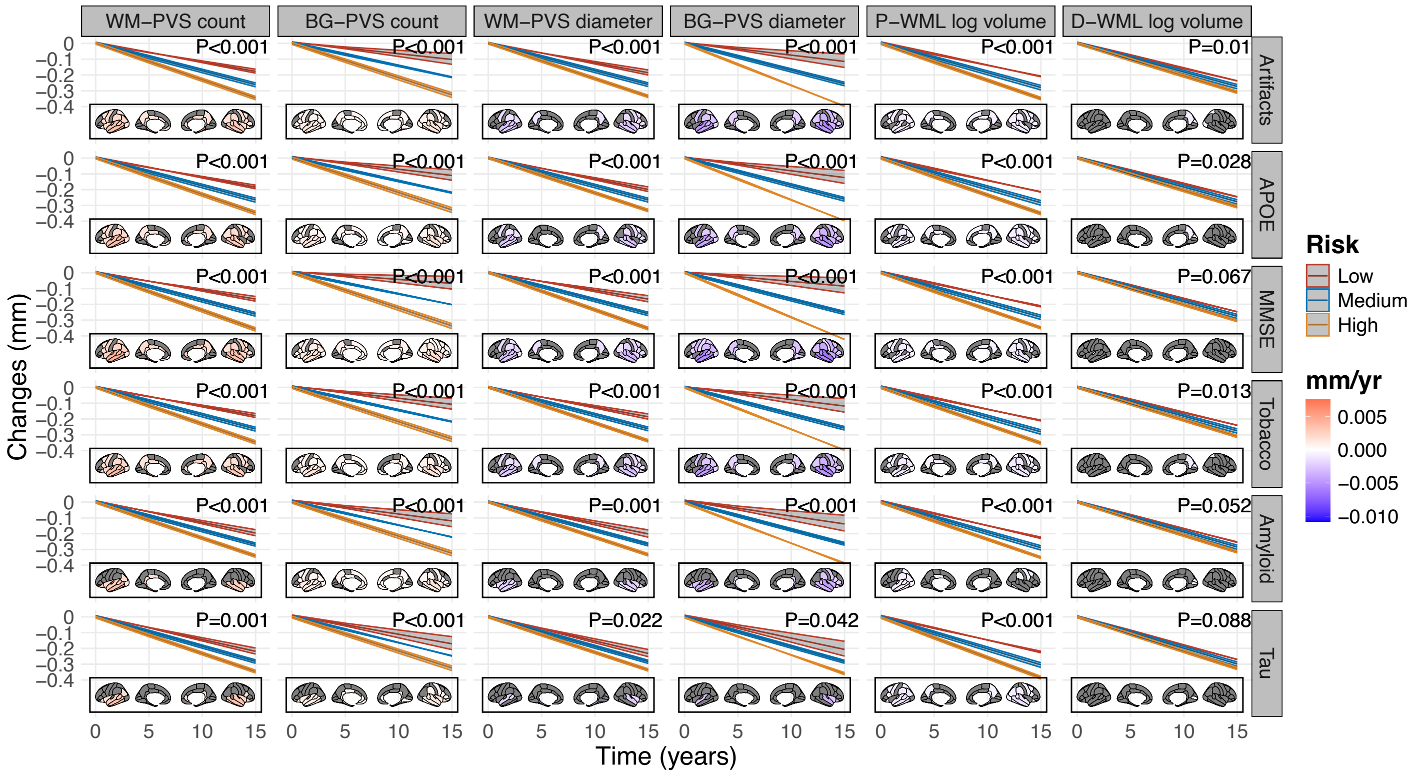


**Figure S9. Sensitivity analyses for the estimated trajectories of cortical thickness atrophy in relation to PVS and WML markers.**

The effects of PVS and WML markers on the trajectories of cortical thickness were estimated from fully adjusted linear mixed-effects models with random intercepts and slopes for each individual participant and controlling for the same covariates as in the primary analysis (Fig. 2). For “Artifacts”, cases where the estimate of the vascular marker may have been affected by MRI artifacts were excluded from the model. For “MMSE” plots, the covariate “CDR global score at the baseline” has been replaced with “MMSE score at the baseline”. In all the other plots, the indicated variable was included as additional covariate in the model. “APOE” denotes the covariate for Apolipoprotein-E allele genotype, “MMSE” denotes the score from the Mini-Mental State Examination, “Tobacco” denotes history of tobacco smoking (positive or negative), “Amyloid” indicates the status for amyloid-beta pathology as assessed on cerebrospinal fluid and/or positron-emission tomography (positive or negative), and “Tau” indicates the status for tau pathology as assessed on cerebrospinal fluid and/or positron-emission tomography (positive or negative). Shaded areas indicate 95% confidence intervals. Three equally spaced values from the low-risk (red), medium-risk (blue), and high-risk (yellow) tertile (Table S16) of each vascular marker are shown. See also Table S13 for coefficients of sensitivity analysis.

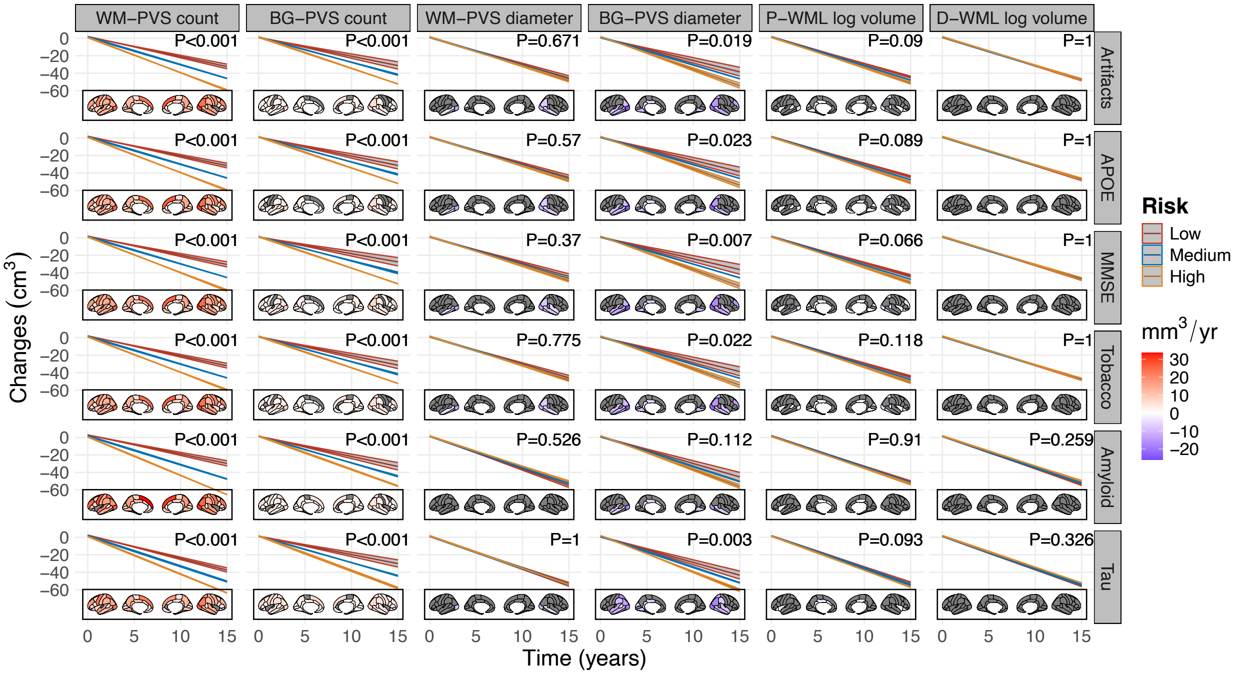


**Figure S10. Sensitivity analyses for the estimated trajectories of white matter atrophy in relation to PVS and WML markers.**
The effects of PVS and WML markers on the trajectories of total white matter volume were estimated from fully adjusted linear mixed-effects models with random intercepts and slopes for each individual participant and controlling for the same covariates as in the primary analysis (Fig. 2). For "Artifacts", cases where the estimate of the vascular marker may have been affected by MRI artifacts were excluded from the model. For "MMSE" plots, the covariate "CDR global score at the baseline" has been replaced with "MMSE score at the baseline". In all the other plots, the indicated variable was included as additional covariate in the model. "APOE" denotes the covariate for Apolipoprotein-E allele genotype, "MMSE" denotes the score from the Mini-Mental State Examination, "Tobacco" denotes history of tobacco smoking (positive or negative), "Amyloid" indicates the status for amyloid-beta pathology as assessed on cerebrospinal fluid and/or positron-emission tomography (positive or negative), and "Tau" indicates the status for tau pathology as assessed on cerebrospinal fluid and/or positron-emission tomography (positive or negative). Shaded areas indicate 95% confidence intervals. Three equally spaced values from the low-risk (red), medium-risk (blue), and high-risk (yellow) tertile (Table S16) of each vascular marker are shown. See also Table S13 for coefficients of sensitivity analysis.

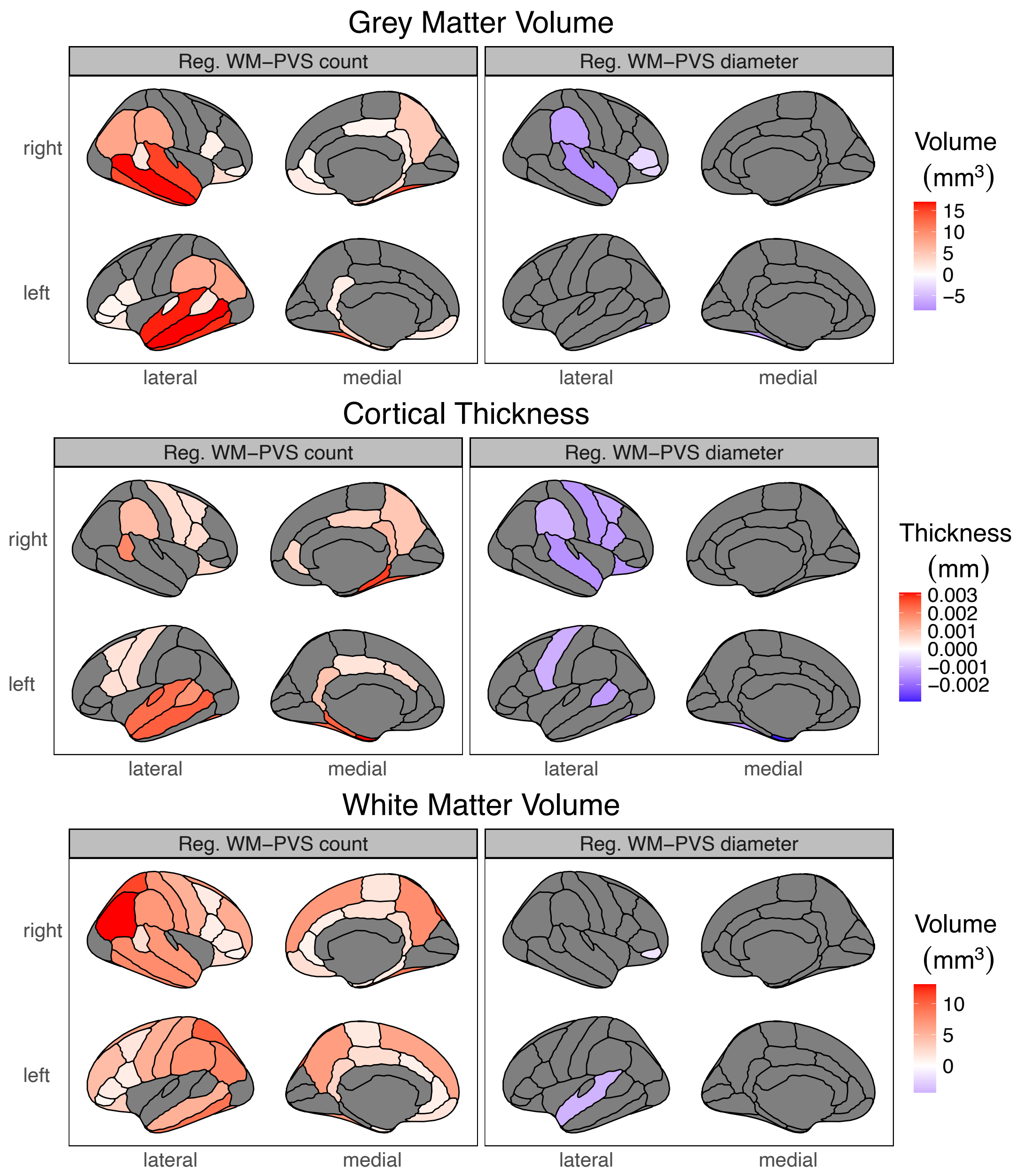


**Figure S11. Plots for the effect of baseline WM-PVS measured in individual lobes on the trajectories of atrophy in the corresponding lobe.**

The regional analysis across parcellations according to the Desikan-Killiany atlas for the grey matter (top panels), cortical thickness (middle panels) and white matter (bottom panels) reports the estimated volume or thickness preserved (positive values in red) or lost (negative values in blue) per year for each additional unit increase in the PVS count (left panels) and diameter (right panels) of the corresponding lobe. Only estimated values for regions that remained statistically significant after correction for multiple comparisons (68 comparisons) are shown; non-significant regions are greyed out. Estimates and corrected significance obtained from fully adjusted linear mixed-effects models with random intercepts and slopes for each individual participant (N=3389 and 14,229 timepoints MRI scans).

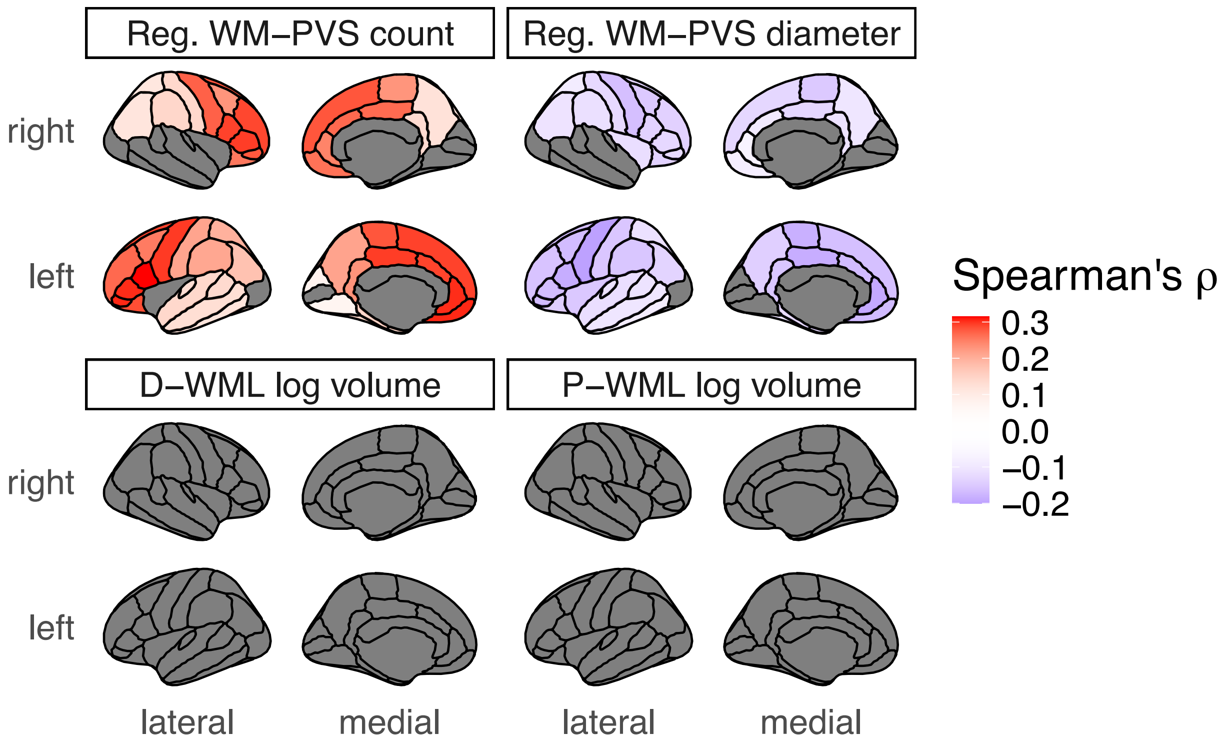


**Figure S12. Plots for the partial correlation between cerebral blood flow and PVS and WML markers.**

Partial correlation analyses assessing the relationship between PVS/WML markers and cerebral blood flow in non-demented individuals while controlling for age, sex, race, educational level, body mass index, history of diabetes, cardio-/cerebro-vascular disease, hypertension, dyslipidemia, family history of dementia, the employed arterial spin labeling technique, the total intracranial volume and the volume of the region where cerebral blood flow was assessed. Cortical parcellations were performed according to the Desikan-Killiany atlas. For PVS markers, regional PVS count and mean diameter were measured in each lobe and correlated with the cerebral blood flow of the corresponding parcellations. Estimated Spearman's ρ correlation coefficients are shown for all regions that were statistically significant after corrections for multiple comparisons (68 comparisons), while non-significant regions are greyed out. The analysis was performed on 1072 brain MRI scans with both T1-weighted images and perfusion data available from ADNI dataset.

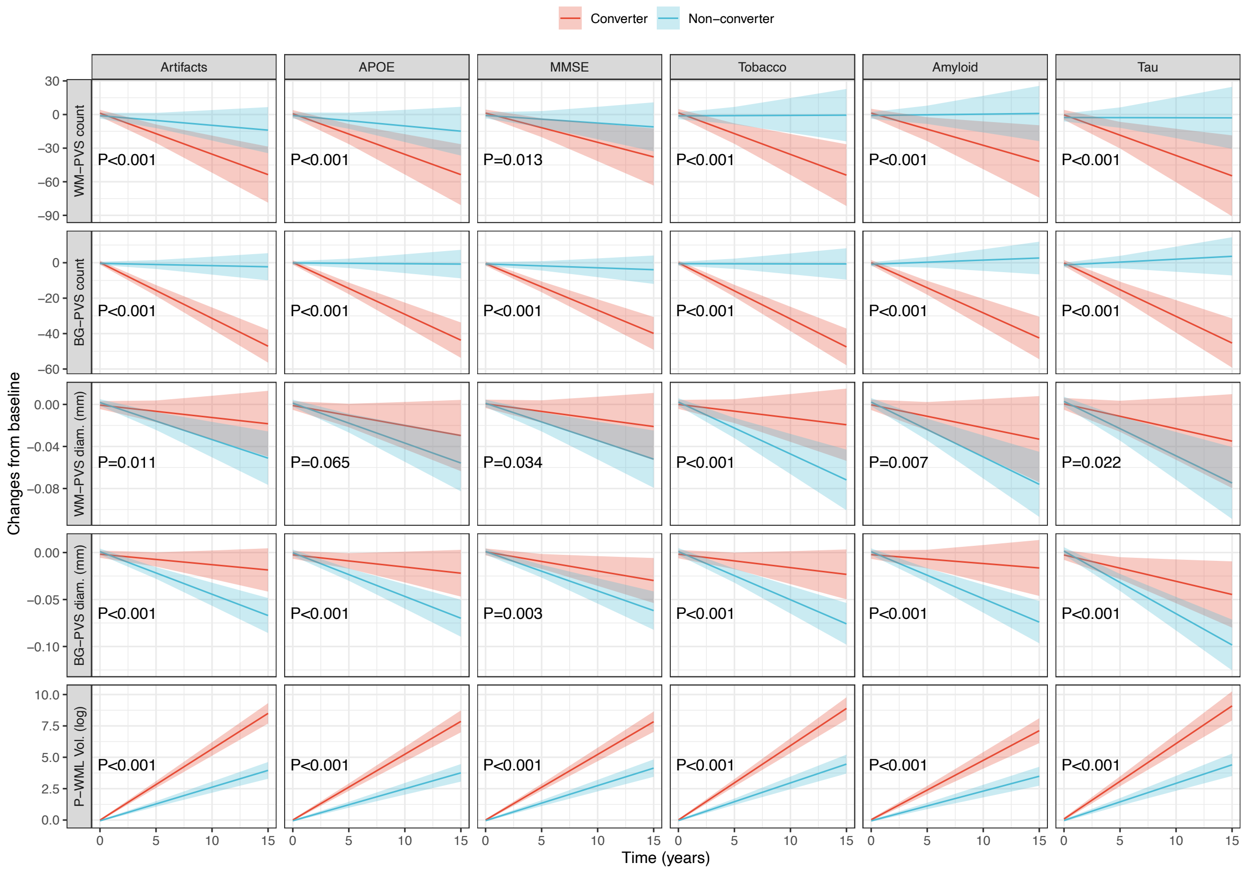


**Figure S13. Sensitivity analyses for the estimated trajectories of PVS and WML markers in relation to dementia conversion status.**

Estimated longitudinal trajectories of WM-PVS, BG-PVS, and P-WML markers for non-demented individuals who converted to dementia (converters, red) and those that did not convert to dementia (non-converters, cyan). Shaded areas indicate 95% confidence intervals. Trajectories are estimated from fully adjusted linear mixed-effect models with random intercepts and slopes for each individual participant (N=3389 and 14,229 timepoints MRI scans); the adjusted p-values (P) indicate whether the trajectories are significantly different between the groups after correction for multiple comparisons. Models included the same covariates as in the primary analysis (Fig. 3). For “Artifacts”, cases where the estimate of the vascular marker may have been affected by MRI artifacts were excluded from the model. For “MMSE” plots, the definition of conversion to dementia was based on MMSE score (dementia if MMSE equal to 24 or below). In all the other plots, the indicated variable was included as additional covariate in the model. “APOE” denotes the covariate for Apolipoprotein-E allele genotype, “MMSE” denotes the score from the Mini-Mental State Examination, “Tobacco” denotes history of tobacco smoking (positive or negative), “Amyloid” indicates the status for amyloid-beta pathology as assessed on cerebrospinal fluid and/or positron-emission tomography (positive or negative), and “Tau” indicates the status for tau pathology as assessed on cerebrospinal fluid and/or positron-emission tomography (positive or negative).

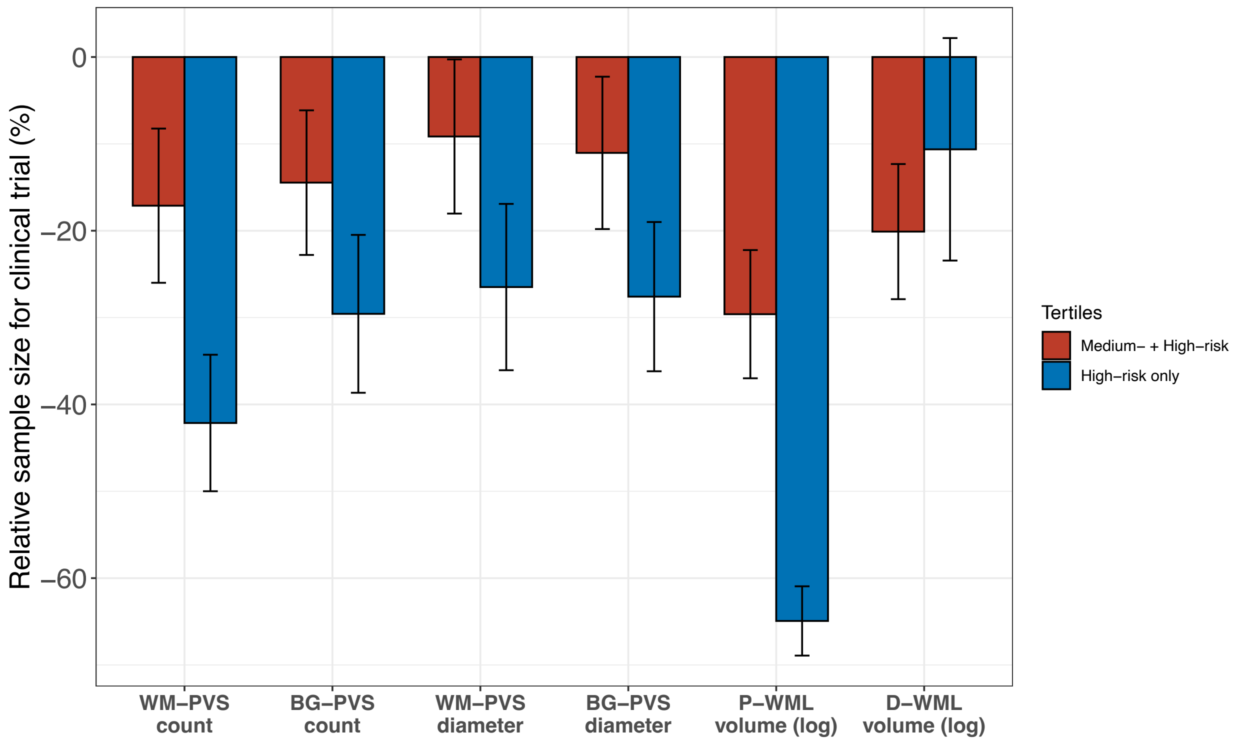


**Figure S14. Barplots for the relative sample size estimation in simulated clinical trials enriched using PVS or WML markers.**

Relative sample sizes for simulated clinical trials in non-demented individuals (i.e., MMSE score above 24) pooled from three studies (the Alzheimer Disease Neuroimaging Initiative, the Open Access Series of Imaging Studies, and the National Alzheimer's Coordinating Center). The simulations had statistical power of 80% at $\alpha$ = .05 and assumed a 30% treatment effect for slopes in cognitive decline, 1:1 allocation of treatment, total trial length of 48 months, and outcome measures every 12 months. Relative sample sizes and standard errors are across 500 bootstrap iterations of hypothetical trials. All available longitudinal cognitive data were used in these models adjusted for age. The reference model (without enrichment and with 100% inclusion) included all the tertiles. In the two enrichment models for each marker ("middle-risk+high-risk tertiles", red bars; "high-risk only tertile", blue bars), only participants in the indicated tertiles were included. Tertile limits for each marker are reported in Table S16.
