## Supplementary Tables for "Cerebral perivascular spaces as predictors of dementia risk and accelerated brain atrophy"

**Table S1. Overview of the MRI scanners and parameters employed to acquire the T1-weighted images analyzed in the study.**

| FS (T) | Manuf. | Model | TR (sec) | TE (sec) | TI (sec) | FA (°) | Native voxel vol. ($mm^3$) | OVERALL (N=20845) N (%) | ADNI (N=10977) N (%) | NACC (N=7819) N (%) | OASIS (N=2049) N (%) |
|---|---|---|---|---|---|---|---|---|---|---|---|
| 1.5 | GE | GENESIS SIGNA | 0.009 to 0.035 | 0.002 to 0.008 | 0.45 to 1 | 8 to 60 | 1.2 | 1641 (7.9) | 271 (2.5) | 1370 (17.5) | 0 |
| 1.5 | GE | SIGNA | 0.018 | 0.002 to 0.003 | NA | 25 | 1.8 | 50 (0.2) | 0 | 50 (0.6) | 0 |
| 1.5 | GE | SIGNA EXCITE | 0.008 to 0.029 | 0.002 to 0.004 | 0.5 to 1 | 8 to 25 | 1.1 | 999 (4.8) | 909 (8.3) | 90 (1.2) | 0 |
| 1.5 | GE | SIGNA HDx | 0.009 to 0.011 | 0.004 | 0.5 to 1 | 8 to 10 | 1.1 | 587 (2.8) | 425 (3.9) | 162 (2.1) | 0 |
| 1.5 | GE | Signa HDxt | 0.009 to 0.02 | 0.004 to 0.007 | 0.45 to 1 | 8 to 30 | 1.1 | 572 (2.7) | 303 (2.8) | 269 (3.4) | 0 |
| 1.5 | Hitachi | OASIS | NA | NA | NA | NA | 0.2 | 1 (0.0) | 0 | 1 (0.0) | 0 |
| 1.5 | Philips | Achieva | 0.009 | 0.004 | 1 | 8 | 1.1 | 70 (0.3) | 70 (0.6) | 0 | 0 |
| 1.5 | Philips | Eclipse 1.5T | 0.009 | 0.002 | NA | 15 | 1.5 | 74 (0.4) | 0 | 74 (0.9) | 0 |
| 1.5 | Philips | Gyroscan Intera | 0.008 to 0.009 | 0.004 | 0.761 to 1 | 8 | 1.1 | 13 (0.1) | 12 (0.1) | 1 (0.0) | 0 |
| 1.5 | Philips | Gyroscan NT | 0.009 | 0.004 | 1 | 8 | 1 | 19 (0.1) | 19 (0.2) | 0 | 0 |
| 1.5 | Philips | Intera | 0.008 to 0.01 | 0.003 to 0.004 | 1 to 1.076 | 8 | 1.1 | 359 (1.7) | 352 (3.2) | 7 (0.1) | 0 |
| 1.5 | Philips | Intera Achieva | 0.009 | 0.004 | 1 | 8 | 1.1 | 6 (0.0) | 6 (0.1) | 0 | 0 |
| 1.5 | Siemens | Aera | NA | NA | 0.8 | NA | 1 | 7 (0.0) | 0 | 7 (0.1) | 0 |
| 1.5 | Siemens | Avanto | 1.7 to 2.4 | 0.002 to 0.004 | 1 to 1.1 | 8 to 15 | 1.9 | 452 (2.2) | 443 (4.0) | 7 (0.1) | 2 (0.1) |
| 1.5 | Siemens | Espree | 0.011 to 2.4 | 0.003 to 0.006 | 1 to 1.1 | 8 to 25 | 1.8 | 36 (0.2) | 24 (0.2) | 12 (0.2) | 0 |
| 1.5 | Siemens | Magnetom VISION | NA | NA | 0.3 | NA | 1.8 | 2 (0.0) | 0 | 2 (0.0) | 0 |
| 1.5 | Siemens | Magnetom ESSENZA | 1.85 | 0.003 | 1.1 | 9 | 1 | 1 (0.0) | 0 | 1 (0.0) | 0 |
| 1.5 | Siemens | NUMARIS/4 | 2.4 | 0.004 | 1 | 8 | 1.9 | 2 (0.0) | 2 (0.0) | 0 | 0 |
| 1.5 | Siemens | Sonata | 1.9 to 3 | 0.003 to 0.005 | 0.93 to 1.1 | 8 to 20 | 1.8 | 498 (2.4) | 376 (3.4) | 88 (1.1) | 34 (1.7) |
| 1.5 | Siemens | Sonata Vision | 2.4 to 3 | 0.004 | 1 | 8 | 1.9 | 40 (0.2) | 40 (0.4) | 0 | 0 |
| 1.5 | Siemens | Symphony | 2 to 3 | 0.003 to 0.004 | 1 to 1.1 | 8 to 15 | 1.9 | 681 (3.3) | 594 (5.4) | 87 (1.1) | 0 |
| 1.5 | Siemens | Symphony Tim | 2.4 to 3 | 0.002 to 0.004 | 1 | 8 | 1.9 | 126 (0.6) | 124 (1.1) | 2 (0.0) | 0 |
| 1.5 | Siemens | Vision | 9.7 | 4 | NA | 10 | 1.3 | 172 (0.8) | 0 | 0 | 172 (8.4) |
| 3 | GE | Discovery MR750 | 0.006 to 0.009 | 0.002 to 0.003 | 0.4 to 1.06 | 8 to 12 | 1.1 | 2990 (14.3) | 958 (8.7) | 2032 (26.0) | 0 |
| 3 | GE | Discovery MR750w | 0.008 | 0.003 | 0.4 | 11 | 1.1 | 132 (0.6) | 132 (1.2) | 0 | 0 |
| 3 | GE | GENESIS SIGNA | 0.007 | 0.003 | 0.9 | 8 | 1.2 | 18 (0.1) | 18 (0.2) | 0 | 0 |
| 3 | GE | SIGNA EXCITE | 0.007 to 0.008 | 0.003 | 0.45 to 0.9 | 8 to 15 | 1.2 | 50 (0.2) | 44 (0.4) | 6 (0.1) | 0 |
| 3 | GE | SIGNA HDx | 0.007 | 0.003 | 0.4 to 0.9 | 8 to 11 | 1.3 | 53 (0.3) | 53 (0.5) | 0 | 0 |

| | | | | | | | | | | | |
|---|---|---|---|---|---|---|---|---|---|---|---|
| 3 | GE | Signa HDxt | 0.005 to 0.02 | 0.002 to 0.004 | 0.4 to 0.9 | 8 to 27 | 1.2 | 658 (3.2) | 427 (3.9) | 231 (3.0) | 0 |
| 3 | GE | Signa MR360 | 2.1 | 0.003 | 0.9 | 8 | 1 | 1 (0.0) | 0 | 1 (0.0) | 0 |
| 3 | GE | SIGNA PET/MR | 0.008 | 0.003 | 0.4 | 11 | 1.3 | 77 (0.4) | 0 | 77 (1.0) | 0 |
| 3 | GE | SIGNA Premier | 0.007 | 0.003 | 0.4 to 0.45 | 11 to 12 | 1 | 89 (0.4) | 56 (0.5) | 33 (0.4) | 0 |
| 3 | GE | SIGNA UHP | 0.007 to 2.077 | 0.003 | 0.4 to 0.9 | 8 to 11 | 1 | 10 (0.0) | 7 (0.1) | 3 (0.0) | 0 |
| 3 | Philips | Achieva | 0.006 to 0.007 | 0.003 | 0.805 to 0.9 | 8 to 9 | 1.2 | 889 (4.3) | 597 (5.4) | 292 (3.7) | 0 |
| 3 | Philips | Achieva dStream | 0.006 to 0.007 | 0.003 | NA | 8 to 9 | 1 | 186 (0.9) | 138 (1.3) | 48 (0.6) | 0 |
| 3 | Philips | GEMINI | 0.007 | 0.003 | NA | 9 | 1.2 | 44 (0.2) | 35 (0.3) | 9 (0.1) | 0 |
| 3 | Philips | Ingenia | 0.007 | 0.003 | 0.9 | 9 | 1.1 | 209 (1.0) | 209 (1.9) | 0 | 0 |
| 3 | Philips | Ingenia Elition X | 0.007 | 0.003 | NA | 9 | 1 | 14 (0.1) | 14 (0.1) | 0 | 0 |
| 3 | Philips | Ingenuity | 0.007 | 0.003 | NA | 9 | 1.3 | 19 (0.1) | 16 (0.1) | 3 (0.0) | 0 |
| 3 | Philips | Intera | 0.007 | 0.003 | 0.845 to 0.869 | 8 to 9 | 1.2 | 273 (1.3) | 273 (2.5) | 0 | 0 |
| 3 | Philips | Intera Achieva | 0.007 | 0.003 | 0.855 | 8 | 1.2 | 1 (0.0) | 1 (0.0) | 0 | 0 |
| 3 | Siemens | Allegra | 2.3 to 2.5 | 0.003 to 0.004 | 0.9 to 1.1 | 8 to 9 | 1.1 | 136 (0.7) | 108 (1.0) | 28 (0.4) | 0 |
| 3 | Siemens | Biograph mMR | 2.3 to 2.4 | 0.002 to 0.003 | 0.9 to 1 | 8 to 9 | 1.3 | 584 (2.8) | 26 (0.2) | 5 (0.1) | 553 (27.0) |
| 3 | Siemens | Magnetom Prisma Fit | 2.3 | 0.003 | 0.9 | 9 | 1 | 12 (0.1) | 12 (0.1) | 0 | 0 |
| 3 | Siemens | Magnetom Vida | 2.3 to 2.4 | 0.003 | 0.9 | 8 to 9 | 1.2 | 219 (1.1) | 3 (0.0) | 1 (0.0) | 215 (10.5) |
| 3 | Siemens | Prisma | 2.3 to 2.4 | 0.002 to 0.003 | 0.9 to 1.06 | 8 to 9 | 1 | 957 (4.6) | 255 (2.3) | 702 (9.0) | 0 |
| 3 | Siemens | Prisma fit | 1.8 to 2.4 | 0.002 to 0.005 | 0.9 to 1 | 8 to 10 | 1 | 810 (3.9) | 663 (6.0) | 146 (1.9) | 1 (0.0) |
| 3 | Siemens | Skyra | 1.8 to 2.3 | 0.002 to 0.003 | 0.649 to 0.962 | 8 to 10 | 1.1 | 1696 (8.1) | 489 (4.5) | 1207 (15.4) | 0 |
| 3 | Siemens | Skyra fit | 2.3 | 0.003 | 0.9 | 9 | 1 | 26 (0.1) | 26 (0.2) | 0 | 0 |
| 3 | Siemens | Trio | 1.62 to 2.5 | 0.002 to 0.004 | 0.9 to 1.1 | 7 to 15 | 1.2 | 144 (0.7) | 121 (1.1) | 23 (0.3) | 0 |
| 3 | Siemens | TrioTim | 1.31 to 3.2 | 0.002 to 0.455 | 0.9 to 1.2 | 7 to 120 | 1.1 | 3098 (14.9) | 1547 (14.1) | 479 (6.1) | 1072 (52.3) |
| 3 | Siemens | Verio | 1.7 to 2.3 | 0.002 to 0.004 | 0.9 | 8 to 9 | 1.2 | 805 (3.9) | 767 (7.0) | 38 (0.5) | 0 |
| Not available / Unknown | | | 0.007 to 2.3 | 0.002 to 0.005 | 0.4 to 0.9 | 8 to 60 | 1 | 237 (1.1) | 12 (0.1) | 225 (2.9) | 0 |

**Table S2. Multi-variate associations of PVS and WML markers with the demographic and clinical covariates included in all the models of this study.**

| Variable | Estimate | Standard error | T value | Adjusted P value |
|---|---|---|---|---|
| **WM-PVS count** | | | | |
| Age (years) | **-4.31** | **0.14** | **-30.43** | **<0.001** |
| Female sex | **-24.35** | **3.12** | **-7.80** | **<0.001** |
| Education (years) | **-1.50** | **0.44** | **-3.40** | **0.001** |
| Race (compared to White) | | | | |
| Black | **27.18** | **4.13** | **6.58** | **<0.001** |
| Asian | 28.27 | 14.07 | 2.01 | 0.089 |
| American Indians | 9.36 | 9.53 | 0.98 | 0.652 |
| More than one reported | 1.03 | 20.64 | 0.05 | 1 |
| CDR global score | **-62.10** | **3.20** | **-19.42** | **<0.001** |
| Body Mass Index (kg/m$^2$) | **1.97** | **0.26** | **7.63** | **<0.001** |
| Dyslipidemia | 2.02 | 2.64 | 0.77 | 0.887 |
| Hypertension | **-6.43** | **2.78** | **-2.32** | **0.041** |
| Diabetes | **-11.14** | **4.04** | **-2.76** | **0.012** |
| History of cardio-/cerebro-vascular disease | -1.68 | 2.88 | -0.58 | 1 |
| Family history of dementia | -4.13 | 2.58 | -1.60 | 0.219 |
| **BG-PVS count** | | | | |
| Age (years) | **-1.43** | **0.05** | **-30.68** | **<0.001** |
| Female sex | **-5.89** | **1.03** | **-5.72** | **<0.001** |
| Education (years) | **0.43** | **0.15** | **2.94** | **0.006** |
| Race (compared to White) | | | | |
| Black | **11.96** | **1.36** | **8.77** | **<0.001** |
| Asian | **11.30** | **4.64** | **2.43** | **0.03** |
| American Indians | **7.15** | **3.15** | **2.27** | **0.046** |
| More than one reported | **15.82** | **6.81** | **2.32** | **0.04** |
| CDR global score | **-16.96** | **1.06** | **-16.07** | **<0.001** |
| Body Mass Index (kg/m$^2$) | **0.91** | **0.09** | **10.68** | **<0.001** |
| Dyslipidemia | 1.14 | 0.87 | 1.31 | 0.379 |
| Hypertension | **-4.57** | **0.92** | **-4.99** | **<0.001** |
| Diabetes | **-3.37** | **1.33** | **-2.53** | **0.023** |
| History of cardio-/cerebro-vascular disease | -1.89 | 0.95 | -1.99 | 0.093 |
| Family history of dementia | 0.54 | 0.85 | 0.63 | 1 |
| **WM-PVS diameter** | | | | |
| Age (years) | **0.03** | **0.00** | **17.81** | **<0.001** |
| Female sex | **0.18** | **0.03** | **5.39** | **<0.001** |
| Education (years) | 0.00 | 0.00 | -0.31 | 1 |
| Race (compared to White) | | | | |
| Black | **-0.15** | **0.04** | **-3.28** | **0.002** |

| | | | | |
|---|---|---|---|---|
| Asian | -0.09 | 0.15 | -0.56 | 1 |
| American Indians | **-0.42** | **0.10** | **-4.14** | **<0.001** |
| More than one reported | **-0.61** | **0.22** | **-2.72** | **0.013** |
| CDR global score | **0.19** | **0.03** | **5.44** | **<0.001** |
| Body Mass Index (kg/m$^2$) | **-0.02** | **0.00** | **-5.63** | **<0.001** |
| Dyslipidemia | 0.03 | 0.03 | 0.98 | 0.658 |
| Hypertension | **0.21** | **0.03** | **6.86** | **<0.001** |
| Diabetes | **-0.16** | **0.04** | **-3.76** | **<0.001** |
| History of cardio-/cerebro-vascular disease | -0.04 | 0.03 | -1.23 | 0.436 |
| Family history of dementia | 0.01 | 0.03 | 0.25 | 1 |
| **BG-PVS diameter** | | | | |
| Age (years) | **0.02** | **0.00** | **19.30** | **<0.001** |
| Female sex | **0.09** | **0.02** | **4.13** | **<0.001** |
| Education (years) | **-0.01** | **0.00** | **-2.82** | **0.01** |
| Race (compared to White) | | | | |
| Black | **-0.19** | **0.03** | **-6.56** | **<0.001** |
| Asian | -0.20 | 0.10 | -2.03 | 0.085 |
| American Indians | **-0.15** | **0.07** | **-2.26** | **0.047** |
| More than one reported | **-0.38** | **0.14** | **-2.64** | **0.017** |
| CDR global score | **0.18** | **0.02** | **8.03** | **<0.001** |
| Body Mass Index (kg/m$^2$) | **-0.02** | **0.00** | **-9.19** | **<0.001** |
| Dyslipidemia | -0.03 | 0.02 | -1.92 | 0.111 |
| Hypertension | **0.12** | **0.02** | **6.39** | **<0.001** |
| Diabetes | -0.05 | 0.03 | -1.77 | 0.152 |
| History of cardio-/cerebro-vascular disease | **0.05** | **0.02** | **2.34** | **0.039** |
| Family history of dementia | -0.02 | 0.02 | -1.16 | 0.494 |
| **P-WML log volume** | | | | |
| Age (years) | **0.15** | **0.00** | **47.25** | **<0.001** |
| Female sex | **0.64** | **0.07** | **9.32** | **<0.001** |
| Education (years) | -0.01 | 0.01 | -1.50 | 0.267 |
| Race (compared to White) | | | | |
| Black | 0.03 | 0.09 | 0.31 | 1 |
| Asian | -0.56 | 0.31 | -1.82 | 0.138 |
| American Indians | 0.15 | 0.21 | 0.72 | 0.943 |
| More than one reported | -0.25 | 0.45 | -0.55 | 1 |
| CDR global score | **1.33** | **0.07** | **19.06** | **<0.001** |
| Body Mass Index (kg/m$^2$) | **-0.03** | **0.01** | **-5.98** | **<0.001** |
| Dyslipidemia | 0.00 | 0.06 | 0.05 | 1 |
| Hypertension | **0.45** | **0.06** | **7.36** | **<0.001** |
| Diabetes | 0.13 | 0.09 | 1.44 | 0.298 |
| History of cardio-/cerebro-vascular disease | 0.08 | 0.06 | 1.24 | 0.433 |
| Family history of dementia | -0.10 | 0.06 | -1.72 | 0.17 |

| **D-WML log volume** | | | | |
|---|---|---|---|---|
| Age (years) | **0.02** | **0.00** | **10.38** | **<0.001** |
| Female sex | **0.28** | **0.05** | **5.51** | **<0.001** |
| Education (years) | -0.01 | 0.01 | -1.18 | 0.479 |
| Race (compared to White) | | | | |
| Black | **0.32** | **0.07** | **4.83** | **<0.001** |
| Asian | -0.21 | 0.23 | -0.94 | 0.698 |
| American Indians | -0.18 | 0.15 | -1.15 | 0.504 |
| More than one reported | -0.66 | 0.33 | -1.97 | 0.097 |
| CDR global score | **-0.25** | **0.05** | **-4.83** | **<0.001** |
| Body Mass Index (kg/m$^2$) | **-0.01** | **0.00** | **-2.83** | **0.009** |
| Dyslipidemia | -0.03 | 0.04 | -0.65 | 1 |
| Hypertension | **0.21** | **0.04** | **4.73** | **<0.001** |
| Diabetes | -0.09 | 0.07 | -1.42 | 0.31 |
| History of cardio-/cerebro-vascular disease | -0.03 | 0.05 | -0.58 | 1 |
| Family history of dementia | 0.00 | 0.04 | -0.09 | 1 |

This analysis was run in the entire study population (N=10,004), including non-demented and demented subjects.

**Table S3. Multi-variate associations of PVS and WML markers with the clinical covariates assessed in the sensitivity analyses.**

| Variable | Total number of subjects | Estimate | Standard error | T value | Adjusted P value |
|---|---|---|---|---|---|
| **WM-PVS count** | | | | | |
| APOE (ref. ε3ε3) | | | | | |
| ε2ε2 | | -40.02 | 23.01 | -1.74 | 0.164 |
| ε2ε3 | 8599 | 1.48 | 4.79 | 0.31 | 1 |
| ε2ε4 | | 3.61 | 8.66 | 0.42 | 1 |
| ε3ε4 | | 0.55 | 3.07 | 0.18 | 1 |
| ε4ε4 | | -8.51 | 5.42 | -1.57 | 0.233 |
| Baseline MMSE | 7036 | **6.43** | **0.46** | **14.11** | **<0.001** |
| Tobacco smoking | 9267 | -5.29 | 2.74 | -1.93 | 0.106 |
| A-beta positivity | 3906 | -8.77 | 4.06 | -2.16 | 0.062 |
| Tau positivity | 2466 | 8.72 | 4.92 | 1.77 | 0.153 |
| **BG-PVS count** | | | | | |
| APOE (ref. ε3ε3) | | | | | |
| ε2ε2 | | -7.57 | 7.42 | -1.02 | 0.616 |
| ε2ε3 | 8599 | 1.65 | 1.55 | 1.07 | 0.573 |
| ε2ε4 | | 0.49 | 2.79 | 0.18 | 1 |
| ε3ε4 | | -1.19 | 0.99 | -1.20 | 0.461 |
| ε4ε4 | | **-5.07** | **1.75** | **-2.90** | **0.007** |
| Baseline MMSE | 7036 | **1.56** | **0.14** | **11.16** | **<0.001** |
| Tobacco smoking | 9267 | **-7.24** | **0.90** | **-8.03** | **<0.001** |
| A-beta positivity | 3906 | **-3.92** | **1.45** | **-2.71** | **0.014** |
| Tau positivity | 2466 | -1.62 | 1.69 | -0.96 | 0.679 |
| **WM-PVS diameter** | | | | | |
| APOE (ref. ε3ε3) | | | | | |
| ε2ε2 | | 0.21 | 0.25 | 0.83 | 0.81 |
| ε2ε3 | 8599 | -0.09 | 0.05 | -1.72 | 0.17 |
| ε2ε4 | | 0.04 | 0.09 | 0.44 | 1 |
| ε3ε4 | | 0.02 | 0.03 | 0.61 | 1 |
| ε4ε4 | | **0.16** | **0.06** | **2.73** | **0.013** |
| Baseline MMSE | 7036 | **-0.02** | **0.01** | **-2.94** | **0.007** |
| Tobacco smoking | 9267 | **0.16** | **0.03** | **5.59** | **<0.001** |
| A-beta positivity | 3906 | **0.15** | **0.05** | **2.95** | **0.006** |
| Tau positivity | 2466 | **0.17** | **0.06** | **2.80** | **0.01** |
| **BG-PVS diameter** | | | | | |
| APOE (ref. ε3ε3) | | | | | |
| ε2ε2 | | 0.05 | 0.16 | 0.33 | 1 |
| ε2ε3 | 8599 | -0.02 | 0.03 | -0.72 | 0.943 |
| ε2ε4 | | -0.04 | 0.06 | -0.73 | 0.934 |
| ε3ε4 | | 0.02 | 0.02 | 0.79 | 0.856 |
| ε4ε4 | | **0.09** | **0.04** | **2.31** | **0.041** |

| | | | | | |
|---|---|---|---|---|---|
| Baseline MMSE | 7036 | **-0.01** | **0.00** | **-3.63** | **<0.001** |
| Tobacco smoking | 9267 | **0.12** | **0.02** | **6.08** | **<0.001** |
| A-beta positivity | 3906 | 0.07 | 0.03 | 2.05 | 0.081 |
| Tau positivity | 2466 | **0.10** | **0.04** | **2.39** | **0.034** |
| **P-WML log volume** | | | | | |
| APOE (ref. ε3ε3) | 8599 | | | | |
| ε2ε2 | | 0.51 | 0.50 | 1.03 | 0.609 |
| ε2ε3 | | -0.01 | 0.10 | -0.14 | 1 |
| ε2ε4 | | 0.19 | 0.19 | 1.01 | 0.624 |
| ε3ε4 | | -0.02 | 0.07 | -0.30 | 1 |
| ε4ε4 | | **0.26** | **0.12** | **2.22** | **0.053** |
| Baseline MMSE | 7036 | **-0.14** | **0.01** | **-14.05** | **<0.001** |
| Tobacco smoking | 9267 | **0.19** | **0.06** | **3.14** | **0.003** |
| A-beta positivity | 3906 | **0.40** | **0.09** | **4.26** | **<0.001** |
| Tau positivity | 2466 | -0.22 | 0.11 | -1.91 | 0.113 |
| **D-WML log volume** | | | | | |
| APOE (ref. ε3ε3) | 8599 | | | | |
| ε2ε2 | | 0.07 | 0.37 | 0.19 | 1 |
| ε2ε3 | | -0.09 | 0.08 | -1.11 | 0.536 |
| ε2ε4 | | 0.09 | 0.14 | 0.62 | 1 |
| ε3ε4 | | -0.06 | 0.05 | -1.21 | 0.449 |
| ε4ε4 | | **0.24** | **0.09** | **2.70** | **0.014** |
| Baseline MMSE | 7036 | 0.01 | 0.01 | 1.77 | 0.153 |
| Tobacco smoking | 9267 | -0.01 | 0.04 | -0.16 | 1 |
| A-beta positivity | 3906 | **0.17** | **0.07** | **2.44** | **0.029** |
| Tau positivity | 2466 | 0.10 | 0.09 | 1.22 | 0.444 |

These general linear models included the same covariates as in the primary analysis (Table S2), and the indicated variable was included as additional covariate. For “MMSE”, the variable “MMSE score at the baseline” was used in place of the CDR score.

**Table S4. Estimated marginal means for PVS and WML markers.**

| Marker | Estimated marginal mean ± standard error | | Adj. P value |
|---|---|---|---|
| | Non-demented | Demented | |
| WM-PVS count | 445 ± 6 | 371 ± 7 | <0.001 |
| BG-PVS count | 144 ± 2 | 123 ± 2 | <0.001 |
| WM-PVS diameter (mm) | 1.96 ± 0.01 | 1.99 ± 0.01 | <0.001 |
| BG-PVS diameter (mm) | 1.57 ± 0 | 1.60 ± 0 | <0.001 |
| P-WML log volume | 4.48 ± 0.12 | 5.89 ± 0.15 | <0.001 |
| D-WML log volume | 2.56 ± 0.09 | 2.41 ± 0.11 | 0.051 |

Marginal means were estimated controlling for the same covariates as in the primary analysis (Table S2).

**Table S5. Sensitivity analysis on the ANCOVA models.**

| Covariate | N. of demented patients / total | Marker | Estimated marginal mean ± standard error | | Adj. P value |
|---|---|---|---|---|---|
| | | | Non-demented | Demented | |
| **Artifacts** | 1123 / 9955 | WM-PVS count | 446 ± 6 | 372 ± 7 | <0.001 |
| | | BG-PVS count | 144 ± 2 | 123 ± 2 | <0.001 |
| | | WM-PVS diameter | 1.96 ± 0.01 | 1.99 ± 0.01 | <0.001 |
| | | BG-PVS diameter | 1.57 ± 0 | 1.6 ± 0 | <0.001 |
| | | P-WML log volume | 4.47 ± 0.12 | 5.89 ± 0.15 | <0.001 |
| | | D-WML log volume | 2.56 ± 0.09 | 2.43 ± 0.11 | 0.1 |
| **APOE** | 974 / 8599 | WM-PVS count | 437 ± 7 | 362 ± 8 | <0.001 |
| | | BG-PVS count | 139 ± 2 | 119 ± 3 | <0.001 |
| | | WM-PVS diameter | 1.97 ± 0.01 | 2 ± 0.01 | <0.001 |
| | | BG-PVS diameter | 1.58 ± 0.01 | 1.6 ± 0.01 | <0.001 |
| | | P-WML log volume | 4.67 ± 0.16 | 6.05 ± 0.19 | <0.001 |
| | | D-WML log volume | 2.64 ± 0.12 | 2.49 ± 0.14 | 0.08 |
| **Baseline MMSE** | 1255 / 7036 | WM-PVS count | 450 ± 7 | 403 ± 8 | <0.001 |
| | | BG-PVS count | 143 ± 2 | 131 ± 2 | <0.001 |
| | | WM-PVS diameter | 1.95 ± 0.01 | 1.96 ± 0.01 | 0.026 |
| | | BG-PVS diameter | 1.57 ± 0 | 1.58 ± 0.01 | 0.004 |
| | | P-WML log volume | 4.47 ± 0.14 | 5.37 ± 0.16 | <0.001 |
| | | D-WML log volume | 2.52 ± 0.11 | 2.41 ± 0.12 | 0.205 |
| **History of tobacco smoking** | 1053 / 9267 | WM-PVS count | 447 ± 6 | 372 ± 7 | <0.001 |
| | | BG-PVS count | 1.96 ± 0.01 | 1.99 ± 0.01 | <0.001 |
| | | WM-PVS diameter | 143 ± 2 | 122 ± 2 | <0.001 |
| | | BG-PVS diameter | 1.57 ± 0 | 1.6 ± 0 | <0.001 |
| | | P-WML log volume | 4.48 ± 0.13 | 5.89 ± 0.15 | <0.001 |
| | | D-WML log volume | 2.55 ± 0.09 | 2.39 ± 0.11 | 0.037 |
| **A-beta positivity** | 327 / 3906 | WM-PVS count | 445 ± 10 | 391 ± 12 | <0.001 |
| | | BG-PVS count | 142 ± 4 | 128 ± 4 | <0.001 |
| | | WM-PVS diameter | 1.97 ± 0.01 | 1.98 ± 0.01 | 0.306 |
| | | BG-PVS diameter | 1.58 ± 0.01 | 1.58 ± 0.01 | 0.553 |
| | | P-WML log volume | 4.94 ± 0.23 | 6.11 ± 0.27 | <0.001 |
| | | D-WML log volume | 2.55 ± 0.17 | 2.41 ± 0.2 | 0.43 |
| **Tau positivity** | 217 / 2466 | WM-PVS count | 453 ± 12 | 380 ± 14 | <0.001 |
| | | BG-PVS count | 137 ± 4 | 121 ± 5 | <0.001 |
| | | WM-PVS diameter | 1.98 ± 0.01 | 2.01 ± 0.02 | 0.01 |
| | | BG-PVS diameter | 1.59 ± 0.01 | 1.6 ± 0.01 | 0.169 |
| | | P-WML log volume | 4.84 ± 0.27 | 6.3 ± 0.32 | <0.001 |
| | | D-WML log volume | 2.63 ± 0.2 | 2.46 ± 0.24 | 0.487 |

These models were adjusted for the same covariates as in the primary analysis (Table S2). For “Artifacts”, cases where the estimate of the vascular marker may have been affected by MRI artifacts or the MRI data were acquired after the injection of a contrast agent were excluded from the model. For “MMSE”, the definition of dementia was based on the MMSE score rather than CDR. For all the other models, the indicated variable was included as additional covariate in the model.

**Table S6. Baseline characteristics of non-demented individuals included in the analysis of the hazard ratio for dementia.**

| Characteristic | Overall (N=7518) | ADNI (N=1931) | NACC (N=4433) | OASIS (N=1154) |
|---|---|---|---|---|
| Age at baseline – yr | 71.0 ± 9.5 | 73.1 ± 7.1 | 70.3 ± 10.3 | 69.9 ± 9.1 |
| Female – % | 56.4 | 46.6 | 61 | 55.5 |
| Education – yr | 15.9 ± 2.9 | 16.1 ± 2.7 | 15.8 ± 3.0 | 15.9 ± 2.7 |
| Race – % | | | | |
| White | 86.1 | 91.6 | 84.1 | 84.6 |
| Black | 11.1 | 5 | 12.9 | 14.7 |
| Asian | 0.8 | 0.2 | 1.2 | 0 |
| American Indians | 1.7 | 2 | 1.8 | 0.4 |
| More than one reported | 0.4 | 1.2 | 0 | 0.3 |
| Body Mass Index – kg/m$^2$ | 27.2 ± 5.2 | 27.1 ± 4.9 | 27.0 ± 5.2 | 27.9 ± 5.4 |
| Dyslipidemia – % | 55.4 | 53.6 | 58.1 | 48.1 |
| Hypertension – % | 50.2 | 48.8 | 51.2 | 48.6 |
| Diabetes – % | 12.5 | 10.3 | 14 | 10.8 |
| History of cardio-/cerebro-vascular disease – % | 28.1 | 36.5 | 25.2 | 24.9 |
| Family history of dementia – % | 53.6 | 53.6 | 52.2 | 59.5 |
| History of tobacco smoking* – % | 26.7 | 50.5 | 44.8 | 37.1 |
| APOE* – % | | | | |
| ε2ε2 | 0.3 | 0.1 | 0.4 | 0.6 |
| ε2ε3 | 9.6 | 7.8 | 10.3 | 10.6 |
| ε2ε4 | 2.5 | 2 | 2.5 | 3.7 |
| ε3ε3 | 50.4 | 48 | 52.1 | 48.7 |
| ε3ε4 | 30.8 | 33.1 | 29.6 | 31 |
| ε4ε4 | 6.3 | 9 | 5.3 | 5.3 |
| A-beta positivity* – % | 49.5 | 63.9 | 40.5 | 34 |
| Tau positivity* – % | 54.9 | 65.6 | 39.2 | 43 |
| Median follow-up – yr | 4.1 (2.3 to 7.1) | 4.0 (2.1 to 6.8) | 4.1 (2.3 to 6.6) | 6.1 (3.2 to 10.1) |
| Number of conversions to dementia | 1493 | 469 | 803 | 221 |
| WM-PVS count | 435 (346 to 537) | 382 (308 to 480) | 459 (369 to 563) | 434 (350 to 525) |

| | | | | |
|---|---|---|---|---|
| BG-PVS count | 135 (107 to 167) | 105 (88 to 132) | 147 (122 to 177) | 133 (113 to 158) |
| WM-PVS diameter – mm | 1.96 (1.88 to 2.07) | 2.07 (1.96 to 2.18) | 1.93 (1.86 to 2.02) | 1.96 (1.88 to 2.05) |
| BG-PVS diameter – mm | 1.57 (1.52 to 1.64) | 1.65 (1.57 to 1.71) | 1.54 (1.50 to 1.59) | 1.57 (1.53 to 1.62) |
| P-WML volume – $mm^3$ | 254.50 (0 to 1452) | 581 (46 to 1920.50) | 143 (0 to 1189) | 223 (0 to 1521) |
| D-WML volume – $mm^3$ | 17 (1 to 74) | 37 (7 to 107) | 8 (0 to 55) | 25 (4 to 94.75) |

Plus–minus values are means ± standard deviations, entries with parentheses are medians (interquartile range). Data from three studies — the Alzheimer's Disease Neuroimaging Initiative (ADNI), the National Alzheimer's Coordinating Center (NACC), and the Open Access Series of Imaging Studies (OASIS) — are shown. *Total number of subjects with available history of tobacco smoking, Apolipoprotein E (APOE) genotype, amyloid-beta status and tau status were 3918, 6968, 3774, and 2434, respectively.

**Table S7. Sensitivity analysis on the HR for dementia**

| Characteristic and overall prevalence (%) or mean ± standard deviation | | N. of conversions to dementia / total | HR [95% CI] | | | | |
|---|---|---|---|---|---|---|---|
| | | | WM-PVS count | BG-PVS count | WM-PVS diameter | BG-PVS diameter | P-WML |
| Artifacts – % | 0.4 | 1487 / 7490 | 0.80 [0.76 0.84] | 0.91 [0.89 0.93] | 1.10 [1.05 1.15] | 1.21 [1.13 1.30] | 1.14 [1.11 1.16] |
| APOE – % | | 1418 / 6914 | 0.79 [0.75 0.83] | 0.91 [0.89 0.93] | 1.09 [1.04 1.14] | 1.19 [1.11 1.27] | 1.14 [1.11 1.17] |
| ε2ε2 | 0.3 | | | | | | |
| ε2ε3 | 9.6 | | | | | | |
| ε2ε4 | 2.5 | | | | | | |
| ε3ε3 | 50.4 | | | | | | |
| ε3ε4 | 30.8 | | | | | | |
| ε4ε4 | 6.3 | | | | | | |
| Baseline MMSE | 28.5 ± 1.5 | 1259 / 5311 | 0.86 [0.81 0.91] | 0.93 [0.91 0.95] | 1.09 [1.04 1.14] | 1.13 [1.05 1.22] | 1.10 [1.07 1.14] |
| History of tobacco smoking – % | 37.1 | 1056 / 3918 | 0.80 [0.76 0.85] | 0.92 [0.9 0.94] | 1.09 [1.04 1.14] | 1.17 [1.09 1.26] | 1.12 [1.09 1.15] |
| A-beta positivity – % | 49.7 | 697 / 3774 | 0.80 [0.74 0.86] | 0.91 [0.89 0.94] | 1.05 [0.99 1.12] | 1.15 [1.05 1.27] | 1.12 [1.08 1.16] |
| Tau positivity – % | 54.9 | 465 / 2434 | 0.77 [0.70 0.85] | 0.91 [0.88 0.94] | 1.08 [1.01 1.17] | 1.18 [1.06 1.32] | 1.13 [1.08 1.18] |

These models were adjusted for the same covariates as in the primary analysis (Fig. 1). For “Artifacts”, cases where the estimate of the vascular marker may have been affected by MRI artifacts or the MRI data were acquired after the injection of a contrast agent were excluded from the model. For “MMSE”, the definition of dementia was based on the MMSE score rather than CDR, and the corresponding MMSE at the baseline was used as covariate in place of CDR score at the baseline. For all the other models, the indicated variable was included as additional covariate in the model. Hazard ratios (HR) for WM-PVS and BG-PVS count are per 100 and 10 units increase, respectively, for WM-PVS and BG-PVS diameter are per 0.1 mm units increase, and for P-WML volume are per 1 logarithmic unit increase. See also Fig. S7 for spline plots.

**Table S8. Baseline characteristics of non-demented individuals included in the analysis of the linear mixed-effects model for brain atrophy and for the longitudinal trajectories of PVS and WML markers.**

| Characteristic | Overall (N=3389) | ADNI (N=1833) | NACC (N=1176) | OASIS (N=380) |
|---|---|---|---|---|
| Age at baseline – yr | 70.7 ± 9.8 | 73.3 ± 7.0 | 67.8 ± 12.1 | 67.5 ± 9.6 |
| Female – % | 53.3 | 46.0 | 63.4 | 56.8 |
| Education – yr | 15.9 ± 2.9 | 16.1 ± 2.7 | 15.7 ± 3.1 | 15.9 ± 2.6 |
| Race – % | | | | |
| White | 89.2 | 92.4 | 84.6 | 87.8 |
| Black | 7.9 | 4.4 | 12.4 | 11.4 |
| Asian | 0.5 | 0.3 | 0.9 | 0 |
| American Indians | 1.7 | 1.7 | 2 | 0.5 |
| More than one reported | 0.7 | 1.2 | 0 | 0.3 |
| Body Mass Index – $kg/m^2$ | 27.0 ± 5.0 | 27.0 ± 4.8 | 27.3 ± 5.2 | 27.9 ± 5.0 |
| Dyslipidemia – % | 51.5 | 53.4 | 50.8 | 44.3 |
| Hypertension – % | 46.4 | 48.9 | 42.6 | 45.9 |
| Diabetes – % | 10.9 | 10 | 12.8 | 9.5 |
| History of cardio-/cerebro-vascular disease – % | 30.3 | 36.8 | 23.7 | 19.6 |
| Family history of dementia – % | 55.5 | 53.8 | 55.2 | 64 |
| History of tobacco smoking* – % | 33.7 | 27.3 | 40.3 | 45.4 |
| APOE* – % | | | | |
| ε2ε2 | 0.1 | 0.1 | 0.2 | 0.3 |
| ε2ε3 | 9.7 | 7.9 | 11.5 | 13 |
| ε2ε4 | 2.6 | 2.1 | 3.3 | 2.9 |
| ε3ε3 | 49.2 | 47.5 | 51.2 | 51.3 |
| ε3ε4 | 31.6 | 33.4 | 29.9 | 27.8 |
| ε4ε4 | 6.7 | 8.9 | 3.9 | 4.8 |
| A-beta positivity* – % | 53.3 | 62.2 | 24.2 | 25.8 |
| Tau positivity* – % | 58.6 | 62.9 | 34.1 | 42 |
| Median follow-up – yr | 3.1 (2.0 to 5.6) | 3.1 (2.0 to 5.7) | 3.1 (2.0 to 4.8) | 4.0 (1.7 to 7.8) |
| WM-PVS count | 422 (333 to 526) | 379 (306 to 474) | 481 (388 to 586) | 453 (368 to 539) |
| BG-PVS count | 122 | 104 | 148 | 139 |

| | | | | |
|---|---|---|---|---|
| | (97 to 156) | (87 to 128) | (123 to 173) | (117.75 to 169) |
| WM-PVS diameter – mm | 2.00 (1.90 to 2.12) | 2.08 (1.97 to 2.18) | 1.92 (1.85 to 2.01) | 1.94 (1.86 to 2.04) |
| BG-PVS diameter – mm | 1.60 (1.53 to 1.68) | 1.65 (1.58 to 1.71) | 1.55 (1.51 to 1.60) | 1.56 (1.52 to 1.62) |
| P-WML volume – $mm^3$ | 286 (0 to 1535) | 592 (49 to 1923) | 56 (0 to 842) | 101.50 (0 to 829.25) |
| D-WML volume – $mm^3$ | 25 (3 to 90) | 39 (7 to 110) | 12 (1 to 57.25) | 20.50 (4 to 73.25) |

Plus–minus values are means ± standard deviations, entries with parentheses are medians (interquartile range). Data from three studies — the Alzheimer's Disease Neuroimaging Initiative (ADNI), the National Alzheimer's Coordinating Center (NACC), and the Open Access Series of Imaging Studies (OASIS) — are shown. *Total number of subjects with available history of tobacco smoking, Apolipoprotein E (APOE) genotype, amyloid-beta status and tau status were 3297, 3353, 2118, and 1625, respectively.

**Table S9. Estimates and adjusted significance of the interaction term "marker" by "time" in linear mixed-effects models with the grey matter volume, cortical thickness, or white matter volume as dependent variables.**

| Marker | Grey matter volume ($mm^3$ / year) | Cortical thickness (µm / year) | White matter volume ($mm^3$ / year) |
|---|---|---|---|
| WM-PVS count (per 100-unit) | 709±100, P<0.001 | 4±1, P<0.001 | 591±84, P<0.001 |
| BG-PVS count (per 10-unit) | 187±33, P<0.001 | 1±0, P<0.001 | 142±26, P<0.001 |
| WM-PVS diameter (per 0.1 mm) | -382±84, P<0.001 | -3±1, P<0.001 | -64±67, P=0.675 |
| BG-PVS diameter (per 0.1 mm) | -576±137, P<0.001 | -5±1, P<0.001 | -283±108, P=0.018 |
| P-WML log volume | -288±46, P<0.001 | -2±0, P<0.001 | -73±35, P=0.08 |
| D-WML log volume | -189±62, P=0.004 | -1±0, P=0.01 | -2±48, P=1 |

Estimates indicate the value of the dependent variable preserved (positive values) or lost (negative values) per year for each unit increase in the marker. All models were adjusted for age, sex, race, educational level, body mass index, CDR global score at the baseline, history of diabetes, cardio-/cerebro-vascular disease, hypertension, dyslipidemia, family history of dementia, intracranial volume, the baseline value of the dependent variable (grey matter volume, cortical thickness, or white matter volume), field strength, manufacturer, and intra-individual consistency of the protocol used for the longitudinal MRI acquisitions (consistent versus non-consistent protocol). "P" indicates adjusted P-values.

**Table S10. Regional estimates ($mm^3$/year) and adjusted significance of the interaction term “marker” by “time” in linear mixed-effects models with the volume of the corresponding region as dependent variable.**

| Region | WM-PVS count | BG-PVS count | WM-PVS diameter | BG-PVS diameter | P-WML log volume | D-WML log volume |
|---|---|---|---|---|---|---|
| **lh_bankssts** | 3.3 ± 0.63, P<0.001 | 0.49 ± 0.21, P=1 | -1.52 ± 0.53, P=0.344 | -1.55 ± 0.86, P=1 | -0.87 ± 0.29, P=0.213 | -0.56 ± 0.39, P=1 |
| **lh_caudalanteriorcingulate** | 0.43 ± 0.48, P=1 | -0.05 ± 0.16, P=1 | 0.9 ± 0.4, P=1 | 1.2 ± 0.66, P=1 | -0.14 ± 0.22, P=1 | -0.21 ± 0.3, P=1 |
| **lh_caudalmiddlefrontal** | 4.83 ± 1.31, P=0.019 | 0.93 ± 0.44, P=1 | -0.34 ± 1.09, P=1 | -0.69 ± 1.78, P=1 | -2.19 ± 0.58, P=0.016 | -0.49 ± 0.79, P=1 |
| **lh_cuneus** | 3.19 ± 0.85, P=0.015 | 0.24 ± 0.28, P=1 | 0.15 ± 0.71, P=1 | -0.3 ± 1.16, P=1 | -0.53 ± 0.38, P=1 | -1.15 ± 0.52, P=1 |
| **lh_entorhinal** | 6.49 ± 0.94, P<0.001 | 2.47 ± 0.31, P<0.001 | -3.83 ± 0.79, P<0.001 | -6.76 ± 1.28, P<0.001 | -1.97 ± 0.43, P<0.001 | -0.04 ± 0.58, P=1 |
| **lh_fusiform** | 21.26 ± 2.56, P<0.001 | 7.13 ± 0.84, P<0.001 | -13.66 ± 2.15, P<0.001 | -21.65 ± 3.49, P<0.001 | -6.01 ± 1.18, P<0.001 | -3.74 ± 1.59, P=1 |
| **lh_inferiorparietal** | 19.68 ± 2.93, P<0.001 | 5.85 ± 0.97, P<0.001 | -9.22 ± 2.47, P=0.016 | -17.55 ± 4, P=0.001 | -6.59 ± 1.34, P<0.001 | -4.41 ± 1.81, P=1 |
| **lh_inferiortemporal** | 24.5 ± 3.08, P<0.001 | 8.04 ± 1.01, P<0.001 | -13.43 ± 2.59, P<0.001 | -24.06 ± 4.2, P<0.001 | -7.63 ± 1.41, P<0.001 | -4.67 ± 1.91, P=1 |
| **lh_isthmuscingulate** | 3.78 ± 0.59, P<0.001 | 0.78 ± 0.2, P=0.006 | -0.59 ± 0.5, P=1 | -1.77 ± 0.81, P=1 | -1.14 ± 0.27, P=0.002 | -0.08 ± 0.36, P=1 |
| **lh_lateraloccipital** | 20.63 ± 3.42, P<0.001 | 7.35 ± 1.13, P<0.001 | -12.97 ± 2.86, P=0.001 | -25.33 ± 4.65, P<0.001 | -8.26 ± 1.55, P<0.001 | -6.05 ± 2.1, P=0.34 |
| **lh_lateralorbitofrontal** | 7.41 ± 1.45, P<0.001 | 2.54 ± 0.48, P<0.001 | -1.34 ± 1.21, P=1 | -4.28 ± 1.97, P=1 | -2.59 ± 0.65, P=0.006 | -1.02 ± 0.88, P=1 |
| **lh_lingual** | 8.04 ± 1.76, P<0.001 | 1.89 ± 0.59, P=0.113 | -2.69 ± 1.48, P=1 | -5.02 ± 2.41, P=1 | -1.6 ± 0.8, P=1 | -0.99 ± 1.08, P=1 |
| **lh_medialorbitofrontal** | 6.28 ± 1.49, P=0.002 | 1.56 ± 0.49, P=0.137 | 0.01 ± 1.25, P=1 | -0.58 ± 2.04, P=1 | -2.22 ± 0.68, P=0.096 | -0.66 ± 0.92, P=1 |
| **lh_middletemporal** | 24.18 ± 3.01, P<0.001 | 8.47 ± 0.98, P<0.001 | -14.21 ± 2.53, P<0.001 | -26.78 ± 4.09, P<0.001 | -6.71 ± 1.38, P<0.001 | -3.15 ± 1.87, P=1 |
| **lh_parahippocampal** | 5.58 ± 0.7, P<0.001 | 1.67 ± 0.23, P<0.001 | -2.83 ± 0.59, P<0.001 | -4.63 ± 0.96, P<0.001 | -1.32 ± 0.32, P=0.004 | -0.4 ± 0.43, P=1 |
| **lh_paracentral** | 0.79 ± 0.7, P=1 | -0.13 ± 0.24, P=1 | 0.62 ± 0.59, P=1 | 1.25 ± 0.96, P=1 | -0.96 ± 0.31, P=0.184 | -0.71 ± 0.42, P=1 |
| **lh_parsopercularis** | 5.58 ± 0.97, P<0.001 | 1.29 ± 0.32, P=0.006 | -2.1 ± 0.81, P=0.858 | -2.92 ± 1.33, P=1 | -1.65 ± 0.44, P=0.015 | -0.77 ± 0.59, P=1 |
| **lh_parsorbitalis** | 3 ± 0.69, P=0.001 | 1.14 ± 0.23, P<0.001 | -1.54 ± 0.58, P=0.661 | -3.02 ± 0.94, P=0.116 | -0.66 ± 0.31, P=1 | -0.23 ± 0.42, P=1 |
| **lh_parstriangularis** | 3.32 ± 0.82, P=0.005 | 0.76 ± 0.27, P=0.5 | -0.78 ± 0.69, P=1 | -1.73 ± 1.12, P=1 | -1.03 ± 0.37, P=0.456 | -0.64 ± 0.5, P=1 |
| **lh_pericalcarine** | 0.03 ± 0.73, P=1 | -1.01 ± 0.24, P=0.003 | 2.01 ± 0.6, P=0.076 | 3.25 ± 0.99, P=0.092 | 0.53 ± 0.33, P=1 | -0.03 ± 0.44, P=1 |
| **lh_postcentral** | 5.3 ± 2, P=0.689 | 0.56 ± 0.68, P=1 | 0.34 ± 1.67, P=1 | -1.76 ± 2.73, P=1 | -2.55 ± 0.89, P=0.352 | -1.21 ± 1.2, P=1 |
| **lh_posteriorcingulate** | 2.75 ± 0.72, P=0.013 | 0.55 ± 0.24, P=1 | -0.39 ± 0.61, P=1 | 0.32 ± 0.99, P=1 | -0.67 ± 0.33, P=1 | -0.37 ± 0.44, P=1 |
| **lh_precentral** | 10.19 ± 2.54, P=0.005 | 1.1 ± 0.87, P=1 | -4.1 ± 2.12, P=1 | -6.74 ± 3.48, P=1 | -4.67 ± 1.13, P=0.003 | -2.17 ± 1.53, P=1 |
| **lh_precuneus** | 9.64 ± 1.93, P<0.001 | 2.46 ± 0.65, P=0.013 | -2.24 ± 1.62, P=1 | -3.73 ± 2.64, P=1 | -3.59 ± 0.87, P=0.003 | -2.64 ± 1.18, P=1 |
| **lh_rostralanteriorcingulate** | 1.39 ± 0.59, P=1 | 0.7 ± 0.2, P=0.032 | -0.59 ± 0.5, P=1 | -1.48 ± 0.81, P=1 | -0.3 ± 0.27, P=1 | -0.68 ± 0.36, P=1 |
| **lh_rostralmiddlefrontal** | 10.25 ± 3.04, P=0.065 | 1.23 ± 1.03, P=1 | -0.89 ± 2.54, P=1 | 1.21 ± 4.15, P=1 | -4.65 ± 1.36, P=0.053 | -2.48 ± 1.84, P=1 |
| **lh_superiorfrontal** | 12.01 ± 4.28, P=0.437 | 3.73 ± 1.44, P=0.831 | 1.03 ± 3.58, P=1 | -2.23 ± 5.84, P=1 | -5.35 ± 1.92, P=0.466 | -3.49 ± 2.6, P=1 |
| **lh_superiorparietal** | 10.82 ± 2.99, P=0.026 | 3.08 ± 1, P=0.19 | -3.55 ± 2.5, P=1 | -6.17 ± 4.08, P=1 | -6 ± 1.34, P=0.001 | -4.91 ± 1.82, P=0.595 |
| **lh_superiortemporal** | 24.98 ± 2.96, P<0.001 | 7.78 ± 0.98, P<0.001 | -13.58 ± 2.49, P<0.001 | -23.93 ± 4.04, P<0.001 | -7.22 ± 1.35, P<0.001 | -2.82 ± 1.84, P=1 |
| **lh_supramarginal** | 17.13 ± 2.57, P<0.001 | 3.48 ± 0.86, P=0.005 | -4.76 ± 2.16, P=1 | -8.85 ± 3.52, P=1 | -6.68 ± 1.16, P<0.001 | -2.12 ± 1.58, P=1 |

| | | | | | | |
|---|---|---|---|---|---|---|
| **lh_frontalpole** | 2.07 ± 0.47, P=0.001 | 0.76 ± 0.16, P<0.001 | -1.01 ± 0.4, P=0.937 | -1.56 ± 0.64, P=1 | -0.22 ± 0.22, P=1 | -0.46 ± 0.29, P=1 |
| **lh_temporalpole** | 7.68 ± 1.22, P<0.001 | 2.19 ± 0.4, P<0.001 | -3.33 ± 1.03, P=0.102 | -6.46 ± 1.66, P=0.009 | -1.85 ± 0.56, P=0.083 | -0.3 ± 0.76, P=1 |
| **lh_transversetemporal** | 1.71 ± 0.33, P<0.001 | 0.34 ± 0.11, P=0.157 | -1.1 ± 0.27, P=0.006 | -1.47 ± 0.45, P=0.088 | -0.49 ± 0.15, P=0.076 | -0.87 ± 0.2, P=0.001 |
| **lh_insula** | 8.98 ± 1.56, P<0.001 | 2.11 ± 0.51, P=0.004 | -2.72 ± 1.31, P=1 | -3.09 ± 2.13, P=1 | -3.15 ± 0.71, P=0.001 | -1.13 ± 0.96, P=1 |
| **rh_bankssts** | 2.9 ± 0.51, P<0.001 | 0.85 ± 0.17, P<0.001 | -2 ± 0.42, P<0.001 | -2.91 ± 0.69, P=0.002 | -1.09 ± 0.23, P<0.001 | -0.98 ± 0.31, P=0.131 |
| **rh_caudalanteriorcingulate** | 1.05 ± 0.64, P=1 | -0.02 ± 0.21, P=1 | 0 ± 0.54, P=1 | 1 ± 0.87, P=1 | -0.21 ± 0.29, P=1 | -0.09 ± 0.39, P=1 |
| **rh_caudalmiddlefrontal** | 4.31 ± 1.24, P=0.046 | 0.81 ± 0.42, P=1 | -0.9 ± 1.04, P=1 | -1.34 ± 1.7, P=1 | -2.69 ± 0.56, P<0.001 | -1.24 ± 0.75, P=1 |
| **rh_cuneus** | 3.39 ± 0.94, P=0.026 | 1.13 ± 0.31, P=0.027 | -1.03 ± 0.78, P=1 | -2.8 ± 1.27, P=1 | -1.36 ± 0.42, P=0.109 | -1.82 ± 0.57, P=0.123 |
| **rh_entorhinal** | 5.68 ± 0.9, P<0.001 | 2.42 ± 0.29, P<0.001 | -3.46 ± 0.76, P<0.001 | -7.42 ± 1.22, P<0.001 | -1.57 ± 0.41, P=0.012 | -1.28 ± 0.56, P=1 |
| **rh_fusiform** | 18.26 ± 2.39, P<0.001 | 7.02 ± 0.78, P<0.001 | -11.15 ± 2.01, P<0.001 | -20.61 ± 3.25, P<0.001 | -7.09 ± 1.09, P<0.001 | -4.93 ± 1.48, P=0.073 |
| **rh_inferiorparietal** | 17.91 ± 3.32, P<0.001 | 5.6 ± 1.11, P<0.001 | -7.45 ± 2.79, P=0.652 | -17.12 ± 4.52, P=0.014 | -7.36 ± 1.5, P<0.001 | -4.76 ± 2.03, P=1 |
| **rh_inferiortemporal** | 22.17 ± 2.92, P<0.001 | 7.42 ± 0.96, P<0.001 | -12.12 ± 2.46, P<0.001 | -21.49 ± 3.99, P<0.001 | -8.36 ± 1.33, P<0.001 | -4.53 ± 1.81, P=1 |
| **rh_isthmuscingulate** | 2.82 ± 0.65, P=0.001 | 0.9 ± 0.22, P=0.003 | -0.42 ± 0.54, P=1 | -1.4 ± 0.89, P=1 | -0.92 ± 0.29, P=0.154 | -0.45 ± 0.4, P=1 |
| **rh_lateraloccipital** | 18.44 ± 3.44, P<0.001 | 4.96 ± 1.15, P=0.001 | -9.21 ± 2.89, P=0.125 | -16.17 ± 4.69, P=0.05 | -9.18 ± 1.56, P<0.001 | -6.2 ± 2.1, P=0.281 |
| **rh_lateralorbitofrontal** | 10.67 ± 1.66, P<0.001 | 3.03 ± 0.55, P<0.001 | -3.22 ± 1.4, P=1 | -7.24 ± 2.27, P=0.123 | -3.5 ± 0.75, P<0.001 | -0.4 ± 1.02, P=1 |
| **rh_lingual** | 8.67 ± 1.8, P<0.001 | 3.26 ± 0.6, P<0.001 | -3.26 ± 1.51, P=1 | -6.57 ± 2.45, P=0.645 | -3.54 ± 0.81, P=0.001 | -2.14 ± 1.1, P=1 |
| **rh_medialorbitofrontal** | 6.02 ± 1.52, P=0.007 | 2.37 ± 0.5, P<0.001 | -2.23 ± 1.27, P=1 | -4.19 ± 2.07, P=1 | -2.68 ± 0.69, P=0.009 | -1.79 ± 0.93, P=1 |
| **rh_middletemporal** | 26.71 ± 3.3, P<0.001 | 9.94 ± 1.08, P<0.001 | -18.26 ± 2.77, P<0.001 | -31.5 ± 4.48, P<0.001 | -8.87 ± 1.51, P<0.001 | -4.03 ± 2.04, P=1 |
| **rh_parahippocampal** | 4.59 ± 0.68, P<0.001 | 1.84 ± 0.22, P<0.001 | -2.63 ± 0.57, P<0.001 | -5.29 ± 0.92, P<0.001 | -1.46 ± 0.31, P<0.001 | -0.79 ± 0.42, P=1 |
| **rh_paracentral** | 0.42 ± 0.86, P=1 | -0.51 ± 0.29, P=1 | 0.41 ± 0.72, P=1 | 2.35 ± 1.17, P=1 | -0.64 ± 0.38, P=1 | -0.82 ± 0.52, P=1 |
| **rh_parsopercularis** | 5.6 ± 0.92, P<0.001 | 1.89 ± 0.3, P<0.001 | -3.04 ± 0.77, P=0.007 | -5.7 ± 1.25, P<0.001 | -2.2 ± 0.42, P<0.001 | -0.6 ± 0.56, P=1 |
| **rh_parsorbitalis** | 4.27 ± 0.89, P<0.001 | 2.17 ± 0.29, P<0.001 | -3.37 ± 0.74, P=0.001 | -6.75 ± 1.2, P<0.001 | -1.44 ± 0.4, P=0.032 | -1.03 ± 0.55, P=1 |
| **rh_parstriangularis** | 4.35 ± 1, P=0.001 | 1.99 ± 0.33, P<0.001 | -3.44 ± 0.83, P=0.003 | -6.29 ± 1.35, P<0.001 | -2.09 ± 0.45, P<0.001 | -1.04 ± 0.61, P=1 |
| **rh_pericalcarine** | 0.93 ± 0.83, P=1 | -1.07 ± 0.28, P=0.01 | 1.85 ± 0.69, P=0.647 | 3.44 ± 1.13, P=0.2 | -0.15 ± 0.37, P=1 | 0 ± 0.5, P=1 |
| **rh_postcentral** | 2.4 ± 2.03, P=1 | 0.43 ± 0.69, P=1 | 0.97 ± 1.69, P=1 | -0.06 ± 2.77, P=1 | -1.58 ± 0.91, P=1 | -1.65 ± 1.23, P=1 |
| **rh_posteriorcingulate** | 3.02 ± 0.77, P=0.007 | 1.09 ± 0.25, P=0.002 | -0.66 ± 0.64, P=1 | -1.21 ± 1.05, P=1 | -1.05 ± 0.35, P=0.228 | -0.5 ± 0.47, P=1 |
| **rh_precentral** | 8.97 ± 2.56, P=0.04 | 2.14 ± 0.88, P=1 | -5.21 ± 2.13, P=1 | -9.12 ± 3.5, P=0.795 | -4.91 ± 1.13, P=0.001 | -3.43 ± 1.54, P=1 |
| **rh_precuneus** | 8.95 ± 1.97, P=0.001 | 2.55 ± 0.66, P=0.01 | -3.99 ± 1.65, P=1 | -5.67 ± 2.69, P=1 | -4.38 ± 0.89, P<0.001 | -3.95 ± 1.2, P=0.09 |
| **rh_rostralanteriorcingulate** | 2.25 ± 0.51, P=0.001 | 0.36 ± 0.17, P=1 | -0.35 ± 0.43, P=1 | -0.22 ± 0.7, P=1 | -0.57 ± 0.23, P=1 | -0.19 ± 0.31, P=1 |
| **rh_rostralmiddlefrontal** | 12.57 ± 3.19, P=0.007 | 5.41 ± 1.07, P<0.001 | -6.83 ± 2.67, P=0.921 | -13.38 ± 4.35, P=0.184 | -7.04 ± 1.43, P<0.001 | -4.87 ± 1.94, P=1 |
| **rh_superiorfrontal** | 11.89 ± 4.45, P=0.654 | 3.35 ± 1.49, P=1 | 1.65 ± 3.72, P=1 | -0.25 ± 6.07, P=1 | -7.76 ± 2, P=0.009 | -3.85 ± 2.7, P=1 |
| **rh_superiorparietal** | 9.03 ± 2.95, P=0.195 | 2.26 ± 0.99, P=1 | -0.74 ± 2.47, P=1 | -3.58 ± 4.02, P=1 | -5.43 ± 1.32, P=0.004 | -5.08 ± 1.79, P=0.395 |
| **rh_superiortemporal** | 24.55 ± 2.96, P<0.001 | 9.99 ± 0.97, P<0.001 | -18.43 ± 2.48, P<0.001 | -31.12 ± 4.02, P<0.001 | -8.92 ± 1.35, P<0.001 | -5.29 ± 1.83, P=0.34 |
| **rh_supramarginal** | 13.95 ± 2.2, P<0.001 | 4.96 ± 0.73, P<0.001 | -8.98 ± 1.85, P<0.001 | -16.7 ± 3, P<0.001 | -5.36 ± 1, P<0.001 | -2.46 ± 1.35, P=1 |

| | | | | | | |
|---|---|---|---|---|---|---|
| **rh_frontalpole** | 3.01 ± 0.62, P<0.001 | 1.22 ± 0.2, P<0.001 | -2.15 ± 0.52, P=0.003 | -3.34 ± 0.85, P=0.007 | -1.3 ± 0.28, P<0.001 | -1.44 ± 0.38, P=0.014 |
| **rh_temporalpole** | 6.78 ± 1.27, P<0.001 | 2.7 ± 0.41, P<0.001 | -4.87 ± 1.06, P<0.001 | -8.03 ± 1.73, P<0.001 | -2.87 ± 0.58, P<0.001 | -2.44 ± 0.78, P=0.161 |
| **rh_transversetemporal** | 1.67 ± 0.28, P<0.001 | 0.58 ± 0.09, P<0.001 | -0.84 ± 0.23, P=0.028 | -1.94 ± 0.38, P<0.001 | -0.45 ± 0.13, P=0.032 | -0.54 ± 0.17, P=0.122 |
| **rh_insula** | 9.11 ± 1.55, P<0.001 | 2.83 ± 0.51, P<0.001 | -3.75 ± 1.3, P=0.342 | -6.26 ± 2.11, P=0.266 | -3.52 ± 0.71, P<0.001 | -0.9 ± 0.96, P=1 |
| **Left-Thalamus-Proper** | -1.49 ± 1.27, P=1 | -0.49 ± 0.42, P=1 | 0.29 ± 1.06, P=1 | 2.46 ± 1.73, P=1 | -1.01 ± 0.57, P=1 | -1.13 ± 0.77, P=1 |
| **Right-Thalamus-Proper** | 1.04 ± 1.29, P=1 | -0.06 ± 0.43, P=1 | -0.97 ± 1.08, P=1 | 1.25 ± 1.76, P=1 | -1.36 ± 0.59, P=1 | -1.18 ± 0.79, P=1 |
| **Right-Lateral-Ventricle** | -117.67 ± 13.2, P<0.001 | -34.94 ± 4.29, P<0.001 | 55.56 ± 11.12, P<0.001 | 92.96 ± 18.04, P<0.001 | 47.6 ± 6.1, P<0.001 | 9.66 ± 8.23, P=1 |
| **Left-Hippocampus** | 2.49 ± 0.88, P=0.43 | 1.88 ± 0.29, P<0.001 | -4.12 ± 0.74, P<0.001 | -6.63 ± 1.19, P<0.001 | -0.55 ± 0.4, P=1 | -1.81 ± 0.54, P=0.074 |
| **Left-Lateral-Ventricle** | -112.15 ± 15.48, P<0.001 | -27.37 ± 5.03, P<0.001 | 37.26 ± 13.03, P=0.367 | 63.74 ± 21.14, P=0.223 | 44.61 ± 7.15, P<0.001 | -2.75 ± 9.62, P=1 |
| **Right-Putamen** | 2.98 ± 1.2, P=1 | 0.36 ± 0.4, P=1 | -2.2 ± 1, P=1 | -0.15 ± 1.64, P=1 | -0.76 ± 0.55, P=1 | -0.38 ± 0.74, P=1 |
| **Right-Amygdala** | 1.01 ± 0.42, P=1 | 0.33 ± 0.14, P=1 | -1.81 ± 0.35, P<0.001 | -1.62 ± 0.58, P=0.438 | -0.33 ± 0.19, P=1 | -1.02 ± 0.26, P=0.007 |
| **Left-Putamen** | 1.1 ± 1.2, P=1 | -0.4 ± 0.4, P=1 | -1.69 ± 1.01, P=1 | 3.47 ± 1.64, P=1 | 0.04 ± 0.55, P=1 | -0.61 ± 0.74, P=1 |
| **Right-VentralDC** | -1.25 ± 0.57, P=1 | 0.29 ± 0.2, P=1 | -0.47 ± 0.47, P=1 | -0.45 ± 0.78, P=1 | 0.08 ± 0.25, P=1 | -0.38 ± 0.33, P=1 |
| **Left-VentralDC** | -2.39 ± 0.56, P=0.002 | 0.28 ± 0.2, P=1 | 0.04 ± 0.46, P=1 | -0.38 ± 0.77, P=1 | 0.24 ± 0.24, P=1 | -0.48 ± 0.32, P=1 |
| **Right-Hippocampus** | 3.17 ± 0.91, P=0.046 | 2.16 ± 0.3, P<0.001 | -3.95 ± 0.76, P<0.001 | -7.34 ± 1.24, P<0.001 | -0.69 ± 0.42, P=1 | -1.88 ± 0.56, P=0.071 |
| **Left-Amygdala** | 0.63 ± 0.39, P=1 | 0.18 ± 0.13, P=1 | -1.13 ± 0.33, P=0.048 | -1.57 ± 0.53, P=0.282 | -0.2 ± 0.18, P=1 | -0.52 ± 0.24, P=1 |
| **Left-Pallidum** | -0.84 ± 0.45, P=1 | 0.25 ± 0.15, P=1 | -0.21 ± 0.38, P=1 | -1.2 ± 0.62, P=1 | 0.27 ± 0.2, P=1 | 0.1 ± 0.27, P=1 |
| **Right-Pallidum** | -1.3 ± 0.45, P=0.296 | 0.33 ± 0.15, P=1 | -0.29 ± 0.37, P=1 | -1.1 ± 0.61, P=1 | 0.61 ± 0.2, P=0.215 | 0.13 ± 0.27, P=1 |
| **Left-Caudate** | 1.48 ± 0.96, P=1 | -0.55 ± 0.32, P=1 | -0.01 ± 0.8, P=1 | 5.32 ± 1.3, P=0.004 | -0.11 ± 0.43, P=1 | -0.07 ± 0.58, P=1 |
| **Right-Caudate** | -0.33 ± 1.02, P=1 | -0.52 ± 0.34, P=1 | 0.48 ± 0.85, P=1 | 5.27 ± 1.38, P=0.012 | 0.49 ± 0.46, P=1 | -0.27 ± 0.62, P=1 |
| **3rd-ventricle** | 0.1 ± 0.79, P=1 | -1.28 ± 0.26, P<0.001 | 2.04 ± 0.66, P=0.163 | 3.54 ± 1.07, P=0.079 | 0.7 ± 0.36, P=1 | 0.93 ± 0.48, P=1 |
| **4th-ventricle** | -0.37 ± 0.87, P=1 | -1.18 ± 0.29, P=0.004 | 0.99 ± 0.72, P=1 | 1.84 ± 1.18, P=1 | 1.46 ± 0.39, P=0.016 | 0.04 ± 0.53, P=1 |

The estimates indicate the grey matter or ventricular volume for that region preserved (positive values) or lost (negative values) per year for each unit increase in the marker assessed at the baseline. Units were 100 for WM-PVS count, 10 for BG-PVS count, 0.1 mm for WM- and BG-PVS diameter, and 1 log for P-WML and D-WML volume. See also Figure 2A. Parcellations performed by Freesurfer according to the Desikan-Killany atlas. Bankssts: Banks of the Superior Temporal Sulcus; DC: diencephalon. Lh: left hemisphere; Rh: right hemisphere. Models were fully adjusted for all the same covariates included in the main linear mixed-effect model. P values are corrected for 86 comparisons with Holm-Bonferroni procedure.

**Table S11. Regional estimates (µm/year) and adjusted significance of the interaction term "marker" by "time" in linear mixed-effects models with the thickness of the corresponding region as dependent variable.**

| Region | WM-PVS count | BG-PVS count | WM-PVS diameter | BG-PVS diameter | P-WML log volume | D-WML log volume |
|---|---|---|---|---|---|---|
| **lh_bankssts** | 2.89 ± 0.49, P<0.001 | 0.98 ± 0.17, P<0.001 | -2.22 ± 0.42, P<0.001 | -3.44 ± 0.69, P<0.001 | -0.69 ± 0.23, P=0.148 | -0.56 ± 0.31, P=1 |
| **lh_caudalanteriorcingulate** | 2.05 ± 0.54, P=0.012 | 0.64 ± 0.18, P=0.032 | -0.7 ± 0.47, P=1 | -0.79 ± 0.76, P=1 | -0.75 ± 0.25, P=0.175 | -0.07 ± 0.34, P=1 |
| **lh_caudalmiddlefrontal** | 1.61 ± 0.41, P=0.007 | 0.66 ± 0.14, P<0.001 | -0.81 ± 0.35, P=1 | -1.69 ± 0.58, P=0.247 | -0.77 ± 0.18, P=0.002 | -0.23 ± 0.25, P=1 |
| **lh_cuneus** | 0.49 ± 0.31, P=1 | -0.05 ± 0.11, P=1 | 0.22 ± 0.26, P=1 | 0 ± 0.43, P=1 | -0.2 ± 0.14, P=1 | -0.45 ± 0.19, P=1 |
| **lh_entorhinal** | 5.79 ± 0.95, P<0.001 | 2.97 ± 0.31, P<0.001 | -4.7 ± 0.81, P<0.001 | -8.9 ± 1.32, P<0.001 | -1.92 ± 0.44, P=0.001 | -1.16 ± 0.6, P=1 |
| **lh_frontalpole** | 1.3 ± 0.64, P=1 | 1.12 ± 0.22, P<0.001 | -0.72 ± 0.54, P=1 | -3 ± 0.89, P=0.052 | -0.12 ± 0.29, P=1 | -0.17 ± 0.39, P=1 |
| **lh_fusiform** | 3.79 ± 0.52, P<0.001 | 1.64 ± 0.17, P<0.001 | -3 ± 0.45, P<0.001 | -5.42 ± 0.73, P<0.001 | -1.24 ± 0.24, P<0.001 | -0.65 ± 0.33, P=1 |
| **lh_inferiorparietal** | 2.34 ± 0.43, P<0.001 | 0.8 ± 0.14, P<0.001 | -1.56 ± 0.37, P=0.002 | -2.69 ± 0.6, P<0.001 | -0.92 ± 0.19, P<0.001 | -0.64 ± 0.27, P=1 |
| **lh_inferiortemporal** | 3.29 ± 0.52, P<0.001 | 1.54 ± 0.17, P<0.001 | -2.25 ± 0.45, P<0.001 | -4.57 ± 0.73, P<0.001 | -0.95 ± 0.24, P=0.005 | -0.57 ± 0.33, P=1 |
| **lh_insula** | 2.26 ± 0.56, P=0.004 | 0.89 ± 0.19, P<0.001 | -1.03 ± 0.48, P=1 | -2.04 ± 0.78, P=0.645 | -0.59 ± 0.26, P=1 | -0.26 ± 0.35, P=1 |
| **lh_isthmuscingulate** | 1.74 ± 0.38, P<0.001 | 0.64 ± 0.13, P<0.001 | -0.73 ± 0.32, P=1 | -1.87 ± 0.53, P=0.029 | -0.52 ± 0.17, P=0.164 | -0.17 ± 0.24, P=1 |
| **lh_lateraloccipital** | 1.41 ± 0.36, P=0.007 | 0.59 ± 0.12, P<0.001 | -0.85 ± 0.31, P=0.414 | -2.04 ± 0.5, P=0.004 | -0.66 ± 0.16, P=0.003 | -0.44 ± 0.22, P=1 |
| **lh_lateralorbitofrontal** | 1.66 ± 0.42, P=0.006 | 0.93 ± 0.14, P<0.001 | -0.74 ± 0.36, P=1 | -1.8 ± 0.58, P=0.143 | -0.63 ± 0.19, P=0.06 | -0.15 ± 0.26, P=1 |
| **lh_lingual** | 0.64 ± 0.34, P=1 | 0.22 ± 0.12, P=1 | -0.17 ± 0.29, P=1 | -0.68 ± 0.47, P=1 | -0.21 ± 0.15, P=1 | -0.24 ± 0.21, P=1 |
| **lh_medialorbitofrontal** | 1.53 ± 0.43, P=0.03 | 0.74 ± 0.15, P<0.001 | -0.49 ± 0.37, P=1 | -0.97 ± 0.6, P=1 | -0.58 ± 0.2, P=0.239 | -0.29 ± 0.27, P=1 |
| **lh_middletemporal** | 3.73 ± 0.53, P<0.001 | 1.43 ± 0.18, P<0.001 | -2.27 ± 0.46, P<0.001 | -4.34 ± 0.74, P<0.001 | -1.05 ± 0.24, P=0.001 | -0.49 ± 0.34, P=1 |
| **lh_paracentral** | 0.63 ± 0.42, P=1 | 0.37 ± 0.15, P=0.872 | -0.68 ± 0.36, P=1 | -1.09 ± 0.59, P=1 | -0.52 ± 0.19, P=0.383 | -0.55 ± 0.26, P=1 |
| **lh_parahippocampal** | 4.77 ± 0.71, P<0.001 | 1.7 ± 0.24, P<0.001 | -2.72 ± 0.61, P=0.001 | -4.98 ± 0.99, P<0.001 | -1.55 ± 0.32, P<0.001 | -0.86 ± 0.44, P=1 |
| **lh_parsopercularis** | 1.91 ± 0.39, P<0.001 | 0.68 ± 0.13, P<0.001 | -1.02 ± 0.33, P=0.138 | -1.78 ± 0.54, P=0.069 | -0.76 ± 0.17, P=0.001 | -0.35 ± 0.24, P=1 |
| **lh_parsorbitalis** | 0.81 ± 0.49, P=1 | 0.71 ± 0.17, P=0.002 | -0.49 ± 0.42, P=1 | -1.47 ± 0.69, P=1 | -0.22 ± 0.22, P=1 | 0.17 ± 0.3, P=1 |
| **lh_parstriangularis** | 1.01 ± 0.36, P=0.338 | 0.44 ± 0.12, P=0.023 | -0.29 ± 0.3, P=1 | -0.87 ± 0.5, P=1 | -0.39 ± 0.16, P=1 | -0.39 ± 0.22, P=1 |
| **lh_pericalcarine** | -0.43 ± 0.34, P=1 | -0.25 ± 0.12, P=1 | 0.79 ± 0.29, P=0.493 | 0.64 ± 0.48, P=1 | 0.07 ± 0.15, P=1 | -0.14 ± 0.21, P=1 |
| **lh_postcentral** | 0.93 ± 0.33, P=0.323 | 0.33 ± 0.11, P=0.239 | -0.39 ± 0.28, P=1 | -1.42 ± 0.46, P=0.138 | -0.41 ± 0.15, P=0.391 | -0.44 ± 0.2, P=1 |
| **lh_posteriorcingulate** | 1.76 ± 0.38, P<0.001 | 0.64 ± 0.13, P<0.001 | -0.78 ± 0.32, P=1 | -1.22 ± 0.52, P=1 | -0.77 ± 0.17, P<0.001 | -0.17 ± 0.23, P=1 |
| **lh_precentral** | 2.07 ± 0.42, P<0.001 | 0.79 ± 0.15, P<0.001 | -1.49 ± 0.36, P=0.002 | -3 ± 0.59, P<0.001 | -0.82 ± 0.19, P=0.001 | -0.58 ± 0.26, P=1 |
| **lh_precuneus** | 1.49 ± 0.37, P=0.004 | 0.67 ± 0.12, P<0.001 | -1.14 ± 0.31, P=0.02 | -1.96 ± 0.51, P=0.009 | -0.66 ± 0.17, P=0.005 | -0.63 ± 0.23, P=0.383 |
| **lh_rostralanteriorcingulate** | 0.63 ± 0.46, P=1 | 0.62 ± 0.16, P=0.005 | -0.65 ± 0.4, P=1 | -1.72 ± 0.64, P=0.538 | -0.35 ± 0.21, P=1 | -0.05 ± 0.29, P=1 |
| **lh_rostralmiddlefrontal** | 0.59 ± 0.34, P=1 | 0.15 ± 0.12, P=1 | 0 ± 0.29, P=1 | 0.11 ± 0.48, P=1 | -0.38 ± 0.15, P=0.9 | -0.24 ± 0.21, P=1 |
| **lh_superiorfrontal** | 0.83 ± 0.39, P=1 | 0.61 ± 0.13, P<0.001 | -0.39 ± 0.33, P=1 | -1.19 ± 0.54, P=1 | -0.53 ± 0.17, P=0.161 | -0.25 ± 0.24, P=1 |
| **lh_superiorparietal** | 1.37 ± 0.35, P=0.008 | 0.51 ± 0.12, P=0.002 | -0.93 ± 0.3, P=0.135 | -1.68 ± 0.49, P=0.046 | -0.73 ± 0.16, P<0.001 | -0.63 ± 0.22, P=0.265 |

| | | | | | | |
|---|---|---|---|---|---|---|
| **lh_superiortemporal** | 3.5 ± 0.49, P<0.001 | 1.43 ± 0.16, P<0.001 | -2.72 ± 0.42, P<0.001 | -4.95 ± 0.68, P<0.001 | -1.14 ± 0.23, P<0.001 | -0.58 ± 0.31, P=1 |
| **lh_supramarginal** | 2.13 ± 0.39, P<0.001 | 0.71 ± 0.13, P<0.001 | -1.29 ± 0.33, P=0.006 | -2.57 ± 0.54, P<0.001 | -0.96 ± 0.17, P<0.001 | -0.41 ± 0.24, P=1 |
| **lh_temporalpole** | 3.86 ± 0.92, P=0.002 | 2.14 ± 0.31, P<0.001 | -2.82 ± 0.79, P=0.026 | -6.39 ± 1.28, P<0.001 | -1.61 ± 0.42, P=0.009 | -0.55 ± 0.58, P=1 |
| **lh_transversetemporal** | 3.43 ± 0.51, P<0.001 | 1.01 ± 0.18, P<0.001 | -2.57 ± 0.44, P<0.001 | -4.55 ± 0.71, P<0.001 | -1.13 ± 0.23, P<0.001 | -1.38 ± 0.32, P=0.001 |
| **rh_bankssts** | 2.7 ± 0.5, P<0.001 | 1.32 ± 0.17, P<0.001 | -2.56 ± 0.43, P<0.001 | -4.42 ± 0.7, P<0.001 | -0.92 ± 0.23, P=0.004 | -0.86 ± 0.31, P=0.433 |
| **rh_caudalanteriorcingulate** | 0.44 ± 0.58, P=1 | 0.15 ± 0.19, P=1 | -0.07 ± 0.49, P=1 | 0.02 ± 0.8, P=1 | -0.38 ± 0.26, P=1 | -0.09 ± 0.36, P=1 |
| **rh_caudalmiddlefrontal** | 1.75 ± 0.42, P=0.002 | 0.67 ± 0.15, P<0.001 | -1.37 ± 0.36, P=0.01 | -2.27 ± 0.59, P=0.009 | -0.97 ± 0.19, P<0.001 | -0.68 ± 0.26, P=0.617 |
| **rh_cuneus** | 0.38 ± 0.34, P=1 | 0.24 ± 0.12, P=1 | -0.3 ± 0.29, P=1 | -0.94 ± 0.48, P=1 | -0.29 ± 0.15, P=1 | -0.73 ± 0.21, P=0.035 |
| **rh_entorhinal** | 5.76 ± 1.01, P<0.001 | 2.81 ± 0.34, P<0.001 | -4.15 ± 0.87, P<0.001 | -9.28 ± 1.41, P<0.001 | -1.97 ± 0.46, P=0.002 | -1.56 ± 0.64, P=1 |
| **rh_frontalpole** | 0.97 ± 0.69, P=1 | 0.97 ± 0.23, P=0.003 | -0.52 ± 0.59, P=1 | -3.24 ± 0.95, P=0.049 | -0.53 ± 0.31, P=1 | -0.52 ± 0.42, P=1 |
| **rh_fusiform** | 3.38 ± 0.54, P<0.001 | 1.77 ± 0.18, P<0.001 | -2.8 ± 0.46, P<0.001 | -5.67 ± 0.75, P<0.001 | -1.31 ± 0.25, P<0.001 | -0.98 ± 0.34, P=0.272 |
| **rh_inferiorparietal** | 1.65 ± 0.41, P=0.004 | 0.83 ± 0.14, P<0.001 | -0.85 ± 0.35, P=1 | -2.41 ± 0.57, P=0.002 | -0.87 ± 0.18, P<0.001 | -0.71 ± 0.25, P=0.339 |
| **rh_inferiortemporal** | 3.09 ± 0.51, P<0.001 | 1.53 ± 0.17, P<0.001 | -2.27 ± 0.44, P<0.001 | -4.81 ± 0.71, P<0.001 | -1.29 ± 0.23, P<0.001 | -0.9 ± 0.32, P=0.347 |
| **rh_insula** | 2 ± 0.58, P=0.043 | 1.28 ± 0.19, P<0.001 | -1.8 ± 0.5, P=0.022 | -3.53 ± 0.81, P=0.001 | -0.89 ± 0.27, P=0.063 | -0.48 ± 0.37, P=1 |
| **rh_isthmuscingulate** | 1.86 ± 0.42, P=0.001 | 0.75 ± 0.14, P<0.001 | -1.01 ± 0.36, P=0.392 | -2.3 ± 0.59, P=0.007 | -0.59 ± 0.19, P=0.169 | -0.19 ± 0.26, P=1 |
| **rh_lateraloccipital** | 0.84 ± 0.37, P=1 | 0.44 ± 0.13, P=0.03 | -0.44 ± 0.31, P=1 | -1.63 ± 0.51, P=0.109 | -0.56 ± 0.17, P=0.056 | -0.76 ± 0.23, P=0.06 |
| **rh_lateralorbitofrontal** | 2.17 ± 0.46, P<0.001 | 1.14 ± 0.15, P<0.001 | -1.71 ± 0.4, P=0.001 | -3.61 ± 0.64, P<0.001 | -0.89 ± 0.21, P=0.002 | -0.31 ± 0.29, P=1 |
| **rh_lingual** | 0.71 ± 0.36, P=1 | 0.51 ± 0.12, P=0.002 | -0.79 ± 0.31, P=0.751 | -1.63 ± 0.5, P=0.083 | -0.32 ± 0.16, P=1 | -0.41 ± 0.22, P=1 |
| **rh_medialorbitofrontal** | 1.19 ± 0.47, P=0.807 | 0.65 ± 0.16, P=0.003 | -0.57 ± 0.4, P=1 | -1.37 ± 0.65, P=1 | -0.66 ± 0.21, P=0.141 | -0.58 ± 0.29, P=1 |
| **rh_middletemporal** | 3.14 ± 0.52, P<0.001 | 1.55 ± 0.17, P<0.001 | -2.46 ± 0.44, P<0.001 | -4.97 ± 0.71, P<0.001 | -1.04 ± 0.24, P=0.001 | -0.75 ± 0.32, P=1 |
| **rh_paracentral** | 0.82 ± 0.44, P=1 | 0.32 ± 0.15, P=1 | -0.7 ± 0.37, P=1 | -0.86 ± 0.61, P=1 | -0.49 ± 0.19, P=0.808 | -0.52 ± 0.27, P=1 |
| **rh_parahippocampal** | 4.24 ± 0.71, P<0.001 | 1.93 ± 0.24, P<0.001 | -2.94 ± 0.61, P<0.001 | -5.91 ± 0.99, P<0.001 | -1.66 ± 0.33, P<0.001 | -1.17 ± 0.45, P=0.654 |
| **rh_parsopercularis** | 2.07 ± 0.4, P<0.001 | 0.87 ± 0.14, P<0.001 | -1.59 ± 0.34, P<0.001 | -2.93 ± 0.56, P<0.001 | -0.87 ± 0.18, P<0.001 | -0.35 ± 0.25, P=1 |
| **rh_parsorbitalis** | 0.94 ± 0.49, P=1 | 1.09 ± 0.17, P<0.001 | -1.14 ± 0.42, P=0.485 | -3.47 ± 0.68, P<0.001 | -0.57 ± 0.22, P=0.784 | -0.46 ± 0.31, P=1 |
| **rh_parstriangularis** | 1.23 ± 0.37, P=0.057 | 0.62 ± 0.13, P<0.001 | -1.23 ± 0.31, P=0.006 | -2.1 ± 0.51, P=0.003 | -0.59 ± 0.16, P=0.025 | -0.53 ± 0.23, P=1 |
| **rh_pericalcarine** | -0.39 ± 0.35, P=1 | -0.23 ± 0.12, P=1 | 0.23 ± 0.3, P=1 | 0.21 ± 0.49, P=1 | 0 ± 0.16, P=1 | -0.49 ± 0.21, P=1 |
| **rh_postcentral** | 0.56 ± 0.34, P=1 | 0.38 ± 0.12, P=0.09 | -0.3 ± 0.29, P=1 | -1.31 ± 0.47, P=0.395 | -0.36 ± 0.15, P=1 | -0.44 ± 0.21, P=1 |
| **rh_posteriorcingulate** | 2.18 ± 0.43, P<0.001 | 0.79 ± 0.14, P<0.001 | -0.88 ± 0.37, P=1 | -1.66 ± 0.59, P=0.364 | -0.83 ± 0.19, P=0.001 | -0.01 ± 0.27, P=1 |
| **rh_precentral** | 1.97 ± 0.43, P<0.001 | 0.87 ± 0.15, P<0.001 | -1.63 ± 0.36, P=0.001 | -3.18 ± 0.6, P<0.001 | -0.89 ± 0.19, P<0.001 | -0.62 ± 0.26, P=1 |
| **rh_precuneus** | 1.65 ± 0.36, P<0.001 | 0.66 ± 0.12, P<0.001 | -1.23 ± 0.31, P=0.004 | -2.19 ± 0.5, P=0.001 | -0.73 ± 0.16, P<0.001 | -0.81 ± 0.22, P=0.019 |
| **rh_rostralanteriorcingulate** | 2.07 ± 0.52, P=0.006 | 0.52 ± 0.18, P=0.215 | -0.42 ± 0.45, P=1 | -0.73 ± 0.73, P=1 | -0.5 ± 0.24, P=1 | 0.06 ± 0.33, P=1 |
| **rh_rostralmiddlefrontal** | 0.54 ± 0.36, P=1 | 0.4 ± 0.12, P=0.067 | -0.4 ± 0.3, P=1 | -0.83 ± 0.5, P=1 | -0.48 ± 0.16, P=0.193 | -0.46 ± 0.22, P=1 |
| **rh_superiorfrontal** | 0.53 ± 0.41, P=1 | 0.5 ± 0.14, P=0.026 | -0.16 ± 0.35, P=1 | -1.09 ± 0.57, P=1 | -0.64 ± 0.18, P=0.039 | -0.29 ± 0.25, P=1 |
| **rh_superiorparietal** | 1.03 ± 0.36, P=0.293 | 0.4 ± 0.12, P=0.09 | -0.4 ± 0.31, P=1 | -0.98 ± 0.5, P=1 | -0.67 ± 0.16, P=0.002 | -0.79 ± 0.22, P=0.024 |

| | | | | | | |
|---|---|---|---|---|---|---|
| **rh_superiortemporal** | 2.91 ± 0.51, P<0.001 | 1.66 ± 0.17, P<0.001 | -3.05 ± 0.44, P<0.001 | -5.6 ± 0.71, P<0.001 | -1.19 ± 0.23, P<0.001 | -0.91 ± 0.32, P=0.308 |
| **rh_supramarginal** | 1.93 ± 0.38, P<0.001 | 1.03 ± 0.13, P<0.001 | -1.73 ± 0.33, P<0.001 | -3.57 ± 0.53, P<0.001 | -0.9 ± 0.17, P<0.001 | -0.65 ± 0.24, P=0.413 |
| **rh_temporalpole** | 4.67 ± 1, P<0.001 | 2.66 ± 0.33, P<0.001 | -4.91 ± 0.85, P<0.001 | -8.98 ± 1.38, P<0.001 | -1.86 ± 0.46, P=0.003 | -1.82 ± 0.62, P=0.247 |
| **rh_transversetemporal** | 3.08 ± 0.57, P<0.001 | 1.39 ± 0.2, P<0.001 | -2.36 ± 0.49, P<0.001 | -5.33 ± 0.79, P<0.001 | -0.79 ± 0.26, P=0.153 | -0.86 ± 0.35, P=1 |
| **lh_MeanThickness** | 1.93 ± 0.33, P<0.001 | 0.7 ± 0.11, P<0.001 | -1.22 ± 0.28, P=0.001 | -2.17 ± 0.45, P<0.001 | -0.72 ± 0.15, P<0.001 | -0.47 ± 0.2, P=1 |
| **rh_MeanThickness** | 1.65 ± 0.33, P<0.001 | 0.78 ± 0.11, P<0.001 | -1.22 ± 0.28, P=0.001 | -2.51 ± 0.46, P<0.001 | -0.76 ± 0.15, P<0.001 | -0.64 ± 0.21, P=0.138 |

The estimates indicate the cortical thickness for that region preserved (positive values) or lost (negative values) per year for each unit increase in the marker assessed at the baseline. Units were 100 for WM-PVS count, 10 for BG-PVS count, 0.1 mm for WM- and BG-PVS diameter, and 1 log for P-WML and D-WML volume. See also Figure 2B. Parcellations performed by Freesurfer according to the Desikan-Killany atlas. Bankssts: Banks of the Superior Temporal Sulcus; Lh: left hemisphere; Rh: right hemisphere. Models were fully adjusted for all the same covariates included in the main linear mixed-effect model. P values are corrected for 70 comparisons with Holm-Bonferroni procedure.

**Table S12. Regional estimates (mm$^3$/year) and adjusted significance of the interaction term "marker" by "time" in linear mixed-effects models with the white matter volume of the corresponding region as dependent variable.**

| Region | WM-PVS count | BG-PVS count | WM-PVS diameter | BG-PVS diameter | P-WML log volume | D-WML log volume |
|---|---|---|---|---|---|---|
| lh_bankssts | 3.41 ± 0.55, P<0.001 | 0.67 ± 0.17, P=0.005 | -0.44 ± 0.44, P=1 | -1.47 ± 0.71, P=1 | -0.82 ± 0.23, P=0.028 | -0.15 ± 0.32, P=1 |
| lh_caudalanteriorcingulate | 4.32 ± 0.85, P<0.001 | 1.24 ± 0.26, P<0.001 | -0.35 ± 0.67, P=1 | -2.62 ± 1.09, P=1 | -0.94 ± 0.36, P=0.614 | -0.99 ± 0.49, P=1 |
| lh_caudalmiddlefrontal | 7.82 ± 1.14, P<0.001 | 1.47 ± 0.36, P=0.003 | 1.15 ± 0.91, P=1 | -0.77 ± 1.48, P=1 | -1.06 ± 0.48, P=1 | 0.25 ± 0.66, P=1 |
| lh_cuneus | 6.73 ± 0.86, P<0.001 | 2.1 ± 0.27, P<0.001 | -2.39 ± 0.68, P=0.031 | -5.81 ± 1.11, P<0.001 | -1.32 ± 0.36, P=0.017 | -0.11 ± 0.49, P=1 |
| lh_entorhinal | 2.4 ± 0.37, P<0.001 | 0.57 ± 0.11, P<0.001 | -0.87 ± 0.29, P=0.18 | -1.94 ± 0.47, P=0.003 | -0.49 ± 0.15, P=0.107 | 0.17 ± 0.21, P=1 |
| lh_frontalpole | 0.95 ± 0.21, P=0.001 | 0.21 ± 0.07, P=0.151 | -0.43 ± 0.17, P=0.703 | -0.4 ± 0.28, P=1 | -0.11 ± 0.09, P=1 | -0.15 ± 0.12, P=1 |
| lh_fusiform | 14.5 ± 1.5, P<0.001 | 3.25 ± 0.48, P<0.001 | -2.96 ± 1.21, P=1 | -8.63 ± 1.97, P=0.001 | -2.47 ± 0.64, P=0.009 | -0.28 ± 0.88, P=1 |
| lh_inferiorparietal | 14.3 ± 2.12, P<0.001 | 3.27 ± 0.67, P<0.001 | -2.69 ± 1.69, P=1 | -9.9 ± 2.74, P=0.021 | -1.31 ± 0.9, P=1 | -0.46 ± 1.22, P=1 |
| lh_inferiortemporal | 16.75 ± 1.65, P<0.001 | 3.75 ± 0.52, P<0.001 | -5.95 ± 1.34, P=0.001 | -12.98 ± 2.17, P<0.001 | -2.48 ± 0.71, P=0.036 | -0.75 ± 0.97, P=1 |
| lh_insula | 7.6 ± 1.39, P<0.001 | 2.08 ± 0.43, P<0.001 | -2.42 ± 1.09, P=1 | -4.98 ± 1.77, P=0.345 | -3.05 ± 0.58, P<0.001 | -1.51 ± 0.79, P=1 |
| lh_isthmuscingulate | 6.37 ± 0.71, P<0.001 | 1.59 ± 0.22, P<0.001 | -0.79 ± 0.57, P=1 | -4.27 ± 0.92, P<0.001 | -1.37 ± 0.3, P<0.001 | -0.15 ± 0.41, P=1 |
| lh_lateraloccipital | 25.09 ± 2.75, P<0.001 | 7.68 ± 0.87, P<0.001 | -12.72 ± 2.18, P<0.001 | -24.8 ± 3.57, P<0.001 | -4.65 ± 1.17, P=0.005 | -2.02 ± 1.59, P=1 |
| lh_lateralorbitofrontal | 11.53 ± 1.31, P<0.001 | 2.65 ± 0.41, P<0.001 | -1.7 ± 1.05, P=1 | -6.45 ± 1.71, P=0.011 | -1.77 ± 0.56, P=0.107 | -0.38 ± 0.76, P=1 |
| lh_lingual | 12.16 ± 1.53, P<0.001 | 3.51 ± 0.48, P<0.001 | -3.92 ± 1.21, P=0.08 | -9.81 ± 1.97, P<0.001 | -2.31 ± 0.64, P=0.021 | -0.01 ± 0.87, P=1 |
| lh_medialorbitofrontal | 6.51 ± 0.92, P<0.001 | 1.29 ± 0.29, P=0.001 | 1.52 ± 0.73, P=1 | -1.57 ± 1.2, P=1 | -0.74 ± 0.39, P=1 | 0.99 ± 0.53, P=1 |
| lh_middletemporal | 12.6 ± 1.5, P<0.001 | 3.33 ± 0.47, P<0.001 | -4.78 ± 1.21, P=0.005 | -10.63 ± 1.96, P<0.001 | -1.28 ± 0.65, P=1 | -0.37 ± 0.88, P=1 |
| lh_paracentral | 5.64 ± 0.98, P<0.001 | 0.72 ± 0.31, P=1 | 1.77 ± 0.77, P=1 | 0.61 ± 1.26, P=1 | -0.14 ± 0.41, P=1 | 0.83 ± 0.56, P=1 |
| lh_parahippocampal | 2.2 ± 0.42, P<0.001 | 0.67 ± 0.13, P<0.001 | -1.2 ± 0.33, P=0.017 | -1.94 ± 0.53, P=0.019 | -0.33 ± 0.17, P=1 | 0.19 ± 0.24, P=1 |
| lh_parsopercularis | 4.69 ± 0.92, P<0.001 | 0.97 ± 0.29, P=0.05 | 0.52 ± 0.73, P=1 | -1.1 ± 1.18, P=1 | 0.25 ± 0.39, P=1 | 0.57 ± 0.53, P=1 |
| lh_parsorbitalis | 2.12 ± 0.4, P<0.001 | 0.52 ± 0.13, P=0.003 | -0.76 ± 0.31, P=1 | -1.47 ± 0.51, P=0.292 | -0.12 ± 0.17, P=1 | 0.07 ± 0.23, P=1 |
| lh_parstriangularis | 5.06 ± 0.77, P<0.001 | 0.91 ± 0.24, P=0.011 | -1.25 ± 0.61, P=1 | -2.51 ± 0.99, P=0.801 | -0.49 ± 0.33, P=1 | 0.38 ± 0.44, P=1 |
| lh_pericalcarine | 5.53 ± 0.88, P<0.001 | 1.19 ± 0.28, P=0.001 | -1.27 ± 0.69, P=1 | -3.5 ± 1.13, P=0.134 | -0.67 ± 0.37, P=1 | 0.45 ± 0.5, P=1 |
| lh_postcentral | 11.37 ± 1.95, P<0.001 | 1.84 ± 0.63, P=0.237 | 0.86 ± 1.54, P=1 | -0.98 ± 2.53, P=1 | 0.05 ± 0.81, P=1 | 2.05 ± 1.1, P=1 |
| lh_posteriorcingulate | 8.58 ± 1.17, P<0.001 | 2.44 ± 0.35, P<0.001 | -1.93 ± 0.93, P=1 | -5.41 ± 1.51, P=0.023 | -2.23 ± 0.5, P=0.001 | -1.29 ± 0.68, P=1 |
| lh_precentral | 20.53 ± 3.05, P<0.001 | 3.42 ± 0.97, P=0.03 | 4.2 ± 2.43, P=1 | 0.56 ± 3.98, P=1 | -2.78 ± 1.28, P=1 | 3.01 ± 1.75, P=1 |
| lh_precuneus | 12.86 ± 1.84, P<0.001 | 2.31 ± 0.58, P=0.005 | 1.41 ± 1.47, P=1 | -4.67 ± 2.39, P=1 | -1.9 ± 0.78, P=0.998 | 0.75 ± 1.06, P=1 |
| lh_rostralanteriorcingulate | 3.37 ± 0.52, P<0.001 | 0.82 ± 0.16, P<0.001 | -0.04 ± 0.41, P=1 | -1.84 ± 0.67, P=0.406 | -0.45 ± 0.22, P=1 | -0.47 ± 0.3, P=1 |
| lh_rostralmiddlefrontal | 16.78 ± 2.88, P<0.001 | 3.93 ± 0.9, P=0.001 | -0.61 ± 2.29, P=1 | -3.79 ± 3.74, P=1 | -1.41 ± 1.22, P=1 | 0.62 ± 1.66, P=1 |
| lh_superiorfrontal | 23.88 ± 4.15, P<0.001 | 3.41 ± 1.31, P=0.619 | 6.11 ± 3.31, P=1 | 3.53 ± 5.4, P=1 | -0.78 ± 1.76, P=1 | 2.37 ± 2.39, P=1 |
| lh_superiorparietal | 19.46 ± 2.67, P<0.001 | 4.75 ± 0.84, P<0.001 | -1.73 ± 2.14, P=1 | -9.56 ± 3.48, P=0.413 | -3.14 ± 1.13, P=0.382 | 0.33 ± 1.55, P=1 |

| | | | | | | |
|---|---|---|---|---|---|---|
| **lh_superiortemporal** | 15.38 ± 1.64, P<0.001 | 3.93 ± 0.52, P<0.001 | -2.75 ± 1.32, P=1 | -9.05 ± 2.15, P=0.002 | -1.54 ± 0.7, P=1 | 0.23 ± 0.96, P=1 |
| **lh_supramarginal** | 13.78 ± 1.85, P<0.001 | 2.77 ± 0.59, P<0.001 | -1.18 ± 1.49, P=1 | -6.09 ± 2.43, P=0.836 | -1.47 ± 0.79, P=1 | 0.72 ± 1.08, P=1 |
| **lh_temporalpole** | 2.75 ± 0.4, P<0.001 | 0.66 ± 0.12, P<0.001 | -1.5 ± 0.31, P<0.001 | -2.68 ± 0.51, P<0.001 | -0.48 ± 0.17, P=0.274 | -0.09 ± 0.23, P=1 |
| **lh_transversetemporal** | 0.42 ± 0.3, P=1 | -0.06 ± 0.09, P=1 | 0.56 ± 0.23, P=1 | 0.54 ± 0.38, P=1 | 0.34 ± 0.12, P=0.361 | 0.19 ± 0.17, P=1 |
| **rh_bankssts** | 4.59 ± 0.57, P<0.001 | 1.05 ± 0.17, P<0.001 | -1.08 ± 0.45, P=1 | -1.93 ± 0.73, P=0.577 | -0.88 ± 0.24, P=0.018 | -0.34 ± 0.33, P=1 |
| **rh_caudalanteriorcingulate** | 4.03 ± 0.85, P<0.001 | 0.45 ± 0.26, P=1 | -0.48 ± 0.67, P=1 | 0.03 ± 1.08, P=1 | -0.49 ± 0.36, P=1 | -0.61 ± 0.49, P=1 |
| **rh_caudalmiddlefrontal** | 6.12 ± 1.1, P<0.001 | 0.92 ± 0.35, P=0.546 | 1.04 ± 0.87, P=1 | 1.25 ± 1.43, P=1 | -1.07 ± 0.46, P=1 | 0.26 ± 0.63, P=1 |
| **rh_cuneus** | 6.17 ± 0.83, P<0.001 | 1.8 ± 0.26, P<0.001 | -0.64 ± 0.66, P=1 | -3.39 ± 1.07, P=0.111 | -1.34 ± 0.35, P=0.008 | 0.15 ± 0.47, P=1 |
| **rh_entorhinal** | 2.31 ± 0.34, P<0.001 | 0.53 ± 0.11, P<0.001 | -0.75 ± 0.27, P=0.327 | -1.42 ± 0.44, P=0.075 | -0.38 ± 0.14, P=0.509 | -0.22 ± 0.19, P=1 |
| **rh_frontalpole** | 1.53 ± 0.27, P<0.001 | 0.46 ± 0.09, P<0.001 | -1.03 ± 0.21, P<0.001 | -1.22 ± 0.35, P=0.04 | -0.35 ± 0.11, P=0.173 | -0.32 ± 0.16, P=1 |
| **rh_fusiform** | 14.68 ± 1.42, P<0.001 | 3.1 ± 0.45, P<0.001 | -2.98 ± 1.16, P=0.702 | -6.34 ± 1.89, P=0.055 | -3.36 ± 0.61, P<0.001 | -0.89 ± 0.84, P=1 |
| **rh_inferiorparietal** | 23.49 ± 2.55, P<0.001 | 5.05 ± 0.82, P<0.001 | -7.48 ± 2.07, P=0.022 | -14.86 ± 3.36, P=0.001 | -3.08 ± 1.1, P=0.363 | -0.63 ± 1.51, P=1 |
| **rh_inferiortemporal** | 15.48 ± 1.56, P<0.001 | 3.12 ± 0.5, P<0.001 | -5.1 ± 1.28, P=0.005 | -8.77 ± 2.07, P=0.002 | -2.74 ± 0.68, P=0.004 | -0.89 ± 0.93, P=1 |
| **rh_insula** | 4.55 ± 1.26, P=0.02 | 1.22 ± 0.39, P=0.113 | -1.76 ± 0.98, P=1 | -0.93 ± 1.6, P=1 | -1.88 ± 0.52, P=0.021 | -1.32 ± 0.71, P=1 |
| **rh_isthmuscingulate** | 4.04 ± 0.77, P<0.001 | 0.9 ± 0.24, P=0.012 | 0.1 ± 0.61, P=1 | -1.14 ± 1, P=1 | -1 ± 0.33, P=0.158 | 0.23 ± 0.45, P=1 |
| **rh_lateraloccipital** | 29.68 ± 2.68, P<0.001 | 7.59 ± 0.86, P<0.001 | -11.84 ± 2.17, P<0.001 | -22.17 ± 3.54, P<0.001 | -5.87 ± 1.15, P<0.001 | -1.17 ± 1.58, P=1 |
| **rh_lateralorbitofrontal** | 9.87 ± 1.3, P<0.001 | 2.07 ± 0.41, P<0.001 | -0.11 ± 1.04, P=1 | -2.06 ± 1.7, P=1 | -2.23 ± 0.55, P=0.004 | 0.21 ± 0.76, P=1 |
| **rh_lingual** | 12.35 ± 1.49, P<0.001 | 3.97 ± 0.47, P<0.001 | -2.66 ± 1.19, P=1 | -8.88 ± 1.94, P<0.001 | -2.66 ± 0.62, P=0.001 | -1.09 ± 0.85, P=1 |
| **rh_medialorbitofrontal** | 8.24 ± 0.97, P<0.001 | 1.96 ± 0.31, P<0.001 | -1.47 ± 0.78, P=1 | -3.36 ± 1.27, P=0.565 | -1.41 ± 0.41, P=0.043 | 0.3 ± 0.56, P=1 |
| **rh_middletemporal** | 15.73 ± 1.77, P<0.001 | 3.75 ± 0.56, P<0.001 | -7.46 ± 1.43, P<0.001 | -11.3 ± 2.32, P<0.001 | -2.2 ± 0.77, P=0.284 | -1.28 ± 1.05, P=1 |
| **rh_paracentral** | 5.82 ± 1.03, P<0.001 | 0.63 ± 0.33, P=1 | 2.2 ± 0.81, P=0.481 | 2 ± 1.33, P=1 | -0.9 ± 0.43, P=1 | 0.79 ± 0.59, P=1 |
| **rh_parahippocampal** | 2.83 ± 0.39, P<0.001 | 0.95 ± 0.12, P<0.001 | -1.48 ± 0.31, P<0.001 | -2.33 ± 0.5, P<0.001 | -0.77 ± 0.16, P<0.001 | -0.37 ± 0.22, P=1 |
| **rh_parsopercularis** | 4.98 ± 0.8, P<0.001 | 1.16 ± 0.25, P<0.001 | -0.52 ± 0.64, P=1 | -2.26 ± 1.03, P=1 | -0.8 ± 0.34, P=1 | 0.08 ± 0.46, P=1 |
| **rh_parsorbitalis** | 2.97 ± 0.49, P<0.001 | 0.97 ± 0.16, P<0.001 | -1.84 ± 0.39, P<0.001 | -2.97 ± 0.64, P<0.001 | -0.39 ± 0.21, P=1 | -0.28 ± 0.28, P=1 |
| **rh_parstriangularis** | 5.64 ± 0.91, P<0.001 | 1.39 ± 0.28, P<0.001 | -1.99 ± 0.72, P=0.414 | -3.62 ± 1.18, P=0.147 | -0.76 ± 0.39, P=1 | 0.01 ± 0.53, P=1 |
| **rh_pericalcarine** | 4.95 ± 0.79, P<0.001 | 1.27 ± 0.26, P<0.001 | -0.71 ± 0.62, P=1 | -2.16 ± 1.03, P=1 | -0.82 ± 0.33, P=0.828 | 0.29 ± 0.45, P=1 |
| **rh_postcentral** | 12.42 ± 2.06, P<0.001 | 1.47 ± 0.66, P=1 | 0.01 ± 1.63, P=1 | 0.13 ± 2.68, P=1 | -0.8 ± 0.86, P=1 | 0.85 ± 1.17, P=1 |
| **rh_posteriorcingulate** | 7.06 ± 1.03, P<0.001 | 1.8 ± 0.31, P<0.001 | -0.97 ± 0.82, P=1 | -2.26 ± 1.33, P=1 | -2.04 ± 0.44, P<0.001 | -0.73 ± 0.6, P=1 |
| **rh_precentral** | 19.91 ± 3.01, P<0.001 | 2.99 ± 0.96, P=0.127 | 3.08 ± 2.39, P=1 | 0.82 ± 3.92, P=1 | -2.48 ± 1.26, P=1 | 1.67 ± 1.72, P=1 |
| **rh_precuneus** | 15.5 ± 1.73, P<0.001 | 3.21 ± 0.55, P<0.001 | 0.13 ± 1.4, P=1 | -3.04 ± 2.29, P=1 | -2.38 ± 0.74, P=0.095 | 0.45 ± 1.01, P=1 |
| **rh_rostralanteriorcingulate** | 2.69 ± 0.45, P<0.001 | 0.41 ± 0.14, P=0.251 | -0.5 ± 0.36, P=1 | -0.46 ± 0.58, P=1 | -0.48 ± 0.19, P=0.776 | -0.02 ± 0.26, P=1 |
| **rh_rostralmiddlefrontal** | 21.01 ± 3, P<0.001 | 5.78 ± 0.94, P<0.001 | -6.93 ± 2.4, P=0.27 | -13.51 ± 3.9, P=0.037 | -4.3 ± 1.28, P=0.054 | -1.29 ± 1.74, P=1 |
| **rh_superiorfrontal** | 26.24 ± 3.99, P<0.001 | 4.8 ± 1.26, P=0.01 | 0.5 ± 3.21, P=1 | -0.54 ± 5.23, P=1 | -4.27 ± 1.7, P=0.836 | -0.08 ± 2.32, P=1 |
| **rh_superiorparietal** | 22.44 ± 2.39, P<0.001 | 5.36 ± 0.76, P<0.001 | -4.3 ± 1.93, P=1 | -12.29 ± 3.14, P=0.007 | -3.84 ± 1.02, P=0.012 | 0.42 ± 1.39, P=1 |

| | | | | | | |
|---|---|---|---|---|---|---|
| **rh_superiortemporal** | 14.03 ± 1.76, P<0.001 | 3.41 ± 0.55, P<0.001 | -4.63 ± 1.42, P=0.078 | -7.77 ± 2.31, P=0.055 | -2.28 ± 0.76, P=0.181 | -0.73 ± 1.04, P=1 |
| **rh_supramarginal** | 12.76 ± 1.73, P<0.001 | 2.71 ± 0.55, P<0.001 | -2.73 ± 1.4, P=1 | -5.7 ± 2.28, P=0.847 | -2.22 ± 0.74, P=0.189 | 0.33 ± 1.01, P=1 |
| **rh_temporalpole** | 1.86 ± 0.34, P<0.001 | 0.49 ± 0.11, P<0.001 | -0.82 ± 0.27, P=0.171 | -1.43 ± 0.44, P=0.083 | -0.51 ± 0.14, P=0.023 | -0.3 ± 0.19, P=1 |
| **rh_transversetemporal** | 0.4 ± 0.26, P=1 | 0.06 ± 0.08, P=1 | 0.4 ± 0.2, P=1 | 0.08 ± 0.33, P=1 | 0.05 ± 0.11, P=1 | -0.12 ± 0.15, P=1 |

The estimates indicate the white matter volume for that region preserved (positive values) or lost (negative values) per year for each unit increase in the marker assessed at the baseline. Units were 100 for WM-PVS count, 10 for BG-PVS count, 0.1 mm for WM- and BG-PVS diameter, and 1 log for P-WML and D-WML volume. See also Figure 2C. Parcellations performed by Freesurfer according to the Desikan-Killany atlas. Bankssts: Banks of the Superior Temporal Sulcus; Lh: left hemisphere; Rh: right hemisphere. Models were fully adjusted for all the same covariates included in the main linear mixed-effect model. P values are corrected for 68 comparisons with Holm-Bonferroni procedure.

**Table S13. Sensitivity analysis on the interaction term “marker” by “time” in linear mixed-effects models with the grey matter volume, cortical thickness, or white matter volume as dependent variables.**

| Marker | Covariate | Grey matter volume ($mm^3$ / year) | Cortical thickness (µm / year) | White matter volume ($mm^3$ / year) |
|---|---|---|---|---|
| WM-PVS count (per 100-unit) | Artifacts | 720±100, P<0.001 | 4±1, P<0.001 | 582±84, P<0.001 |
| | APOE | 696±99, P<0.001 | 4±1, P<0.001 | 605±84, P<0.001 |
| | MMSE | 812±104, P<0.001 | 4±1, P<0.001 | 645±88, P<0.001 |
| | Tobacco | 728±101, P<0.001 | 4±1, P<0.001 | 596±85, P<0.001 |
| | Abeta | 561±113, P<0.001 | 3±1, P<0.001 | 744±84, P<0.001 |
| | Tau | 413±112, P<0.001 | 3±1, P=0.001 | 550±79, P<0.001 |
| BG-PVS count (per 10-unit) | Artifacts | 190±33, P<0.001 | 1±0, P<0.001 | 143±26, P<0.001 |
| | APOE | 178±33, P<0.001 | 1±0, P<0.001 | 140±26, P<0.001 |
| | MMSE | 255±35, P<0.001 | 2±0, P<0.001 | 170±27, P<0.001 |
| | Tobacco | 190±33, P<0.001 | 1±0, P<0.001 | 142±26, P<0.001 |
| | Abeta | 161±35, P<0.001 | 1±0, P<0.001 | 151±25, P<0.001 |
| | Tau | 88±36, P=0.031 | 1±0, P<0.001 | 189±24, P<0.001 |
| WM-PVS diameter (per 0.1 mm) | Artifacts | -374±84, P<0.001 | -2±1, P<0.001 | -64±67, P=0.671 |
| | APOE | -341±83, P<0.001 | -2±1, P<0.001 | -71±67, P=0.57 |
| | MMSE | -463±87, P<0.001 | -3±1, P<0.001 | -93±70, P=0.37 |
| | Tobacco | -378±85, P<0.001 | -3±1, P<0.001 | -58±67, P=0.775 |
| | Abeta | -251±90, P=0.011 | -2±1, P=0.001 | 73±66, P=0.526 |
| | Tau | -170±91, P=0.123 | -2±1, P=0.022 | -15±62, P=1 |
| BG-PVS diameter (per 0.1 mm) | Artifacts | -571±136, P<0.001 | -5±1, P<0.001 | -282±108, P=0.019 |
| | APOE | -543±136, P<0.001 | -4±1, P<0.001 | -274±108, P=0.023 |
| | MMSE | -726±141, P<0.001 | -6±1, P<0.001 | -334±114, P=0.007 |
| | Tobacco | -577±138, P<0.001 | -5±1, P<0.001 | -278±109, P=0.022 |
| | Abeta | -338±146, P=0.041 | -4±1, P<0.001 | -202±106, P=0.112 |
| | Tau | -177±145, P=0.444 | -2±1, P=0.042 | -311±98, P=0.003 |
| P-WML log volume | Artifacts | -277±45, P<0.001 | -1±0, P<0.001 | -71±36, P=0.09 |
| | APOE | -274±45, P<0.001 | -1±0, P<0.001 | -71±36, P=0.089 |
| | MMSE | -283±48, P<0.001 | -1±0, P<0.001 | -81±38, P=0.066 |
| | Tobacco | -285±46, P<0.001 | -2±0, P<0.001 | -68±36, P=0.118 |
| | Abeta | -208±48, P<0.001 | -1±0, P<0.001 | -25±34, P=0.91 |
| | Tau | -227±47, P<0.001 | -2±0, P<0.001 | -61±31, P=0.093 |
| D-WML log volume | Artifacts | -185±61, P=0.005 | -1±0, P=0.01 | 1±48, P=1 |
| | APOE | -165±61, P=0.014 | -1±0, P=0.028 | 7±48, P=1 |
| | MMSE | -144±65, P=0.053 | -1±0, P=0.067 | 2±52, P=1 |
| | Tobacco | -191±62, P=0.004 | -1±0, P=0.013 | -1±49, P=1 |
| | Abeta | -157±65, P=0.03 | -1±0, P=0.052 | 69±46, P=0.259 |
| | Tau | -151±62, P=0.031 | -1±0, P=0.088 | 57±41, P=0.326 |

Estimates indicate the value of the dependent variable preserved (positive values) or lost (negative values) per year for each unit increase in the marker. All models were adjusted for the same covariates as in all the other main models. "P" indicates adjusted P-values.

**Table S14. Estimated marginal means for baseline PVS and WML markers in non-demented individuals who converted to dementia versus non-converters.**

| Marker (at baseline MRI) | Estimated marginal mean ± standard error | | Adj. P value |
|---|---|---|---|
| | Non-converter | Converter | |
| WM-PVS count | 453 ± 7 | 415 ± 8 | <0.001 |
| BG-PVS count | 143 ± 2 | 127 ± 2 | <0.001 |
| WM-PVS diameter (mm) | 1.96 ± 0.01 | 1.98 ± 0.01 | <0.001 |
| BG-PVS diameter (mm) | 1.58 ± 0 | 1.6 ± 0.01 | <0.001 |
| P-WML log volume | 4.28 ± 0.15 | 5.22 ± 0.17 | <0.001 |
| D-WML log volume | 2.5 ± 0.11 | 2.51 ± 0.13 | 1 |

Marginal means were estimated controlling for the same covariates as in the primary analysis (Table S2).

**Table S15. Estimates and adjusted significance of the logistic regression models assessing the association of medium- and high-risk tertiles with amyloid and tau positive status controlling for demographic and clinical covariates.**

| Tertile | Marker | Amyloid-β + status | Tau + status |
|---|---|---|---|
| Medium-risk + High-risk (compared to Low-risk) | WM-PVS count | 0.12 ± 0.1, P=0.489 | 0.01 ± 0.12, P=1 |
| | BG-PVS count | 0.18 ± 0.1, P=0.16 | 0.15 ± 0.13, P=0.444 |
| | WM-PVS diameter | 0.13 ± 0.1, P=0.352 | 0.17 ± 0.12, P=0.318 |
| | BG-PVS diameter | 0.15 ± 0.1, P=0.236 | 0.13 ± 0.12, P=0.57 |
| | P-WML volume | 0.27 ± 0.1, P=0.017 | -0.24 ± 0.12, P=0.096 |
| | D-WML volume | 0.16 ± 0.09, P=0.193 | -0.16 ± 0.11, P=0.33 |
| | Total grey matter volume | 0.28 ± 0.1, P=0.013 | 0.07 ± 0.13, P=1 |
| | Mean cortical thickness | 0.49 ± 0.11, P<0.001 | 0.17 ± 0.11, P=0.227 |
| High-risk only (compared to Low-risk + Medium-risk) | WM-PVS count | 0.01 ± 0.09, P=1 | -0.04 ± 0.11, P=1 |
| | BG-PVS count | 0.12 ± 0.09, P=0.408 | 0.01 ± 0.12, P=1 |
| | WM-PVS diameter | -0.05 ± 0.09, P=1 | -0.17 ± 0.1, P=0.199 |
| | BG-PVS diameter | 0.07 ± 0.09, P=0.867 | 0.13 ± 0.12, P=0.57 |
| | P-WML volume | 0.03 ± 0.08, P=1 | -0.07 ± 0.1, P=0.965 |
| | D-WML volume | 0.05 ± 0.08, P=1 | -0.32 ± 0.1, P=0.003 |
| | Total grey matter volume | 0.3 ± 0.11, P=0.01 | -0.14 ± 0.13, P=0.554 |
| | Mean cortical thickness | 0.25 ± 0.09, P=0.009 | 0.01 ± 0.12, P=1 |

**Table S16. Tertile limits for PVS and WML markers.**

| Marker | Low-risk tertile limits | Medium-risk tertile limits | High-risk tertile limits |
|---|---|---|---|
| WM-PVS count | 840 to 498 | 497 to 377 | 376 to 86 |
| BG-PVS count | 272 to 155 | 154 to 117 | 116 to 34 |
| WM-PVS diameter (mm) | 1.69 to 1.9 | 1.91 to 2.03 | 2.04 to 2.37 |
| BG-PVS diameter (mm) | 1.37 to 1.53 | 1.54 to 1.61 | 1.62 to 1.83 |
| P-WML volume (log) | 0 to 2.83 | 2.84 to 6.74 | 6.75 to 9.56 |
| D-WML volume (log) | 0 to 1.1 | 1.11 to 3.81 | 3.82 to 6.7 |

**Table S17. Baseline characteristics of the participants whose brain MRI data have been used for the robust vessel segmentation method development and validation.**

| Dataset | Cohort | N. subjects | Age – yr | Female – % | Body Mass Index – kg/m$^2$ |
|---|---|---|---|---|---|
| Test-Retest | MarkVCID | 39 | 71.6 ± 10.1 | 69.2 | NA |
| Inter-scanner | MarkVCID | 19 | 68.5 ± 9.3 | 52.6 | NA |
| Inter-field-strength | ADNI | 115 | 74.6 ± 6.6 | 51.3 | 26.3 ± 4.5 |
| Biological validation | HCP | 2163 | 33.1 ± 17.8 | 55.4 | 25.4 ± 5.6* |

*Body Mass index was available in 2160 subjects.

**Table S18. Overview of the MRI scanners and T1-weighted image parameters employed to acquire the data used for the robust vessel segmentation method development and validation.**

| FS (T) | Manuf. | Model | TR (sec) | TE (msec) | TI (sec) | FA (°) | Native voxel vol. ($mm^3$) | N. scans analyzed | Cohort |
|---|---|---|---|---|---|---|---|---|---|
| **Test-retest dataset** | | | | | | | | | |
| 3 | Philips | Achieva dStream | 0.0095 | 1.8 / 3.7 / 5.5 / 7.3 | NA | 7 | 1 | 12 | MarkVCID |
| 3 | Siemens | Prisma | 2.53 | 1.7 / 3.6 / 5.4 / 7.3 | 1.1 | 7 | 1 | 12 | MarkVCID |
| 3 | Siemens | Prisma fit | 2.53 | 1.7 / 3.6 / 5.4 / 7.3 | 1.1 | 7 | 1 | 40 | MarkVCID |
| 3 | Siemens | TrioTim | 2.53 | 1.6 / 3.5 / 5.3 / 7.2 | 1.2 | 7 | 1 | 14 | MarkVCID |
| **Inter-scanner dataset** | | | | | | | | | |
| 3 | GE | DISCOVERY MR750w | 0.0095 | 3.7 | 0.6 | 10 | 0.5 | 19 | MarkVCID |
| 3 | Philips | Achieva dStream | 0.0095 | 1.8 / 3.7 / 5.5 / 7.3 | NA | 7 | 1 | 19 | MarkVCID |
| 3 | Siemens | Prisma fit | 2.53 | 1.7 / 3.6 / 5.4 / 7.3 | 1.1 | 7 | 1 | 19 | MarkVCID |
| 3 | Siemens | TrioTim | 2.53 | 1.6 / 3.5 / 5.3 / 7.2 | 1.2 | 7 | 1 | 19 | MarkVCID |
| **Inter-field-strength dataset** | | | | | | | | | |
| 1.5 | GE | GENESIS SIGNA | 0.01 | 4.1 | 1 | 8 | 1.1 | 47 | ADNI |
| 1.5 | GE | SIGNA EXCITE | 0.0086 to 0.0092 | 3.8 to 4.1 | 1 | 8 | 1.1 | 93 | ADNI |
| 1.5 | GE | SIGNA HDx | 0.0086 to 0.0088 | 3.8 to 3.9 | 1 | 8 | 1.1 | 30 | ADNI |
| 1.5 | GE | Signa HDxt | 0.0086 | 3.8 | 1 | 8 | 1.1 | 4 | ADNI |
| 1.5 | Philips | Achieva | 0.0086 | 4 | NA | 8 | 1.1 | 8 | ADNI |
| 1.5 | Philips | Gyroscan NT | 0.0086 | 4 | 1 | 8 | 1.1 | 5 | ADNI |
| 1.5 | Philips | Intera | 0.0085 to 0.0086 | 4 | 1 | 8 | 1.1 | 34 | ADNI |
| 1.5 | Siemens | Avanto | 2.4 | 3.5 | 1 | 8 | 1.9 | 6 | ADNI |
| 1.5 | Siemens | Espree | 2.4 | 3.6 | 1 | 8 | 1.9 | 3 | ADNI |
| 1.5 | Siemens | Sonata | 3 | 3.5 | 1 | 8 | 1.9 | 9 | ADNI |
| 1.5 | Siemens | Symphony | 3 | 3.9 | 1 | 8 | 1.9 | 13 | ADNI |
| 1.5 | Siemens | Symphony Tim | 3 | 3.6 | 1 | 8 | 1.9 | 47 | ADNI |
| 3 | GE | GENESIS SIGNA | 0.0075 | 3.1 | 0.9 | 8 | 1.2 | 9 | ADNI |
| 3 | GE | SIGNA EXCITE | 0.0066 to 0.0070 | 2.8 to 2.9 | 0.9 | 8 | 1.2 | 15 | ADNI |
| 3 | GE | SIGNA HDx | 0.0066 | 2.8 | 0.9 | 8 | 1.2 | 20 | ADNI |
| 3 | GE | Signa HDxt | 0.0066 | 2.8 | 0.9 | 8 | 1.2 | 1 | ADNI |
| 3 | Philips | Achieva | 0.0066 to 0.0069 | 3.1 to 3.2 | 0.85 | 8 | 1.2 | 47 | ADNI |
| 3 | Philips | Intera | 0.0067 to 0.007 | 3.1 to 3.3 | 0.85 | 8 | 1.2 | 61 | ADNI |
| 3 | Siemens | Allegra | 2.3 | 2.9 | 0.9 | 9 | 1.2 | 7 | ADNI |
| 3 | Siemens | Trio | 2.3 | 2.9 | 0.9 | 9 | 1.2 | 90 | ADNI |
| 3 | Siemens | TrioTim | 2.3 | 2.9 | 0.9 | 9 | 1.2 | 49 | ADNI |
| **Biological validation dataset** | | | | | | | | | |
| 3 | Siemens | Prisma | 2.5 | 1.8 / 3.6 / 5.4 / 7.2 | 1 | 8 | 0.512 | 1096 | HCPA/HCPD |
| 3 | Siemens | Connectome Skyra | 2.4 | 2.1 | 1 | 8 | 0.343 | 1067 | HCPYA |
